# Psychological interventions for adult PTSD: A network and pairwise meta-analysis of short and long-term efficacy, acceptability and trial quality

**DOI:** 10.1101/2022.05.03.22274616

**Authors:** Thole H. Hoppen, Marvin Jehn, Heinz Holling, Julian Mutz, Ahlke Kip, Nexhmedin Morina

## Abstract

A large number of randomized controlled trials (RCTs) on psychological interventions for adult posttraumatic stress disorder (PTSD) have been published. We aimed at providing a comprehensive quantitative summary covering short and long-term efficacy, acceptability and trial quality. We conducted systematic searches in bibliographical databases to identify RCTs examining the efficacy (standardized mean differences in PTSD severity, SMDs) and acceptability (relative risk of all-cause dropout, RR) of trauma-focused cognitive behaviour therapy (TF-CBT), Eye Movement Desensitization and Reprocessing (EMDR), other trauma-focused psychological interventions (other-TF-PIs) and non-trauma-focused psychological interventions (non-TF-PIs) compared to each other or to passive or active control conditions. Hundred-fifty-seven RCTs met inclusion criteria comprising 11,565, 4,830 and 3,338 patients at post-treatment assessment, ≤ 5 months follow-up and > 5 months follow-up, respectively. TF-CBT was by fore the most frequently examined intervention. We performed random effects network meta-analyses (efficacy) and pairwise meta-analyses (acceptability). All therapies produced large effects compared to passive control conditions (SMDs ≥ 0.80) at post-treatment. Compared to active control conditions, TF-CBT and EMDR yielded medium treatment effects (SMDs ≥ 0.50 < 0.80), and other-TF-PIs and non-TF-PIs yielded small treatment effects (SMDs ≥ 0.20 < 0.50). Interventions did not differ in their short-term efficacy, yet TF-CBT was more effective than non-TF-PIs (SMD = 0.17) and other-TF-PIs (SMD = 0.30). Results remained robust in sensitivity and outlier-adjusted analyses. Similar results were found for long-term efficacy. Interventions also did not differ in terms of their acceptability, except for TF-CBT being associated with a slightly increased risk of dropout compared to non-TF-PIs (RR=1.36; 95% CI: 1.08-1.70). In sum, interventions with and without trauma focus appear effective and acceptable in the treatment of adult PTSD. TF-CBT is by far the most well-researched intervention and yields the highest efficacy. However, TF-CBT appears somewhat less acceptable than non-TF-PIs.

## Introduction

Post-traumatic stress disorder (PTSD) is a common condition that often follows a chronic course if left untreated and puts a substantial burden on affected individuals and society at large^1–3^. Recent wars, pandemics and natural disasters illustrate the high prevalence of trauma exposure in the general population^4–7^. Since a substantial minority of individuals will develop PTSD in the aftermath of trauma exposure, its effective treatment remains a national and global health priority. Psychological interventions are recommended as the first-line treatment for PTSD in leading treatment guidelines^8^. Over the past four decades, a range of psychological interventions for PTSD in adulthood were developed and their efficacy and acceptability have been examined in numerous randomized controlled trials (RCTs). Several recent pairwise meta-analyses^9–11^ and network meta-analyses^12–15^ (NMAs) found evidence of large short-term and long-term efficacy of psychological interventions relative to passive control conditions, moderate short-term and long-term efficacy compared to active control conditions and adequate acceptability of psychological interventions with similar or slightly higher dropout rates relative to control conditions. Trauma focused cognitive behaviour therapy (TF-CBT) is the most well-researched psychological intervention for PTSD, followed by Eye Movement Desensitization and Reprocessing (EMDR). However, the evidence base for other psychological interventions with or without trauma focus (other-TF-PIs & non-TF-PIs) is expanding, and the debate on whether a trauma focus is necessary for high efficacy in PTSD treatment continues. Advocates of non-TF-PIs have postulated that these interventions may be equally effective and yet more acceptable than trauma-focused psychological interventions because patients do not need to get exposed to distressing traumatic memories^16^.

A comprehensive NMA may inform about the relative efficacy and acceptability of trauma-focused psychological interventions relative to non-trauma-focused psychological interventions. This method allows integrating data from direct and indirect treatment comparisons and provides estimates of the relative effect of any given pair of interventions within the network of included interventions. That is, NMAs include comparisons between pairs of interventions that have never been evaluated within an individual RCT. NMAs may yield more precise estimates than the direct estimates produced in pairwise meta-analyses^17^. While three NMAs have recently been published in the field of PTSD, the present NMA adds to these in four main regards. First and foremost, a comprehensive NMA is lacking. Two recently published NMAs focused on one specific psychological intervention rather than all psychological interventions ^13,14^ and the other NMA^15^ is based on a search performed in January 2018 with 90 included RCTs compared to 157 included RCTs in the present NMA. Second, no NMA has adequately investigated the impact of trial quality, which may bias quantitative summaries of clinical research^18^. While trial quality was assessed in all previous NMAs and qualitative judgments were reported in terms of the general risk of bias and the general level of confidence in study conclusions, the potential impact of risk of bias on treatment effects was not examined (e.g., analyses including only trials with a low risk of bias). Third, the only recent NMA on the efficacy of all psychological interventions^15^ did not examine the potential impact of varying treatment delivery formats on outcome estimates (e.g., group TF-CBT and individual TF-CBT were merged in their analyses). Fourth, a comprehensive NMA of long-term efficacy is lacking. Mavranezouli et al.^15^ included follow-up data up to 4 months post-treatment. However, various studies reported longer follow-up assessments such as twelve months^19^. To this end, we conducted a comprehensive NMA of RCTs on the short-term and long-term efficacy and acceptability of psychological interventions with and without trauma focus for adult PTSD while statistically controlling for trial quality and treatment delivery format.

## Method

The aims and methods of the present NMA were pre-registered with the PROSPERO database (CRD42020206290). This NMA adheres to the Preferred Reporting Items for Systematic Reviews and Meta-analyses (PRISMA) guidelines for NMAs^20^. See Appendix A (in the supplementary information) for the PRISMA checklist. The systematic literature search, data extractions, and quality ratings were carried out independently by at least two authors. We defined the main structured research question outlining the Population, Intervention, Comparison, Outcome, and Study design (PICOS) as follows: in adult individuals with diagnosed PTSD (P), how do psychological interventions with or without trauma focus (I), compared to control conditions (C), perform in terms of their efficacy and acceptability in reducing adult PTSD (O) in randomized controlled trials (S)?

### Identification and selection of studies

For the timespan from inception to September 2020, we relied on our previous literature search^10^. For the timespan thereafter and up to April 21^st^ 2022, we conducted a new systematic literature search by applying the same search strategy. THH and MJ carried out the systematic literature search. Discrepancies were discussed amongst THH, MJ and NM. We searched MEDLINE, PsycINFO, Web of Science and PTSDpubs with multi-field searches (titles, abstracts and key words) utilizing various Thesaurus and MeSH search terms for PTSD as well as treatment (see Appendix B for the search string). No restrictions on language or format of publication (e.g., dissertations or book chapters) were applied. We also inspected 176 related qualitative and quantitative reviews for further eligible trials. See Appendix C for their references. We further searched the US Veteran Affairs trial registry and the reference lists of included trials for further eligible trials.

### Identification and selection of studies

In line with our previous study^10^, we included trials that met all of the following inclusion criteria: 1) random group allocation, 2) at least one arm investigating the efficacy of a psychological intervention was compared to either a control condition (passive or active) or to another psychological intervention from a different intervention family (see below), 3) mean age of sample ≥ 18 years, 4) at least ten participants analysed per comparison group, 5) PTSD was the primary complaint and primary treatment target, 6) at least 70% of the total participants were diagnosed with PTSD at baseline by means of a (semi-)structured interview based on PTSD criteria reported in any iteration of the Diagnostic and Statistical Manual of Mental Disorders (DSM^21^) or the International Statistical Classification of Diseases (ICD^22^). RCTs were excluded if they included only patients with PTSD and comorbid substance use disorders^23^ or comorbid traumatic brain injury^24^. Other than that, comorbidities were allowed as long as inclusion criterion 5) was met. Attentional trainings^25^ and attentional bias modification trainings^26^ were not deemed to belong to the category of psychological interventions and excluded. In cases where all inclusion criteria were met but the reporting was insufficient to calculate effect sizes, we contacted the corresponding author of the given publication via e-mail. In case of no response, we sent a reminder e-mail one month later in an attempt to obtain the relevant data. If we did not receive a response, the respective study was not included.

### Quality assessment

Risk of bias was independently assessed by THH and AK by means of eight quality criteria reported by Cuijpers et al.^18^. These criteria were originally based on the Cochrane Collaboration criteria to assess the methodological validity of clinical research^27^ and criteria to assess the quality of evidence-based treatment delivery^28^. Criteria were scored dichotomously leading to a possible quality sum score ranging from 0 to 8. Trials met the respective quality criterion when: 1) 100% of the included participants were diagnosed with PTSD at baseline by means of a diagnostic interview, 2) a treatment manual was followed and sufficiently reported on, 3) therapists were trained to apply the particular manual, 4) the treatment integrity was formally checked (e.g., regular supervision or standardized quantitative ratings of recordings), 5) intention-to-treat (ITT) results were reported, 6) *N* ≥ 50 (i.e., *n*_*1*_ + *n*_*2*_ ≥ 50), 7) a valid randomization procedure was performed, and 8) PTSD outcomes were assessed by blinded assessors or by self-report measures. Multi-arm trials may receive different quality sum scores per comparison in the light of varying group sizes (see quality criterion 6). In case of insufficient or missing reporting, the respective trial was rated as not meeting the given quality criterion. Trials were categorized as high-quality (i.e., low risk of bias) when at least seven quality criteria were met^18^. Initial agreement on independent ratings was good (90.31%). Disagreements were discussed and solved amongst THH, AK and NM. See the quality assessment tool in Appendix D and the quality ratings per trial in Appendix E.

### Categorization of interventions and control conditions

We divided psychological interventions into the following four categories: TF-CBT, EMDR, other-TF-PIs (e.g., imagery rehearsal therapy^29^) and non-TF-PIs (e.g., present centered therapy^30^). While several different interventions belonged to the other-TF-PIs and non-TF-PIs categories, there were too few trials to analyse these separately. With respect to TF-CBT, various interventions that share the core denominator of being based on CBT principles and involving a focus on the memory of the trauma(s) and/or trauma meaning have been applied in RCTs for PTSD. In the present NMA, the term TF-CBT is therefore an umbrella term for interventions such as cognitive therapy^31^, prolonged exposure^32^ or narrative exposure therapy^33^. Control conditions were divided into active control conditions (e.g., supportive counseling or treatment-as-usual) and passive control conditions (e.g., wait-list control or minimal attention). See Appendix F for categorizations of interventions and control conditions.

### Categorization of assessment timepoints

The following three assessment timepoints were distinguished for the efficacy estimates: post-treatment (i.e., treatment endpoint), follow up 1 (FU1, ≤ five months after treatment termination) and follow-up 2 (FU2, > five months after treatment termination). In cases where several assessments fell into the FU1 category, the one closest to five months was chosen. In cases where several assessments fell into the FU2 category, the longest assessment was chosen. Since we were interested in the acceptability of psychological interventions rather than the acceptability of being part of a clinical trial (i.e., doing repeated assessments over time), we focused on all-cause dropout rates between randomization until treatment endpoint.

### Data extraction

THH, MJ and AK independently extracted data. Disagreements were discussed between THH, MJ, AK, and NM. We extracted characteristics of the study (e.g., country of conduct, completer vs intention-to-treat analyses), participants (e.g., mean age, percentage females), interventions and control conditions (e.g., delivery format), and outcome data (sample size, mean & standard deviation per group at post-treatment, FU1 and FU2; all-cause dropout pre to post-treatment). When a trial included two psychological interventions from the same family, the primary intervention as indicated by the authors was chosen. For example, cognitive processing therapy rather than prolonged exposure therapy was chosen for Resick et al.^34^. When a trial included two variants of the same psychological intervention, we chose the bona fide variant. For example, in trials investigating the potential influence of session frequency on treatment efficacy, we included the psychological interventions delivered at ordinary frequency and excluded the extraordinary (i.e., intensely) delivered psychological interventions^30,35^. If available, we prioritized clinician-based PTSD outcome data such as the Clinician-Administered PTSD Scale (CAPS^36^) over self-report data. Similarly, intention-to-treat (ITT) data were prioritized over completer data.

### Outcomes

Relative short and long-term efficacy were estimated with standardized mean differences (Hedges’ g) of PTSD symptom severity^37^. Acceptability was estimated via relative risk (RR) of all-cause dropout.

### Statistical analysis

Data processing and statistical analyses were performed in R (version 3.6.1)^38^ utilizing the packages dplyr^39^, Rcpp^40^, readxl^41^, netmeta^42^, and metafor^43^. All analyses were based on a statistical significance level of α = .05. We calculated random effects NMAs because substantial heterogeneity was to be expected due to the high diversity in interventions and high diversity in populations studied. The netmeta package uses a frequentist weighted least squares approach. Hedges’ g (standardized mean differences, SMDs) were calculated by subtracting the group mean of the second group from the group mean of the first group, dividing the difference by the pooled standard deviation and then multiplying the quotient with the sample size correction factor *J* = 1 – (3/(4*df* − 1)^37^. Following Cohen’s conventions, SMDs may be interpreted as small (0.20), medium (0.50), and large (0.80)^44^. Effect sizes were reported with their 95% confidence intervals. To check whether the transitivity assumption was met, we compared whether comparison dyads differed on important patient characteristics (e.g., mean age of sample) and/or trial characteristics (e.g., trial quality). Furthermore, we checked for inconsistency in the global network^20,42^ and locally (i.e., per comparison) with net heat plots^45^ and the net splitting method^46^. We calculated τ^2^ and I^2^ to estimate heterogeneity in outcomes^47^. The latter indicates the proportion of the total variance in outcomes attributable to τ^2^ and may be interpreted as low (25%), moderate (50%), and high (75%)^48^. Furthermore, we calculated *Q*_het_ as a measure of heterogeneity within comparison designs and *Q*_inc_ as measure of heterogeneity between comparison designs^49^. To check for potential small-study effects and publication bias, we performed Egger’s test of asymmetry^50^ and inspected funnel plots. We defined outliers as observations that were at least 3.3 standard deviations above or below the pooled SMDs^51^. When we detected a minimum of one outlier, we conducted outlier-adjusted analyses. We further performed two sensitivity analyses for the NMAs of efficacy: a) analyses including only trials with low risk of bias (i.e., high-quality trials, see above); b) analyses including only trials delivering psychological interventions individually and face-to-face (i.e., excluding trials with other delivery formats such as group therapy or technology-based delivery which mainly belonged to TF-CBT potentially violating the transitivity assumption). We also calculated the surface under the cumulative ranking (SUCRA) for the data on short and long-term efficacy per assessment timepoint for the main as well as the sensitivity analyses to rank interventions by efficacy.

Estimating the relative acceptability was impeded by the fact that various studies reported zero pre to post-treatment dropout for all groups, which may bias results in (network) meta-analyses^52,53^. Consequently, we chose to calculate relative risk with pairwise meta-analyses because the calculation of relative risk excludes all comparisons with absolute risks of zero for both groups. Pairwise meta-analyses were only conducted when the evidence base for a given comparison was sufficiently large (i.e., k ≥ 4)^10^. Random effects pairwise meta-analyses were conducted because we expected large heterogeneity in outcomes due to large variability in populations and interventions studied. Relative risk is calculated by dividing the absolute risk (i.e., proportion of all-cause dropout) in the first comparison group by the absolute risk in the second comparison group. A relative risk of 1 indicates an equal risk among the two groups, a relative risk of below 1 indicates a lower risk in the first comparison group compared to the second comparison group and vice versa. We calculated 95% confidence intervals of relative risk around the I^2^-statistics with the non-central chi-squared-based approach^54^. We also calculated the pooled proportions of all-cause dropout per group in percent to provide an estimate of the general incidence of all-cause dropout per group. To summarize proportions, we conducted random effects pairwise meta-analyses of Freeman-Tukey double arcsine transformed prevalence proportions with the inverse variance method^55^. Furthermore, Agresti–Coull 95% confidence intervals were calculated^56^. Heterogeneity in outcomes was estimated by Q-statistics and the I²-statistics. As recommended^57^, we performed the Egger’s test of asymmetry to detect potential publication bias only when the evidence base was sufficiently large (i.e., k ≥ 10). In case Egger’s test indicated significant asymmetry, we used the trim-and-fill method and reported asymmetry-adjusted results^58^. As in the NMAs, we conducted and reported outlier-adjusted pairwise meta-analyses when at least one outlier was identified (see definition above).

## Results

### Study selection and characteristics

The examination of titles and abstracts led to the exclusion of 5,526 references. Of 206 full-texts screened, 21 reported on eligible RCTs. As such, a total of 157 RCTs were included (including 136 RCTs from our previous search^10^) comprising 11,565, 4,830 and 3,338 adult patients at post-treatment assessment, ≤ five months follow-up and > five months follow-up, respectively. Figure 1 depicts the corresponding PRISMA flow chart of the study selection process and search synthesis.

**Figure 1.**
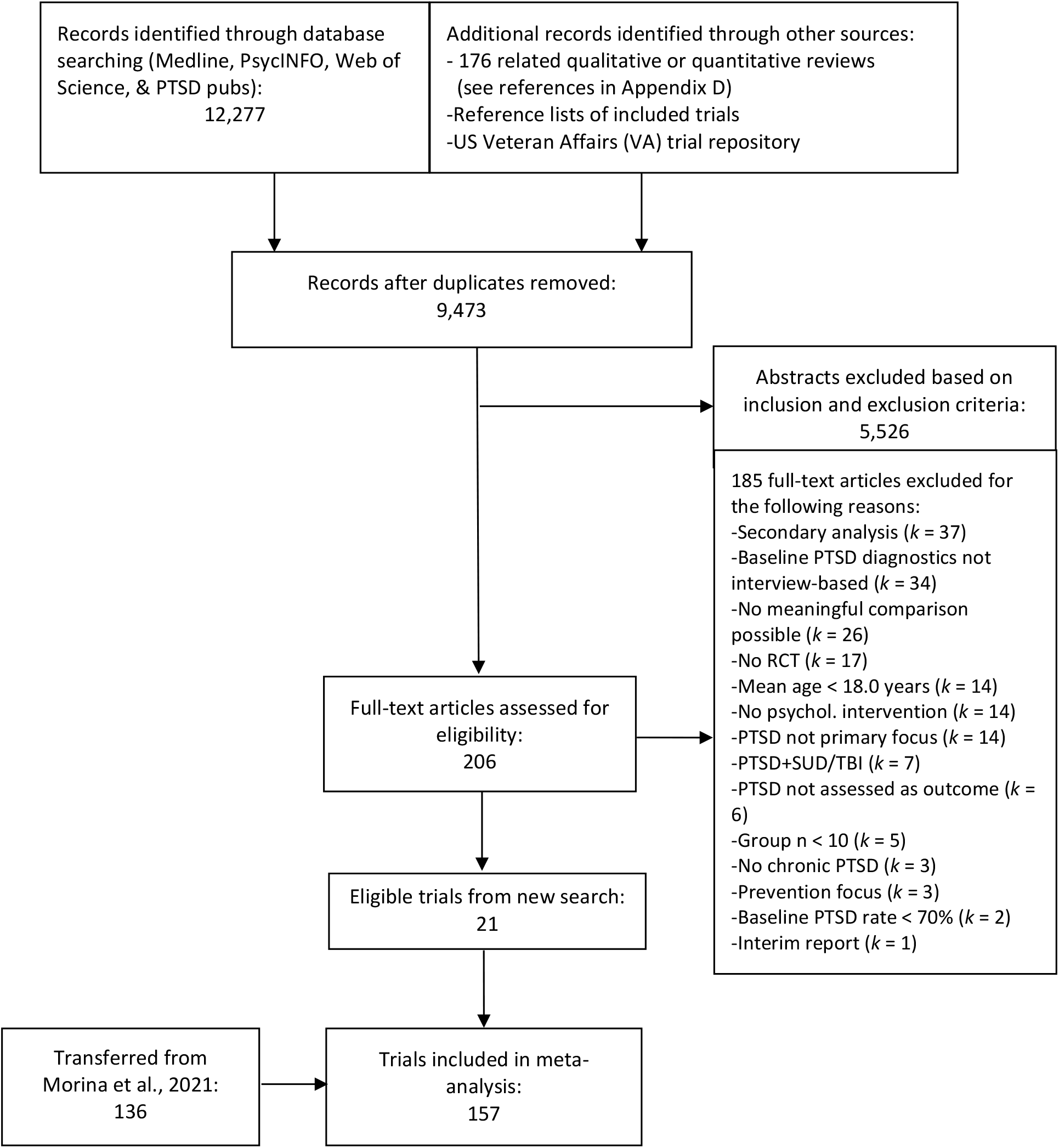
PRISMA flow chart of study selection

The vast majority of published trials (87%) were conducted in Western countries. Across all 157 RCTs (i.e., unweighted mean), participants were on average 39.80 years old and 59% of them were females. Thirty-eight trials (24%) included only female participants, fifteen trials (10%) only male participants and the remaining 66% of trials both sexes. A little less than half of the trials (41%) included participants with diverse trauma history and the remaining trials included participants with a specific trauma history, the most prevalent specific trauma history being combat (21% of trials) and sexual assault (17% of trials). Nine trials (6%) included exclusively participants who experienced their index trauma during childhood. Seventy-five trials (48%) reported on concurrent intake of psychotropic medication (i.e., whether participants were allowed to remain on a stable regimen of psychotropics during the treatment phase or not and how many participants were medicated during trial). Only eight trials (11% of those reporting on psychotropics) administered psychological interventions strictly as monotherapy (i.e., prohibiting participants to take any psychotropic medication during treatment^16,59–65^). Across trials reporting on concurrent intake of psychotropics, about 48% of participants were taking at least one psychotropic during psychological treatment.

On average, total treatment length across psychological interventions was 921 minutes (SD = 757 minutes) with an average of 11 treatment sessions (SD = 5 sessions). Most trials (82%) reported on the frequency of therapy sessions. Of these, three in four trials (75%) administered psychological interventions on a weekly session basis and the other quarter administered the psychological interventions with higher frequency such as twice weekly^66^. Trial quality was moderate on average (mean = 5.81 out of 8, SD = 1.46). Most trials included only participants meeting PTSD diagnostic criterial at baseline (90%), utilized a treatment manual (85%), involved manual-trained study therapists (80%), checked treatment integrity (77%), utilized a valid randomization procedure (62%) and assessed outcomes blinded to treatment allocation (86%). About half of the included trials reported results based on intention-to-treat analyses (53%, rather than completer analyses), involved a well-powered sample size (*N* ≥ 50, 48%) and utilized the gold standard PTSD measure, the CAPS (53%) as (one of the) outcome measure(s). About three in four trials (78%) delivered psychological interventions in individual face-to-face format. Other delivery formats were group therapy^67^, couple therapy^68^, internet/technology-based interventions either completely or mainly delivered remotely^69,70^ or face-to-face technology-aided interventions such as 3MDR (i.e., a virtual reality-assisted version of EMDR^71^). Results of six trials were included in the analyses of follow-up data only. One of these trials reported data on fewer than ten participants in the control condition at the post-treatment assessment^72^ and the other five trials did not conduct post-treatment assessments^61,73–76^. See Appendices F and G for a comprehensive overview of trial characteristics per included trial and a list of their references, respectively.

### Network graph and assumption checks

See Figure 2 for the network graph of the post-treatment assessment data and Appendices H and I for the network graphs for FU1 and FU2, respectively, and Appendix J for the number of trials per dyad for all NMAs including the sensitivity analyses. The vast majority of trials investigated the efficacy of TF-CBT. See Figure 3 for a bean plot on the distribution of participants’ mean age across comparison dyads and Appendix K for a bean plot on the distribution of the proportions of females in the total sample and Appendix L for a general comparison of participant and study characteristics across comparison dyads.

**Figure 2.**
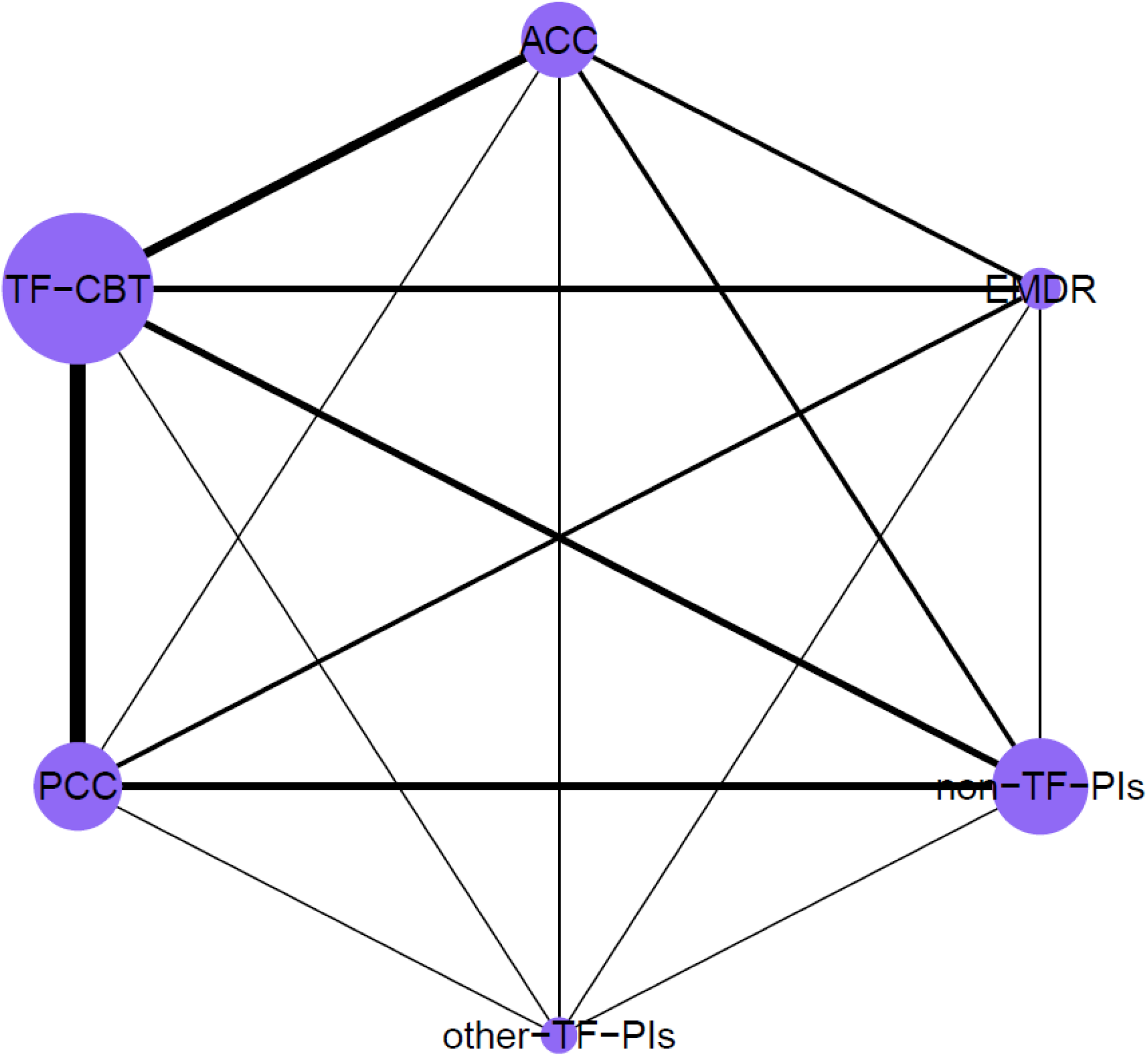
Network graph for the NMA on short-term efficacy Nodes sizes are proportional to the number of included participants per dyad and the thickness of lines is proportional to the number of direct comparisons. ACC – active control conditions (e.g., treatment-as-usual), EMDR – eye movement desensitization and reprocessing, non-TF-PIs – non-trauma-focused psychological interventions, other-TF-PIs – other trauma-focused psychological interventions (i.e., other than TF-CBT and EMDR), PCC – passive control conditions (e.g., waitlist), TF-CBT – trauma-focused cognitive behaviour therapy

**Figure 3.**
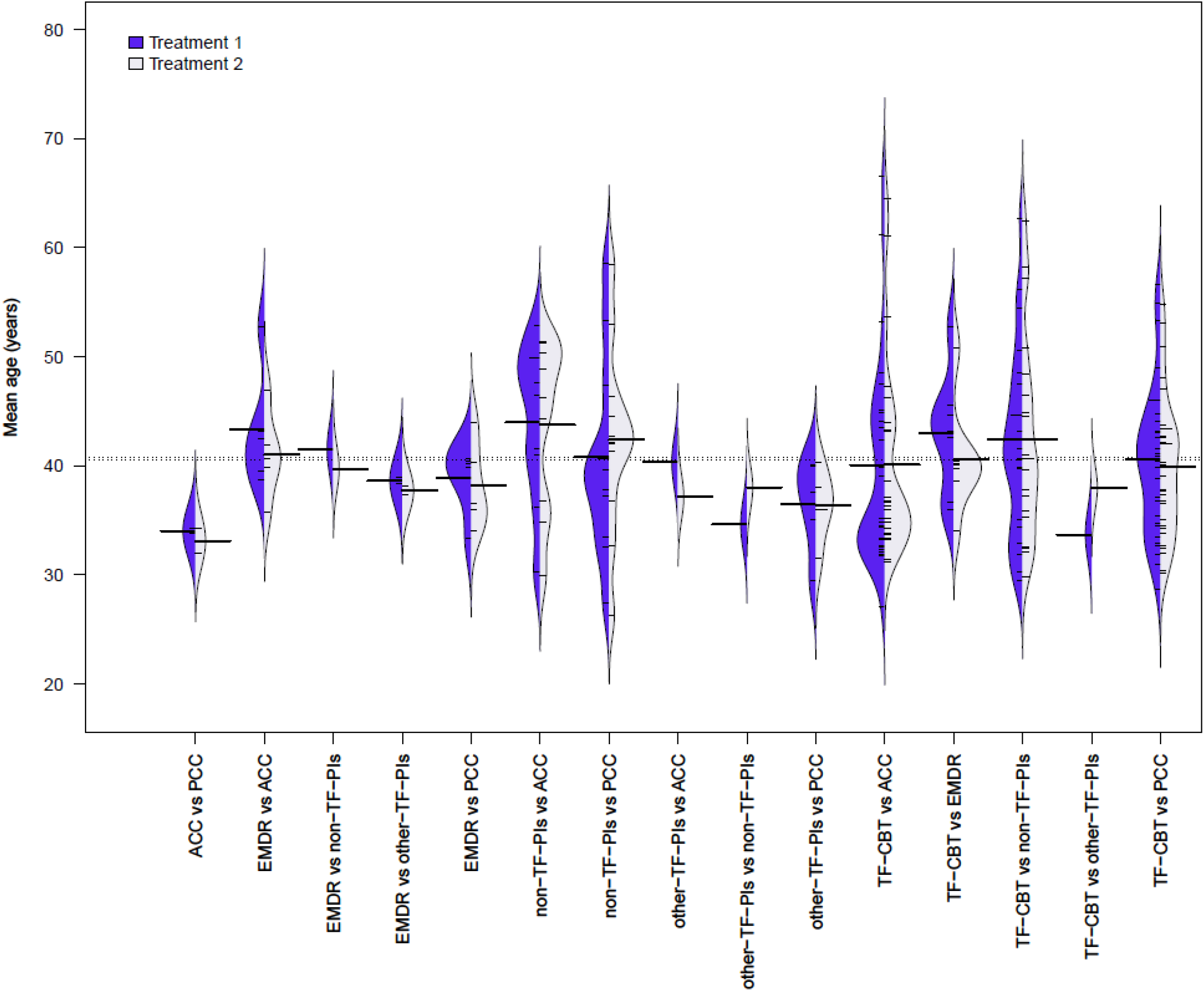
Bean plot of mean age across comparison dyads for the NMA on short-term efficacy ACC – active control conditions (e.g., treatment-as-usual), EMDR – eye movement desensitization and reprocessing, non-TF-PIs – non-trauma-focused psychological interventions, other-TF-PIs – other trauma-focused psychological interventions (i.e., other than TF-CBT and EMDR), PCC – passive control conditions (e.g., waitlist), TF-CBT – trauma-focused cognitive behaviour therapy

### Network meta-analysis on short-term efficacy

At post-treatment assessment, the random effects NMA yielded large effect sizes for all psychological interventions when compared to passive control conditions (i.e., SMDs ≥ 0.8, *p* < .001). Compared to active control conditions, TF-CBT and EMDR were moderately more effective in reducing PTSD (i.e., SMDs > 0.5, *p* < .001) and other-TF-PIs and non-TF-PIs were slightly more effective (i.e., SMDs between 0.2 and 0.5, *p* < .05). Psychological interventions did not differ significantly in their efficacy with two exceptions. TF-CBT was found more effective than other-TF-PIs (SMD = 0.30, *p* = .033) and non-TF-PIs (SMD = 0.17, *p* = .017). Heterogeneity and inconsistency were large and significant within but not between comparison groups (*τ*^2^ = 0.16, *I*^2^ = 69.10%; *Q*_total_ = 533.22, df = 165, *p* < .001; *Q*_*het*_ = 499.04, df = 141, *p* < .001; *Q*_*inc*_ = 34.14, df = 24, *p* = .082). See Table 1 for an overview of the results and Appendices M and N for forest plots of short-term efficacy with passive control conditions and non-TF-PIs as reference groups, respectively, as well as Appendix O for the funnel plot (including the result of the Egger’s test) of all comparisons that included a control condition and Appendix P for the netheat plot. The NMA yielded very similar results when outliers were removed and the heterogeneity remained high (see Appendix Q for an overview and Appendix R for a funnel plot).

**Table 1.** Comparative short-term efficacy of psychological interventions (PIs)

| *Reference group* | *PIs* | *k (N)* | *SMD* | *95% CI* | *p* | *I<sup>2</sup> (τ<sup>2</sup>)* |
| --- | --- | --- | --- | --- | --- | --- |
| **Main analysis (i.e., irrespective of trial quality and delivery format)** |  |  |  |  |  |  |
| relative to PCC | TF-CBT | 53 (2,844) | **1.14** | **1.01 – 1.26** | **< .001** | 69.10<br>***<br>(0.16) |
|  | EMDR | 7 (396) | **1.06** | **0.84 – 1.28** | **< .001** |  |
|  | other-TF-PIs | 6 (385) | **0.83** | **0.56 – 1.11** | **< .001** |  |
|  | non-TF-PIs | 18 (911) | **0.96** | **0.81 – 1.12** | **< .001** |  |
|  | ACC | 5 (154) | **0.53** | **0.36 – 0.70** | **< .001** |  |
| relative to ACC | TF-CBT | 31 (1,736) | **0.60** | **0.46 – 0.75** | **< .001** |  |
|  | EMDR | 11 (449) | **0.53** | **0.31 – 0.75** | **< .001** |  |
|  | other-TF-PIs | 1 (71) | **0.30** | **0.01 – 0.60** | **.043** |  |
|  | non-TF-PIs | 16 (1,026) | **0.43** | **0.27 – 0.60** | **< .001** |  |
| relative to EMDR | TF-CBT | 10 (359) | 0.07 | -0.14 – 0.28 | .495 |  |
|  | other-TF-PIs | 2 (224) | -0.23 | -0.54 – 0.08 | .153 |  |
|  | non-TF-PIs | 1 (46) | -0.10 | -0.33 – 0.14 | .412 |  |
| relative to other-TF-PIs | TF-CBT | 2 (204) | **0.30** | **0.02 – 0.58** | **.033** |  |
|  | non-TF-PIs | 2 (173) | 0.13 | -0.16 – 0.42 | .377 |  |
| relative to non-TF-PIs | TF-CBT | 25 (2,587) | **0.17** | **0.03 – 0.31** | **.017** |  |
| **Sensitivity analysis: high quality trials only** |  |  |  |  |  |  |
| relative to PCC | TF-CBT | 22 (1,535) | **1.02** | **0.83 – 1.22** | **< .001** | 75.80<br>***<br>(0.17) |
|  | EMDR | 2 (200) | **0.97** | **0.61 – 1.34** | **< .001** |  |
|  | other-TF-PIs | 2 (66) | **0.83** | **0.40 – 1.26** | **< .001** |  |
|  | non-TF-PIs | 4 (317) | **0.90** | **0.63 – 1.18** | **< .001** |  |
|  | ACC | 1 (52) | **0.51** | **0.22 – 0.80** | **< .001** |  |
| relative to ACC | TF-CBT | 13 (1,036) | **0.51** | **0.28 – 0.74** | **< .001** |  |
|  | EMDR | 3 (162) | **0.46** | **0.10 – 0.82** | **< .001** |  |
|  | other-TF-PIs | 0 (0) | 0.32 | 0.28 – 0.74 | .178 |  |
|  | non-TF-PIs | 4 (366) | **0.39** | **0.10 – 0.68** | **.008** |  |
| relative to EMDR | TF-CBT | 2 (128) | 0.05 | -0.30 – 0.40 | .774 |  |
|  | other-TF-PIs | 2 (224) | -0.14 | -0.59 – 0.30 | .525 |  |
|  | non-TF-PIs | 1 (46) | -0.07 | -0.46 – 0.32 | .726 |  |
| relative to other-TF-PIs | TF-CBT | 2 (204) | 0.19 | -0.22 – 0.62 | .362 |  |
|  | non-TF-PIs | 0 (0) | 0.08 | -0.39 – 0.54 | .752 |  |
| relative to non-TF-PIs | TF-CBT | 11 (1,390) | 0.12 | -0.11 – 0.35 | .313 |  |
| **Sensitivity analysis: trials with individual face-to-face delivery of PIs only** |  |  |  |  |  |  |
| relative to PCC | TF-CBT | 45 (2,444) | **1.19** | **1.04 – 1.34** | **< .001** | 71.80<br>***<br>(0.19) |
|  | EMDR | 6 (349) | **1.15** | **0.90 – 1.40** | **< .001** |  |
|  | other-TF-PIs | 4 (198) | **0.93** | **0.58 – 1.27** | **< .001** |  |
|  | non-TF-PIs | 11 (653) | **1.02** | **0.81 – 1.23** | **< .001** |  |
|  | ACC | 5 (154) | **0.64** | **0.42 – 0.86** | **< .001** |  |
| relative to ACC | TF-CBT | 27 (1,558) | **0.55** | **0.37 – 0.73** | **< .001** |  |
|  | EMDR | 11 (449) | **0.51** | **0.27 – 0.75** | **< .001** |  |
|  | other-TF-PIs | 0 (0) | 0.29 | -0.08 – 0.66 | .128 |  |
|  | non-TF-PIs | 4 (280) | **0.38** | **0.14 – 0.62** | **.002** |  |
| relative to EMDR | TF-CBT | 10 (359) | 0.04 | -0.19 – 0.27 | .743 |  |
|  | other-TF-PIs | 2 (224) | -0.22 | -0.59 – 0.15 | .241 |  |
|  | non-TF-PIs | 1 (46) | -0.13 | -0.41 – 0.15 | .365 |  |
| relative to other-TF-PIs | TF-CBT | 2 (204) | 0.26 | -0.08 – 0.60 | .135 |  |
|  | non-TF-PIs | 2 (173) | 0.09 | -0.27 – 0.46 | .618 |  |
| relative to non-TF-PIs | TF-CBT | 19 (1,637) | 0.17 | -0.02 – 0.36 | .086 |  |

Bold prints highlight significant differences. \*\*\* $p < .001$ , \*\* $p < .01$ , \* $p < .05$ , correspond to the respective Q-statistic.
ACC – active control conditions (e.g., treatment-as-usual), EMDR – eye movement desensitization and reprocessing, k – number of trials for the given comparison, N – number of participants for the given comparison, non-TF-PIs – non-trauma-focused psychological interventions, other-TF-PIs – other trauma-focused psychological interventions (i.e., other than TF-CBT and EMDR), PCC – passive control conditions (e.g., waitlist), PIs – psychological interventions, SMD – standardized mean differences (i.e., Hedges' $g$ ), TF-CBT – trauma-focused cognitive behaviour therapy

### Sensitivity analyses

At the post-treatment assessment, about one in three trials (38%, k = 57) were classified as high-quality with low risk of bias (i.e., meeting at least seven of the eight quality criteria) and about three in four trials (73%, k = 110) delivered the psychological intervention(s) individually face-to-face. In the sensitivity analyses in which we controlled for the potential impact of risk of bias and heterogeneity in treatment delivery formats, results remained by and large the same. See Table 1 for an overview of the results and Appendix Q for the outlier-adjusted results.

### Network meta-analysis on long-term efficacy

Sixty-five and 37 trials included data collected at FU1 and FU2 assessments, respectively. Follow-up assessments were conducted on average 3.18 months (*SD* = 1.56 months) and 8.14 months (*SD* = 2.78 months) after the end of treatment, respectively. At FU1 (i.e., ≤ five months after the end of treatment), TF-CBT (SMD = 0.92, 95% CI: 0.71-1.13), EMDR (SMD = 0.84, 95% CI: 0.50-1.18) and other-TF-PIs (SMD = 0.80, 95% CI: 0.12-1.10) produced large effect sizes compared to passive control conditions and non-TF-PIs produced moderate effect sizes (SMD = 0.69, 95% CI: 0.44-0.93). Compared to active control conditions at FU1, TF-CBT produced moderate effect sizes (SMD = 0.56, 95% CI: 0.38-0.74) whereas EMDR (SMD = 0.48, 95% CI: 0.14-0.82) and non-TF-PIs (SMD = 0.33, 95% CI: 0.12-0.54) produced small effect sizes. Other-TF-PIs, however, were not found more effective than active control conditions (SMD = 0.44, 95% CI: −0.20-1.08). Generally, psychological interventions did not differ in terms of their efficacy at FU1 except TF-CBT being more effective than non-TF-PIs (SMD = 0.23, 95% CI: 0.06-0.40). Heterogeneity and inconsistency were moderate to large and highly significant within as well as between comparison groups (*τ*^2^ = 0.12, *I*^2^ = 64.00%; *Q*_total_ = 177.74, df = 64, *p* < .001; *Q*_*het*_ = 146.93, df = 52, *p* < .001; *Q*_*inc*_ = 30.66, df = 12, *p* = .002). See Table 2 for an overview of results and Appendices S and T for forest plots of relative efficacy at FU1 with passive control conditions and non-TF-PIs as reference groups, respectively, as well as Appendix U for the funnel plot excluding head-to-head comparisons. Egger’s test was significant (see Appendix U). After removing outliers, however, Egger’s test was not significant anymore (see Appendix V). The netheat plot can be found in Appendix W. Results remained robust when outliers were removed and heterogeneity decreased somewhat but remained significant (see Appendix X).

**Table 2.** Comparative long-term efficacy of psychological interventions

| *Reference group* | *PIs* | *k (N)* | *SMD* | *95% CI* | *p* | *I<sup>2</sup> (τ<sup>2</sup>)* |
| --- | --- | --- | --- | --- | --- | --- |
| **FU1 (i.e., ≤ 5 months follow-up) - main analysis** |  |  |  |  |  |  |
| relative to PCC | TF-CBT | 14 (761) | **0.92** | **0.71 – 1.13** | **< .001** | 64.00<br>***<br>(0.12) |
|  | EMDR | 2 (145) | **0.84** | **0.50 – 1.18** | **< .001** |  |
|  | other-TF-PIs | 1 (42) | **0.80** | **0.18 – 1.42** | **.012** |  |
|  | non-TF-PIs | 4 (256) | **0.69** | **0.44 – 0.93** | **< .001** |  |
|  | ACC | 1 (52) | **0.36** | **0.10 – 0.62** | **.007** |  |
| relative to ACC | TF-CBT | 18 (872) | **0.56** | **0.38 – 0.74** | **< .001** |  |
|  | EMDR | 3 (116) | **0.48** | **0.14 – 0.82** | **.005** |  |
|  | other-TF-PIs | 0 (0) | 0.44 | -0.20 – 1.08 | .182 |  |
|  | non-TF-PIs | 7 (434) | **0.33** | **0.12 – 0.54** | **.002** |  |
| relative to EMDR | TF-CBT | 4 (99) | 0.08 | -0.25 – 0.40 | .638 |  |
|  | other-TF-PIs | 1 (129) | -0.05 | -0.65 – 0.55 | .881 |  |
|  | non-TF-PIs | 1 (46) | -0.16 | -0.50 – 0.19 | .375 |  |
| relative to other-TF-PIs | TF-CBT | 0 (0) | 0.12 | -0.50 – 0.75 | .670 |  |
|  | non-TF-PIs | 0 (0) | -0.11 | -0.75 – 0.53 | .738 |  |
| relative to non-TF-PIs | TF-CBT | 17 (1,878) | **0.23** | **0.06 – 0.40** | **.007** |  |
| **Sensitivity analysis: trials with individual face-to-face delivery of PIs only** |  |  |  |  |  |  |
| relative to PCC | TF-CBT | 12 (670) | **0.98** | **0.72 – 1.24** | **< .001** | 65.30<br>***<br>(0.14) |
|  | EMDR | 1 (98) | **0.96** | **0.56 – 1.36** | **< .001** |  |
|  | other-TF-PIs | 1 (42) | **0.87** | **0.20 – 1.54** | **.011** |  |
|  | non-TF-PIs | 1 (147) | **0.75** | **0.41 – 1.09** | **< .001** |  |
|  | ACC | 1 (52) | **0.48** | **0.15 – 0.81** | **.005** |  |
| relative to ACC | TF-CBT | 16 (727) | **0.50** | **0.27 – 0.73** | **< .001** |  |
|  | EMDR | 3 (116) | **0.48** | **0.10 – 0.87** | **.014** |  |
|  | other-TF-PIs | 0 (0) | 0.39 | -0.31 – 1.09 | .275 |  |
|  | non-TF-PIs | 1 (53) | 0.27 | -0.05 – 0.59 | .095 |  |
| relative to EMDR | TF-CBT | 4 (99) | 0.02 | -0.35 – 0.38 | .933 |  |
|  | other-TF-PIs | 1 (129) | -0.09 | -0.74 – 0.55 | .775 |  |
|  | non-TF-PIs | 1 (46) | -0.21 | -0.62 – 0.20 | .312 |  |
| relative to other-TF-PIs | TF-CBT | 0 (0) | 0.11 | -0.57 – 0.79 | .750 |  |
|  | non-TF-PIs | 0 (0) | -0.12 | -0.83 – 0.59 | .745 |  |
| relative to non-TF-PIs | TF-CBT | 11 (968) | 0.23 | -0.02 – 0.47 | .067 |  |
| **FU2 (i.e., > 5 months follow-up) - main analysis** |  |  |  |  |  |  |
| relative to PCC | TF-CBT | 5 (399) | **0.71** | **0.43 – 0.98** | **< .001** | 45.00<br>**<br>(0.04) |
|  | EMDR | 1 (102) | **0.42** | **0.01 – 0.83** | **.045** |  |
|  | other-TF-PIs | 0 (0) | **0.62** | **0.18 – 1.05** | **.005** |  |
|  | non-TF-PIs | 0 (0) | **0.51** | **0.19 – 0.82** | **.002** |  |
|  | ACC | 1 (51) | -0.01 | -0.33 – 0.31 | .963 |  |
| relative to ACC | TF-CBT | 15 (677) | **0.71** | **0.53 – 0.90** | **< .001** |  |
|  | EMDR | 0 (0) | **0.43** | **0.02 – 0.83** | **.038** |  |
|  | other-TF-PIs | 0 (0) | **0.62** | **0.23 – 1.02** | **.002** |  |
|  | non-TF-PIs | 1 (102) | **0.52** | **0.29 – 0.75** | **< .001** |  |
| relative to EMDR | TF-CBT | 2 (148) | 0.29 | -0.07 – 0.65 | .115 |  |
|  | other-TF-PIs | 1 (106) | 0.20 | -0.21 – 0.60 | .337 |  |
|  | non-TF-PIs | 0 (0) | 0.09 | -0.30 – 0.48 | .647 |  |
| relative to other-TF-PIs | TF-CBT | 2 (204) | 0.09 | -0.26 – 0.44 | .612 |  |
|  | non-TF-PIs | 0 (0) | -0.11 | -0.49 – 0.28 | .586 |  |
| relative to non-TF-PIs | TF-CBT | 13 (1,549) | **0.20** | **0.04 – 0.35** | **.012** |  |

Bold prints highlight significant differences. \*\*\* $p < .001$ , \*\* $p < .01$ , \* $p < .05$ , correspond to the respective Q-statistic.
ACC – active control conditions (e.g., treatment-as-usual), EMDR – eye movement desensitization and reprocessing, k – number of trials for the given comparison, N – number of participants for the given comparison, non-TF-PIs – non-trauma-focused psychological interventions, other-TF-PIs – other trauma-focused psychological interventions (i.e., other than TF-CBT and EMDR), PCC – passive control conditions (e.g., waitlist), PIs – psychological interventions, SMD – standardized mean differences (i.e., Hedges' g), TF-CBT – trauma-focused cognitive behaviour therapy

At FU2 (i.e., > five months after the end of treatment), TF-CBT (SMD = 0.71, 95% CI: 0.43-0.98), other-TF-PIs (SMD = 0.62, 95% CI: 0.18-1.05) and non-TF-PIs (SMD = 0.51, 95% CI: 0.19-0.82) were moderately more effective than passive control conditions. However, SMDs for the latter two categories of psychological interventions were estimated via indirect comparisons only. EMDR was found slighlty more effective than passive control conditions, and with only one available direct comparison (SMD = 0.42, 95% CI: 0.01-0.89). Results for the comparison to active control conditions were similar to those for the comparison to passive control conditions, with TF-CBT robustly yielding the largest SMDs. There was no evidence of differences in efficacy between PIs. Yet, TF-CBT appeared somewhat more effective than non-TF-PIs (SMD = 0.20, 95% CI: 0.04-0.35). Noteworthy, trials examining the long-term efficacy of other-TF-PIs and EMDR were scarce and trials on non-TF-PIs were limited to the comparison with TF-CBT. Heterogeneity and inconsistency were moderate and statistically significant between, but not within, comparison groups (*τ*^2^ = 0.04, *I*^2^ = 45.00%; *Q*_total_ = 61.84, *df* = 34, *p* = .002; *Q*_*het*_ = 49.31, *df* = 28, *p* = .008; *Q*_*inc*_ = 12.51, *df* = 6, *p* = .052). See Table 2 for an overview of the results and Appendices Y and Z for forest plots of the relative efficacy at FU1 with passive control conditions and non-TF-PIs as reference groups, respectively, as well as Appendix AA and AB for the funnel plot and outlier-corrected funnel plot, respectively. The Egger’s test was non-significant in both the main analysis and the outlier-adjusted analysis (see Appendix AA & Appendix AB, respectively). See Appendix AC for the netheat plot. Results remained robust when outliers were removed and heterogeneity decreased somewhat but remained significant (see Appendix X).

### Sensitivity analyses

At FU1, 39% of trials (k = 26) were classified as high-quality and 69% (k = 46) delivered the PI(s) individually face-to-face. At FU2, 53% of trials (k = 20) were classified as high-quality and 84% (k = 32) delivered the psychological intervention(s) individually face-to-face. Due to lacking trials and thus statistical power, the only feasible sensitivity analysis concerned the FU1 data for which it was possible to conduct a sensitivity analysis of trials that delivered psychological interventions individually face-to-face. Results remained were very similar to the main analysis (see Table 2) also when one outlier was removed (see Appendix X).

### Ranking of efficacy

TF-CBT was robustly the best ranking psychological intervention category across timepoints in both main analyses and sensitivity analyses. EMDR was robustly the second-best ranking psychological intervention category apart from the FU2 for which other-TF-PIs were the second-best ranking psychological intervention category. Notably, trials assessing PTSD outcomes beyond five months after the end of treatment (i.e., FU2) remain generally scarce for all psychological intervention categories apart from TF-CBT and results are therefore to be regarded and interpreted with due caution (see Table 3 for all SUCRA results).

**Table 3.** Efficacy rankings^a^ of psychotherapies and control conditions

|  | SMD<sub>post</sub> | SMD<sub>post-HQ</sub> | SMD<sub>post-F2F</sub> | SMD<sub>FU1</sub> | SMD<sub>FU1-F2F</sub> | SMD<sub>FU2</sub> |
| --- | --- | --- | --- | --- | --- | --- |
| TF-CBT | 0.94 | 0.95 | 0.96 | 0.87 | 0.88 | 0.92 |
| EMDR | 0.81 | 0.79 | 0.74 | 0.73 | 0.72 | 0.54 |
| other-TF-PIs | 0.44 | 0.46 | 0.45 | 0.69 | 0.68 | 0.75 |
| non-TF-PIs | 0.60 | 0.60 | 0.66 | 0.50 | 0.50 | 0.59 |
| ACC | 0.20 | 0.20 | 0.20 | 0.21 | 0.22 | 0.11 |
| PCC | 0.00 | 0.00 | 0.00 | 0.00 | 0.00 | 0.10 |

<sup>a</sup>Ranked by means of “surface under the cumulative ranking” (SUCRA) for standardized mean differences (SMD) at treatment endpoint (post), $\leq 5$ months follow up (FU1), and $> 5$ months follow-up (FU2). ACC – active control conditions (e.g., treatment-as-usual), EMDR – eye movement desensitization and reprocessing, F2F – sensitivity-analysis on trials with individual face-to-face delivery of psychotherapies only (i.e., adjusting for the potential influence of other delivery formats such as group or online therapy), HQ – sensitivity-analysis on high-quality trials only (i.e., adjusting for risk of bias through methodological limitations), non-TF-PIs – non-trauma-focused psychological interventions, other-TF-PIs – other trauma-focused psychological interventions (i.e., other than TF-CBT and EMDR), PCC – passive control conditions (e.g., waitlist), SMD – standardized mean differences (i.e., Hedges’ $g$ ), TF-CBT – trauma-focused cognitive behaviour therapy

### Pairwise meta-analyses of acceptability

Most psychological interventions did not differ significantly from the control conditions in terms of their acceptability as measured by the relative risk (RR) of all-cause dropout from pre to post-treatment assessment. TF-CBT (RR=1.53; 95% CI: 1.23-1.90) and other-TF-PIs (RR=1.35; 95% CI: 1.00-1.83) were found somewhat less acceptable compared to passive control conditions. None of the psychological interventions differed from active control conditions in terms of acceptability. Data for other-TF-PIs in comparison to active control conditions were too scarce to allow for meta-analytic review. The only comparisons amongst psychological interventions with a sufficient number of trials were TF-CBT with EMDR and TF-CBT with non-TF-PIs. While no evidence was found for a differential acceptability of TF-CBT compared to EMDR, TF-CBT was found to be somewhat less acceptable than non-TF-PIs (RR=1.36; 95% CI: 1.08-1.70). Outlier-adjusted analyses as well as trim-and-fill-adjusted analyses yielded similar results. Across the various comparisons, dropout rates for psychological interventions ranged from about one in eight (12.17% for EMDR) to about one in four participants (26.34% for other-TF-PIs) and for control conditions from about one in 13 (7.90% for passive control conditions) to about one in five participants (20.56% for active control conditions). See Table 4 for an overview of all results.

**Table 4.** Comparative acceptability of psychological interventions (PIs) and in comparison to control conditions

| *Reference Group (RG)* | *PIs* | *RR of DO* | *95% CI* | *k* | *p* | *I<sup>2</sup>* | *DO rate RG* | *95% CI* | *I<sup>2</sup>* | *k<sup>a</sup>* | *DO rate PIs* | *95% CI* | *I<sup>2</sup>* | *k<sup>a</sup>* |
| --- | --- | --- | --- | --- | --- | --- | --- | --- | --- | --- | --- | --- | --- | --- |
| PCC | TF-CBT | **1.53** | **1.23 – 1.90** | 45 | **< .001** | 14.90 | 7.90% | 5.52% - 10.58% | 57.30*** | 53 | 15.26% | 11.56% - 19.31% | 74.40*** | 53 |
|  | TF-CBT<sup>#</sup> | n.a. (Egger’s test non-significant) |  |  |  |  | 12.71% | 9.42% - 16.33% | 71.60*** | 69 | 23.80% | 18.58% - 29.40% | 83.60*** | 70 |
|  | EMDR | 1.40 | 0.63 – 3.11 | 6 | .328 | 36.20 | 11.76% | 3.80% - 22.62% | 74.90*** | 7 | 22.85% | 16.77% - 29.50% | 43.00 | 7 |
|  | other-TF-PIs | **1.35** | **1.00 – 1.83** | 6 | **.050** | 0.00 | 15.82% | 4.77% - 30.91% | 84.10*** | 6 | 26.34% | 10.38% - 46.01% | 87.50*** | 6 |
|  | non-TF-PIs | 1.09 | 0.73 – 1.62 | 15 | .650 | 9.60 | 8.65% | 3.47% - 15.37% | 73.10*** | 18 | 12.69% | 7.11% - 19.35% | 72.50*** | 18 |
|  | non-TF-PIs<sup>+</sup> | 1.27 | 0.84 – 1.92 | 14 | .230 | 0.00 | 6.98% | 2.84% - 12.33% | 64.10*** | 17 | n.a. (no outlier observed) |  |  |  |
| ACC | TF-CBT | 1.02 | 0.89 – 1.17 | 28 | .806 | 0.00 | 16.67% | 12.65% - 21.07% | 59.80*** | 31 | 15.62% | 11.07% - 20.71% | 71.30*** | 31 |
|  | TF-CBT<sup>#</sup> | n.a. (Egger’s test non-significant) |  |  |  |  | 23.23% | 18.00% - 28.86% | 73.60*** | 42 | n.a. (Egger’s test non-significant) |  |  |  |
|  | EMDR | 1.12 | 0.76 – 1.67 | 7 | .485 | 0.00 | 10.52% | 4.77% - 17.77% | 49.70* | 10 | 13.21% | 7.23% - 20.36% | 46.00 | 10 |
|  | other-TF-PIs | n.a. (*k* = 2) |  |  |  |  |  |  |  |  |  |  |  |  |
|  | non-TF-PIs | 1.03 | 0.80 – 1.33 | 15 | .817 | 11.80 | 20.56% | 14.06% - 27.85% | 73.50*** | 16 | 18.71% | 11.76% - 26.72% | 79.20*** | 16 |
| TF-CBT | EMDR | 1.00 | 0.61 – 1.65 | 6 | .999 | 0.00 | 10.83% | 3.08% - 21.48% | 71.60*** | 10 | 12.17% | 4.65% - 21.92% | 61.20** | 10 |
|  | EMDR<sup>+</sup> | n.a. (Egger’s test not performed since k = 6) |  |  |  |  | n.a. (Egger’s test non-significant) |  |  |  | 16.37% | 7.04% - 28.02% | 68.20*** | 12 |
|  | other-TF-PIs | n.a. (*k* = 2) |  |  |  |  |  |  |  |  |  |  |  |  |
|  | non-TF-PIs | **1.36** | **1.08 – 1.70** | 22 | **.011** | 45.20* | 24.63% | 17.09% - 32.98% | 89.70*** | 25 | 17.40% | 11.55% - 24.07 | 84.30*** | 25 |
| EMDR | other-TF-PIs | n.a. (*k* = 2) |  |  |  |  |  |  |  |  |  |  |  |  |
|  | non-TF-PIs | n.a. (*k* = 1) |  |  |  |  |  |  |  |  |  |  |  |  |
| other-TF-PIs | non-TF-PIs | n.a. (*k* = 2) |  |  |  |  |  |  |  |  |  |  |  |  |

Bold prints highlight significant differences. \*\*\* *p* < .001, \*\* *p* < .01, \* *p* < .05, correspond to the respective Q-statistic. <sup>a</sup>May differ from number of trials for calculated RR since trials with zero dropouts for both groups are omitted from RR calculations (i.e., zero absolute risk). <sup>#</sup>trim-and-fill-adjusted analysis. <sup>+</sup>outlier-adjusted analysis. ACC – active control conditions (e.g., treatment-as-usual), CI – confidence interval, DO rate – all-cause dropout rate pre-to-post-treatment assessment (i.e., acceptability of intervention), EMDR – eye movement desensitization and reprocessing, *k* – number of trials for the given comparison, n.a. – not applicable (reason), non-TF-PIs – non-trauma-focused psychological interventions, other-TF-PIs – other trauma-focused psychological interventions (i.e., other than TF-CBT and EMDR), PCC – passive control conditions (e.g., waitlist), PIs – psychological interventions, RG – reference group for given comparison, RR of DO – relative risk of all-cause dropout between comparison groups, TF-CBT – trauma-focused cognitive behaviour therapy

## Discussion

The aim of this study was to comprehensively summarize the comparative short-term and long-term efficacy and acceptability of PIs for adult PTSD while controlling for trial quality and treatment delivery format. The up-to-date systematic literature search yielded 157 eligible trials. The vast majority of these investigated the efficacy and acceptability of TF-CBT. All psychological interventions were effective in the short-term and in the long-term when compared to control conditions, with TF-CBT consistently yielding the most favourable results. TF-CBT was also the only psychological intervention that distinguished itself from other psychological interventions. TF-CBT was more effective than non-TF-PIs in most analyses and across timepoints. The pairwise meta-analyses of the relative acceptability yielded similar or slightly higher dropout rates for psychological interventions compared to passive control conditions and similar dropout rates relative to active control conditions. The only difference in acceptability between PIs was found for TF-CBT compared to non-TF-PIs, with the former being somewhat less acceptable.

### Comparison with previous research

Our results are in line with previous reviews. Most previous network and pairwise meta-analyses concluded that TF-CBT and EMDR are similarly effective in the treatment of adult PTSD^9,15,77,78^. However, one pairwise meta-analysis^79^ involving 11 trials concluded that EMDR outperforms TF-CBT. The data in our network meta-analysis, which are better suited to assess the relative efficacy of TF-CBT vs EMDR, indicate that these two treatments are similarly effective. As such, our findings support current treatment guidelines, most of which recommend these two interventions as first-line treatments^8^. Notably, the substantial gap in accumulated evidence particularly in terms of long-term outcomes precludes strong conclusions regarding the long-term efficacy of EMDR and some treatment guidelines do favor TF-CBT over EMDR in their recommendations^80^. More research is needed to draw firmer conclusions.

Furthermore, the present study is also in line with previous network and pairwise meta-analyses that did find higher efficacy of TF-CBT compared to non-TF-PIs in the short-run (i.e., at post-treatment assessment^9,15,77^). The relative long-term efficacy of TF-CBT and non-TF-PIs beyond four months follow-ups has not been analyzed yet in an NMA. A recent meta-analysis on the long-term efficacy of psychological interventions for adult PTSD concluded that TF-CBT and non-TF-PIs are similarly effective^11^. The present NMA is at odds with these findings by yielding superior long-term efficacy of TF-CBT over non-TF-PIs. Since our more recent search (i.e., performed in April 2022 vs. November 2019) resulted in a higher number of included trials, the present NMA had more power to find significant differences. Various trials on non-TF-PIs^81^ have been published after Weber et al’s^11^ search. Research on trauma-focused psychological interventions other than TF-CBT and EMDR remains scarce with very few studies evaluating long-term efficacy.

Psychological interventions for adult PTSD appear more effective than psychological interventions more generally, that is, across mental disorders. A recent umbrella review and meta-analysis^82^ summarized findings of 3,782 RCTs (N = 650,514 patients) on the efficacy of psychological interventions and pharmacotherapies for the treatment of various mental disorders and found a pooled SMD of 0.34 (95% CI: 0.26-0.42) for psychological interventions as monotherapy compared to placebo or treatment-as-usual, which is substantially below the SMDs found for psychological interventions in the present NMA (e.g., SMD of TF-CBT vs active control conditions at post-treatment assessment = 0.60). Similar results have been reported elsewhere. For instance, psychological interventions for PTSD produce larger treatment effects than psychological interventions for other anxiety-related disorders^83^.

Our results on acceptability are in line with a recent review of meta-analyses concluding that dropout rates tend to be higher in TF-CBT compared to other psychological interventions.^84^ Results are also mostly in line with a recent meta-analysis on the safety of psychological interventions summarizing 56 RCTs and concluding that psychological interventions were not associated with increased incidences of harms (i.e., deterioration, adverse events and serious adverse events) when compared to passive or active control conditions^85^. Rather, psychological interventions were safer than control conditions (both active and passive) in terms of deterioration risk. One difference to the present results was reported. While in the present work, we found TF-CBT to be somewhat less acceptable than non-TF-PIs, TF-CBT was found as safe as non-TF-PIs in terms of deterioration and adverse events and safer than non-TF-PIs in terms of severe adverse events in the other meta-analysis. This suggests that more patients prematurely drop out from TF-CBT than non-TF-PIs, yet patients receiving TF-CBT treatment do not appear to be at an increased risk of experiencing harms and a decreased risk of severe adverse events.

### Strengths and limitations

Our study represents the first NMA of adult PTSD that comprehensively reports on both short-term and long-term efficacy of psychological interventions while statistically controlling for the potential influence of trial quality and varying delivery formats on outcomes. Inclusion criteria were strict to ensure validity of analyses. Trials with high risk of chance findings (i.e., *N* < 20) were excluded. We further increased internal validity by only including trials with largely clinical samples (i.e., ≥ 70% PTSD rate) and adherence to gold standard diagnostic procedures (i.e., diagnostic interview to assess eligibility of participants). Despite the strict inclusion criteria, a substantial evidence base of 157 RCTs was included. Yet, the present NMA has certain limitations. The main limitation concerns the heterogeneity in some of our psychological interventions categories as well as the active control conditions category. Other-TF-PIs and non-TF-PIs are a conglomerate of different psychological interventions with the common denominator that they do not belong to TF-CBT or EMDR. Our decision to merge interventions in these rather heterogenous categories was based on the availability of trials. As more trials accumulate, more fine-grained approaches of categorizations with sufficient power will become feasible. The content of active control conditions is very context dependent and heterogenous^86^. Future research may bring forward more sophisticated ways of subcategorizing active control conditions into more homogenous subcategories. Another limitation concerns the operationalization of treatment acceptability. A comparison of all-cause dropout rates across trial arms is just a proxy of acceptability and other ways of operationalizing and comparing acceptability should be brought forward. However, this operationalization has been used widely in NMAs^14^, including NMAs of other mental disorders such as major depression^87,88^. The present study did not investigate other important clinical outcomes such as response and remission rates or the tolerability of interventions. While being important, these were not our focus. Lastly, our definition of high-quality trials (i.e., trials with low risk of bias) can be criticized. Other authors have used a cut-off score of eight (out of eight) fulfilled quality criteria^18^ and we have used at least seven. This choice was based on an attempt to balance out strict criteria for the definition of high-quality trials and power consideration for the sensitivity analyses. A threshold of at least seven (out of eight) has been used before for similar reasons.^89^ As more high-quality trials accumulate, more conservative sensitivity analyses (with sufficient power) will become feasible.

### Implication for clinical practice

The present results are reassuring for clinicians and patients because they illustrate that effective and acceptable psychological interventions for the treatment of adult PTSD exist. Most evidence and certainty exist for the short-term and long-term efficacy of TF-CBT in the treatment of adult PTSD. TF-CBT furthermore appears acceptable with only slightly higher all-cause discontinuation rates compared to passive control conditions and non-TF-PIs and no differences compared to active control conditions or EMDR. Other psychological interventions with and without trauma focus also appear to be effective and acceptable though these results are to be regarded with due caution in the light of considerably less available evidence. Though somewhat less acceptable, TF-CBT was found significantly more effective than non-TF-PIs immediately after treatment as well as across follow-ups. Some treatment guidelines do recommend treatment of adult PTSD with non-TF-PIs^90^. The evidence shows that non-TF-PIs do appear to be an effective (though somewhat less effective) alternative treatment, which, however, should rather be considered when treatment with TF-CBT or EMDR is not feasible, not effective or not in line with patient preferences.

### Implications for future research

The present NMA is a significant first step in examining the differential short-term and long-term efficacy and acceptability of psychological interventions with or without trauma focus for adult PTSD. However, once enough research has accumulated to warrant sufficient statistical power, future meta-analytic research needs to further differentiate interventions within the non-TF-PIs and other-TF-PIs categories. Furthermore, the evidence base of trials assessing long-term efficacy remains scarce for all psychological interventions except TF-CBT. To allow for firmer conclusions regarding the relative long-term efficacy, more trials with long-term assessments of efficacy are needed. Additionally, future research needs to investigate potential moderating effects of other delivery formats than individual face-to-face delivery on outcomes. Once more trials accumulate, sensitivity analyses of trials delivering psychological interventions in group format or technology-based will become feasible.

### Conclusions

The current evidence base suggests that bona fide psychological interventions are effective and acceptable in the treatment of PTSD relative to control conditions. However, TF-CBT is both the most well researched psychological intervention to date as well as the psychological intervention with most favourable results in terms of its short-term and long-term efficacy. TF-CBT was the only psychological intervention that distinguished itself from other psychological interventions with higher efficacy compared to non-TF-PIs. Results remained robust across outlier-adjusted analysis and sensitivity analyses. Psychological interventions were also generally acceptable with similar or only slightly increased risk of dropout compared to control conditions. Psychological interventions generally did not differ in their acceptability except for TF-CBT being somewhat less acceptable compared to non-TF-PIs. More research on psychological interventions other than TF-CBT is needed particularly evaluating long-term treatment efficacy to draw firmer conclusions in terms of overall and relative efficacy and acceptability.

## Data Availability

All data analysed are publically accessible. No new data were created. The datasets and the R scripts are available on request via e-mail to the first/corresponding author (THH).

## Data availability

All data analysed were extracted from published journal articles. No new data were created. The datasets and the R scripts are available on request via e-mail to the first author.

## Author contributions

THH and NM conceptualized the present study. THH and MJ independently performed the literature search and data extractions. THH, HH, MJ and JM performed (parts of) the statistical analyses. THH wrote the first draft of the manuscript. All authors contributed to and have approved the final manuscript.

