## Supplementary information for "Psychological interventions for adult PTSD: A network and pairwise meta-analysis of short and long-term efficacy, acceptability and trial quality"

##### Appendix A: PRISMA NMA checklist

##### Appendix B: Search string used for systematic literature search

##### Appendix C: References of related reviews and meta-analyses scrutinized as part of the systematic literature search

##### Appendix D: Risk of bias assessment – quality criteria

##### Appendix E: Risk of bias assessment – quality coding of included trials

##### Appendix F: Trial characteristics of included trials

##### Appendix G: References of trials included in the present network and pairwise meta-analysis

##### Appendix H: Network graph for FU1 (i.e., ≤ 5 months follow-up)

##### Appendix I: Network graph for FU2 (i.e., > 5 months follow-up)

##### Appendix J: Number of trials per dyad for all NMAs (incl. sensitivity analyses)

##### Appendix K: Bean plot for the distribution of participants’ sexes (i.e., % females)

##### Appendix L: Comparison of participant and trial characteristics across comparison dyads

##### Appendix M: Forest plot for NMA on short-term efficacy with passive control conditions as reference group

##### Appendix N: Forest plot for NMA on short-term efficacy with non-TF-PIs as reference group

##### Appendix O: Funnel plot for NMA on short-term efficacy

##### Appendix P: Netheat plot for NMA on short-term efficacy

##### Appendix Q: Outlier-corrected comparative short-term efficacy of psychological interventions (PIs).

##### Appendix R: Outlier-adjusted funnel plot for NMA on short-term efficacy

##### Appendix S: Forest plot for NMA at FU1 (i.e., ≤ 5 months follow-up) with passive control conditions as reference group

##### Appendix T: Forest plot for NMA at FU1 (i.e., ≤ 5 months follow-up) with non-TF-PIs as reference group

##### Appendix U: Funnel plot for NMA on efficacy at FU1 (i.e., ≤ 5 months follow-up)

Appendix V: Outlier-adjusted funnel plot for NMA on efficacy at FU1 (i.e., ≤ 5 months follow-up)

##### Appendix W: Netheat plot for NMA on efficacy at FU1 (i.e., ≤ 5 months follow up)

##### Appendix X: Outlier-corrected comparative long-term efficacy of psychological interventions (PIs)

##### Appendix Y: Forest plot for NMA at FU2 (i.e., > 5 months follow-up) with passive control conditions as reference group

##### Appendix Z: Forest plot for NMA at FU2 (i.e., > 5 months follow-up) with non-TF-PIs as reference group

##### Appendix AA: Funnel plot for NMA on efficacy at FU2 (i.e., > 5 months follow-up)

Appendix AB: Outlier-adjusted funnel plot for NMA on efficacy at FU2 (i.e., > 5 months follow-up)

##### Appendix AC: Netheat plot for NMA on efficacy at FU2 (i.e., > 5 months follow-up)

**Appendix A: PRISMA NMA Checklist**

| **Section/Topic** | **Item #** | **Checklist Item** | **Reported on Page #** |
| --- | --- | --- | --- |
| **TITLE** |  |  |  |
| Title | 1 | Identify the report as a network meta-analysis*.* | ***#***1 |
| **ABSTRACT** |  |  |  |
| Unstructured summary | 2 | Provide an unstructured summary. | #2 |
| **INTRODUCTION** |  |  |  |
| Rationale | 3 | Describe the rationale for the review in the context of what is already known*,* including mention of why a network meta-analysis has been conducted*.* | ***#***3-4 |
| Objectives | 4 | Provide an explicit statement of questions being addressed, with reference to participants, interventions, comparisons, outcomes, and study design (PICOS). | #4 |
| **METHODS** |  |  |  |
| Protocol and registration | 5 | Indicate whether a review protocol exists and if and where it can be accessed (PROSPERO, including registration number). | #4 |
| Eligibility criteria | 6 | Specify study characteristics (e.g., PICOS, length of follow-up) and report characteristics (e.g., years considered, language, publication status) used as criteria for eligibility, giving rationale. Clearly describe eligible treatments included in the treatment network, and note whether any have been clustered or merged into the same node (with justification). | ***#***4-7 |
| Information sources | 7 | Describe all information sources (e.g., databases with dates of coverage, contact with study authors to identify additional studies) in the search and date last searched. | #4-5 |
| Search | 8 | Present full electronic search strategy for at least one database, including any limits used, such that it could be repeated. | #4-5 & Appendix B |
| Study selection | 9 | State the process for selecting studies (i.e., screening, eligibility, included in systematic review, and, if applicable, included in the meta-analysis). | #5 |
| Data collection process | 10 | Describe method of data extraction from reports (independently, in duplicate) and any processes for obtaining and confirming data from investigators. | #4-5 |
| Data items | 11 | List and define all variables for which data were sought (e.g., PICOS, funding sources) and any assumptions and simplifications made. | #6-7 |
| Geometry of the network | S1 | Describe methods used to explore the geometry of the treatment network under study and potential biases related to it. This should include how the evidence base has been graphically summarized for presentation, and what characteristics were compiled and used to describe the evidence base to readers. | ***#***7 |
| Risk of bias within individual studies | 12 | Describe methods used for assessing risk of bias of individual studies (including specification of whether this was done at the study or outcome level), and how this information is to be used in any data synthesis. | #5-6 & #8 |
| Summary measures | 13 | State the principal summary measures (e.g., risk ratio, difference in means). | #8-9 |
| Planned methods of analysis | 14 | Describe the methods of handling data and combining results of studies for each network meta-analysis. This should include, but not be limited to:   - *Handling of multi-arm trials;* - *Selection of variance structure;* - *Assessment of model fit.* | #7-9 |
| Assessment of Inconsistency | S2 | Describe the statistical methods used to evaluate the agreement of direct and indirect evidence in the treatment network(s) studied. Describe efforts taken to address its presence when found. | #7-9 |
| Risk of bias across studies | 15 | Specify any assessment of risk of bias that may affect the cumulative evidence (e.g., publication bias, selective reporting within studies). | **#**8-9 |
| Additional analyses | 16 | Describe methods of additional analyses if done, indicating which were pre-specified. This may include, but not be limited to, the following:   - Sensitivity or subgroup analyses; - Meta-regression analyses; - *Alternative formulations of the treatment network.* | ***#***8-9 |
| **RESULTS** |  |  |  |
| Study selection | 17 | Give numbers of studies screened, assessed for eligibility, and included in the review, with reasons for exclusions at each stage, ideally with a flow diagram. | #9 |
| Presentation of network structure | S3 | Provide a network graph of the included studies to enable visualization of the geometry of the treatment network. | ***#***Fig. 2 & Appendices H & I |
| Summary of network geometry | S4 | Provide a brief overview of characteristics of the treatment network. This may include commentary on the abundance of trials and randomized patients for the different interventions and pairwise comparisons in the network, gaps of evidence in the treatment network, and potential biases reflected by the network structure. | ***#***11 |
| Study characteristics | 18 | For each study, present characteristics for which data were extracted (e.g., study size, PICOS, follow-up period) and provide the citations. | #9-10 |
| Risk of bias within studies | 19 | Present data on risk of bias of each study and, if available, any outcome level assessment. | Appendix E |
| Results of individual studies | 20 | For all outcomes considered (benefits or harms), present, for each study: 1) simple summary data for each intervention group, and 2) effect estimates and confidence intervals. | Appendices M, N, O, S & T |
| Synthesis of results | 21 | Present results of each meta-analysis done, including confidence/credible intervals. *In larger networks, authors may focus on comparisons versus a particular comparator (e.g. placebo or standard care), with full findings presented in an appendix. League tables and forest plots may be considered to summarize pairwise comparisons.* If additional summary measures were explored (such as treatment rankings), these should also be presented. | ***#***11-15; Tables 1-4 |
| Exploration for inconsistency | S5 | Describe results from investigations of inconsistency. This may include such information as measures of model fit to compare consistency and inconsistency models, *P* values from statistical tests, or summary of inconsistency estimates from different parts of the treatment network. | ***#***11-15 |
| Risk of bias across studies | 22 | Present results of any assessment of risk of bias across studies for the evidence base being studied. | #11-15 |
| Results of additional analyses | 23 | Give results of additional analyses, if done (e.g., sensitivity or subgroup analyses, meta-regression analyses*,* and so forth). | #11-15 |
| **DISCUSSION** |  |  |  |
| Summary of evidence | 24 | Summarize the main findings, including the strength of evidence for each main outcome; consider their relevance to key groups (e.g., healthcare providers, users, and policy-makers). | #15-17 |
| Limitations | 25 | Discuss limitations at study and outcome level (e.g., risk of bias), and at review level (e.g., incomplete retrieval of identified research, reporting bias). *Comment on the validity of the assumptions, such as transitivity and consistency. Comment on any concerns regarding network geometry (e.g., avoidance of certain comparisons).* | #17-18 |
| Conclusions | 26 | Provide a general interpretation of the results in the context of other evidence, and implications for future research. | #18-19 |
| **FUNDING** |  |  |  |
| Funding | 27 | Describe sources of funding for the systematic review and other support (e.g., supply of data); role of funders for the systematic review. This should also include information regarding whether funding has been received from manufacturers of treatments in the network and/or whether some of the authors are content experts with professional conflicts of interest that could affect use of treatments in the network. | ***#***20 |

PICOS = population, intervention, comparators, outcomes, study design.

From: Hutton B, Salanti G, Caldwell DM, Chaimani A, Schmid CH, Cameron C, Ioannidis JP, Straus S, Thorlund K, Jansen JP, Mulrow C, Catalá-López F, Gøtzsche PC, Dickersin K, Boutron I, Altman DG, Moher D. The PRISMA Extension Statement for Reporting of Systematic Reviews Incorporating Network Meta-analyses of Health Care Interventions: Checklist and Explanations. [Ann Intern Med. 2015;162(11):777-784.](http://annals.org/article.aspx?articleid=2299856)

**Appendix B:** **Search string used for systematic literature search**

| Databases | Search Terms |
| --- | --- |
| MEDLINE and PsycINFO | ( TI ( ptsd OR ptss OR post-traumatic stress OR posttraumatic stress OR post-traumatic syndrome OR posttraumatic syndrome) OR AB ( ptsd OR ptss OR post-traumatic stress OR posttraumatic stress OR post-traumatic syndrome OR posttraumatic syndrome ) OR SU ( ptsd OR ptss OR post-traumatic stress OR posttraumatic stress OR post-traumatic syndrome OR posttraumatic syndrome ) ) AND ( TI ( treatment* OR intervention* OR therap* OR psychotherap* OR exposure OR counse*ing OR trial* ) OR AB ( treatment* OR intervention* OR therap* OR psychotherap* OR exposure OR counse*ing OR trial* ) OR SU ( treatment* OR intervention* OR therap* OR psychotherap* OR exposure OR counse*ing OR trial* ) ) |
| PTSDpubs | (ptsd OR ptss OR post-traumatic stress OR posttraumatic stress OR post-traumatic syndrome OR posttraumatic syndrome) AND (treatment* OR intervention* OR therap* OR psychotherap* OR exposure OR counse*ing OR trial*) |
| Web of Science | ALL=( ptsd OR ptss OR post-traumatic stress OR posttraumatic stress OR post-traumatic syndrome OR posttraumatic syndrome ) AND ALL=( treatment* OR intervention* OR therap* OR psychotherap* OR exposure OR counse*ing OR trial* ) |
| Note that the search string contains APA thesaurus/MeSH search terms (i.e., “posttraumatic stress”, “treatment”, “intervention”, “psychotherapy”, “exposure” and “counseling” as well as additional terms (e.g., “post-traumatic stress”, “PTSD”, “trial”, “therapy”) in case a particular trial was not registered under these APA thesaurus/MeSH search terms. | |

**Appendix C:** **References of related reviews and meta-analyses scrutinized as part of the systematic literature search**

6.	Badesha K, Wilde S, Dawson DL. Mental health mobile app use to manage psychological difficulties: an umbrella review. *Ment Health Rev*. in press.

18.	Brochu R, Lefrançois J, Matte-Landry A, Grondin F, Bastien CH. Recension systématique sur l’efficacité des traitements des symptômes post-traumatiques nocturnes chez les victimes d’agression sexuelle. *Can Psychol*. in press.

26.	Cloitre M. Complex PTSD: Assessment and treatment. *Eur J Psychotraumatol*. 2021;12(sup1):1866423.

48.	Forman-Hoffman V, Middleton JC, Feltner C, et al. Psychological and pharmacological treatments for adults with posttraumatic stress disorder: a systematic review update. https://doi.org/10.23970/AHRQEPCCER207.

64.	Hollins Martin CJ, Reid K. A scoping review of therapies used to treat psychological trauma post perinatal bereavement. *J Reprod Infant Psychol*. in print.

77.	Klatte R, Strauss B, Flückiger C, Färber F, Rosendahl J. Defining and assessing adverse events and harmful effects in psychotherapy study protocols: A systematic review. *Psychotherapy*. in print.

100.	Madnick D, Spokas M. Reporting and inclusion of specific sociodemographic groups in the adult PTSD treatment outcome literature within the United States: A systematic review. *Clin Psychol: Sci Pract*. in print.

101.	Maglione MA, Chen C, Bialas A, et al. Combat and operational stress control interventions and PTSD: a systematic review and meta-analysis. *Mil Med*. 2022;187(7-8):e846-e855.

102.	Maglione MA, Chen C, Franco M, et al. Effect of patient characteristics on posttraumatic stress disorder treatment retention among veterans: A systematic review. *J Trauma Stress*. 2022;35(2):718-728.

103.	Maglione MA, Chen C, Franco M, et al. *Predictors of PTSD Treatment Retention and Response: A Systematic Review*. Santa Monica, California; 2022. www.rand.org/t/RR4191.

104.	Maher AR, Apaydin EA, Hilton L, et al. Sleep management in posttraumatic stress disorder: a systematic review and meta-analysis. *Sleep Med*. 2021;87:203-219.

105.	Malik N, Facer-Irwin E, Dickson H, Bird A, MacManus D. The effectiveness of trauma-focused interventions in prison settings: a systematic review and meta-analysis. *Trauma Violence Abuse*. in print.

125.	Peeters N, van Passel B, Krans J. The effectiveness of schema therapy for patients with anxiety disorders, OCD, or PTSD: A systematic review and research agenda. *Br J Clin Psychol*. in print.

126.	Pierce ZP, Black JM. The neurophysiology behind trauma-focused therapy modalities used to treat post-traumatic stress disorder across the life course: a systematic review. *Trauma Violence Abuse*. in print.

133.	Ramachandran HJ, Bin Mahmud MS, Rajendran P, Jiang Y, Cheng L, Wang W. Effectiveness of mindfulness‐based interventions on psychological well‐being, burnout and post‐traumatic stress disorder among nurses: A systematic review and meta‐analysis. *J Clin Nurs*. in print.

142.	Shorey S, Downe S, Chua JYX, Byrne SO, Fobelets M, Lalor JG. Effectiveness of psychological interventions to improve the mental well-being of parents who have experienced traumatic childbirth: a systematic review and meta-analysis. *Trauma Violence Abuse*. in print.

**Appendix D: Risk of bias assessment – quality criteria**

Quality criteria based on Cuijpers et al. (2010), sum score ranges from 0 to 8

| 1. All participants met diagnostic criteria for PTSD at baseline as assessed via a diagnostic interview based on any iteration of the DSM or ICD | 1. Positive   0. Negative / insufficient information |
| --- | --- |
| 2. Use of treatment manual   *(i.e., published, or specifically designed for the study;* ***all*** *psychological interventions of the RCT/included in the analyses were manual-based*  *🡪 insufficient: manual mentioned but without a reference/specification)* | 1. Positive   0. Negative / insufficient information |
| 3. Therapists were trained in applying the manual   *(i.e., specifically for the study or general training for respective manual)* | 1. Positive   0. Negative / insufficient information |
| 4. Treatment integrity was checked formally   *(i.e., by regular supervision and/or recordings and/or systematic screenings of protocol adherence with a standardized instrument)* | 1. Positive   0. Negative / insufficient information |
| 5. Data analyzed with intent-to-treat analysis   *(i.e., all persons who were randomized to the conditions initially were included in analyses; insufficient: if authors stated that both ITT and completer analyses were performed but only reported on completer results)* | 1. Positive   0. Negative / insufficient information |
| 6. Study had an adequate level of statistical power to find effects and included *N* ≥ 50 participants in the comparison groups (i.e., *n1+n2*)   *(note: may differ across assessment timepoints due to attrition (in completer analyses))* | 1. Positive   0. Negative / insufficient information |
| 7. Randomization by independent (3^rd^) party   *(e.g., independent person or computerized with valid randomization technique)* | 1. Positive   0. Negative / insufficient information |
| 8. Blind assessors for outcomes   *(i.e., PTSD outcomes were assessed either in a diagnostic interviewer or via self-reports; insufficient: non-blinded interviewers)* | 1. Positive   0. Negative / insufficient information |

| **Appendix E: Risk of bias assessment – quality coding of included trials** | | | | | | | | | |
| --- | --- | --- | --- | --- | --- | --- | --- | --- | --- |
| **Trial** | **Q1 -PTSD** | **Q2 -manual** | **Q3 -training** | **Q4 -integrity** | **Q5 -ITT** | **Q6 –**  **N > 50** | **Q7 -random-ization** | **Q8 -blinding** | **Q sum score** |
| **Acarturk et al. (2016)** | **1** | **1** | **1** | **1** | **1** | **1** | **1** | **1** | **8** |
| Akbarian et al. (2015) | 1 | 1 | 1 | 0 | 1 | 0 | 0 | 1 | 5 |
| **Andersen et al. (2021)** | **1** | **1** | **1** | **1** | **0** | **1** | **1** | **1** | **7** |
| **Asukai et al. (2010)** | **1** | **1** | **1** | **1** | **1** | **0** | **1** | **1** | **7** |
| Basoglu et al. (2005) | 1 | 1 | 0 | 1 | 0 | 1 | 1 | 1 | 6 |
| Basoglu et al. (2007) | 1 | 1 | 1 | 0 | 1 | 0 | 1 | 1 | 6 |
| Beck et al. (2009) | 1 | 1 | 1 | 1 | 0 | 0 | 0 | 1 | 5 |
| Bellehsen et al. (2021) | 1 | 1 | 0 | 0 | 1 | 0 | 1 | 1 | 5 |
| Belleville et al. (2018) | 1 | 1 | 0 | 1 | 1 | 0 | 1 | 1 | 6 |
| Bisson et al. (2020) | 1 | 1 | 1 | 1 | 0 | 0 | 1 | 1 | 6 |
| Blanchard et al. (2003) | 0 | 1 | 1 | 1 | 1 | 1 | 0 | 1 | 6 |
| **Bohus et al. (2013)** | **1** | **1** | **1** | **1** | **1** | **1** | **0** | **1** | **7** |
| Bormann et al. (2008) | 1 | 1 | 0 | 1 | 0 | 0 | 1 | 1 | 5 |
| Bormann et al. (2013) | 1 | 0 | 0 | 1 | 1 | 1 | 1 | 1 | 6 |
| **Boterhoven de Haan et al. (2020)** | **1** | **1** | **1** | **1** | **0** | **1** | **1** | **1** | **7** |
| Brom et al. (1989) | 1 | 0 | 1 | 1 | 0 | 0 | 0 | 1 | 4 |
| **Brom et al. (2017)** | **1** | **1** | **1** | **1** | **0** | **1** | **1** | **1** | **7** |
| Bryant et al. (2003) | 1 | 0 | 1 | 1 | 1 | 0 | 1 | 1 | 6 |
| Bryant et al. (2011) | 1 | 0 | 1 | 0 | 1 | 0 | 1 | 1 | 5 |
| Bryant et al. (2019) | 1 | 0 | 0 | 1 | 0 | 0 | 1 | 1 | 4 |
| **Buhmann et al. (2016)** | **1** | **0** | **1** | **1** | **1** | **1** | **1** | **1** | **7** |
| **Butollo et al. (2016)** | **1** | **1** | **1** | **1** | **1** | **1** | **0** | **1** | **7** |
| Capezzani et al. (2013) | 1 | 0 | 1 | 0 | 1 | 0 | 0 | 1 | 4 |
| Carletto et al. (2016) | 1 | 1 | 1 | 1 | 0 | 0 | 1 | 1 | 6 |
| Carlson et al. (1998) | 1 | 1 | 1 | 1 | 0 | 0 | 0 | 0 | 4 |
| **Carlsson et al. (2018)** | **1** | **1** | **0** | **1** | **1** | **1** | **1** | **1** | **7** |
| Carter et al. (2013) | 1 | 1 | 1 | 0 | 0 | 0 | 1 | 1 | 5 |
| Castillo et al. (2016) | 1 | 1 | 0 | 1 | 1 | 1 | 0 | 1 | 6 |
| Chard (2005) | 1 | 1 | 1 | 1 | 0 | 1 | 0 | 1 | 6 |
| Classen et al. (2020) | 0 | 0 | 0 | 0 | 0 | 0 | 0 | 1 | 1 |
| Cloitre et al. (2002) | 1 | 1 | 1 | 1 | 0 | 0 | 0 | 1 | 5 |
| **Cloitre et al. (2010)** | **1** | **1** | **1** | **1** | **1** | **1** | **1** | **1** | **8** |
| Cottraux et al. (2008) | 1 | 1 | 1 | 1 | 0 | 0 | 1 | 1 | 6 |
| Davis et al. (2020) | 1 | 1 | 0 | 1 | 0 | 1 | 1 | 1 | 6 |
| Devilly et al. (1998) | 1 | 1 | 1 | 0 | 0 | 0 | 0 | 0 | 3 |
| Devilly et al. (1999) | 1 | 1 | 1 | 1 | 0 | 0 | 0 | 0 | 4 |
| Dorrepaal et al. (2012) | 1 | 1 | 0 | 0 | 1 | 1 | 1 | 1 | 6 |
| **Duffy et al. (2007)** | **1** | **1** | **1** | **0** | **1** | **1** | **1** | **1** | **7** |
| Dunne et al. (2012) | 1 | 1 | 1 | 0 | 1 | 0 | 0 | 0 | 4 |
| Echeburua et al. (1997) | 1 | 0 | 1 | 0 | 1 | 0 | 0 | 0 | 3 |
| **Ehlers et al. (2003)** | **1** | **1** | **0** | **1** | **1** | **1** | **1** | **1** | **7** |
| Ehlers et al. (2005) | 1 | 1 | 1 | 1 | 1 | 0 | 0 | 1 | 6 |
| **Ehlers et al. (2014)** | **1** | **1** | **1** | **1** | **1** | **1** | **1** | **1** | **8** |
| **Engel et al. (2015)** | **1** | **1** | **1** | **1** | **0** | **1** | **1** | **1** | **7** |
| Ertl et al. (2011) | 1 | 1 | 1 | 1 | 0 | 1 | 0 | 1 | 6 |
| **Falsetti et al. (2008)** | **1** | **1** | **1** | **1** | **1** | **1** | **0** | **1** | **7** |
| Fecteau et al. (1999) | 1 | 1 | 1 | 1 | 0 | 0 | 1 | 1 | 6 |
| Foa et al. (1991) | 1 | 1 | 1 | 1 | 0 | 0 | 0 | 1 | 5 |
| Foa et al. (1999) | 1 | 1 | 1 | 1 | 0 | 0 | 0 | 1 | 5 |
| **Foa et al. (2005)** | **1** | **1** | **1** | **1** | **1** | **1** | **0** | **1** | **7** |
| **Foa et al. (2018)** | **1** | **1** | **1** | **1** | **1** | **1** | **1** | **1** | **8** |
| Fonzo et al. (2017) | 1 | 1 | 1 | 1 | 1 | 1 | 0 | 0 | 6 |
| **Forbes et al. (2012)** | **1** | **1** | **0** | **1** | **1** | **1** | **1** | **1** | **7** |
| Ford et al. (2011) | 0 | 1 | 1 | 1 | 1 | 1 | 1 | 0 | 6 |
| Ford et al. (2013) | 0 | 1 | 1 | 1 | 0 | 1 | 1 | 1 | 6 |
| **Galovski et al. (2012)** | **1** | **1** | **1** | **1** | **1** | **1** | **1** | **1** | **8** |
| **Gersons et al. (2000)** | **1** | **1** | **1** | **1** | **1** | **0** | **1** | **1** | **7** |
| Ghafoori et al. (2017) | 1 | 0 | 1 | 1 | 1 | 1 | 1 | 0 | 6 |
| Goldstein et al. (2017) | 0 | 0 | 1 | 1 | 1 | 0 | 0 | 1 | 4 |
| **Gray et al. (2019)** | **1** | **1** | **0** | **1** | **1** | **1** | **1** | **1** | **7** |
| Gray et al. (2021) | 1 | 1 | 1 | 1 | 0 | 0 | 1 | 1 | 6 |
| Heffner et al. (2016) | 0 | 1 | 0 | 0 | 0 | 0 | 0 | 0 | 1 |
| Hensel-Dittmann et al. (2011) | 1 | 1 | 1 | 1 | 0 | 0 | 1 | 1 | 6 |
| Hinton et al. (2011) | 1 | 1 | 1 | 0 | 1 | 0 | 0 | 1 | 5 |
| Hollifield et al. (2007) | 1 | 1 | 0 | 0 | 1 | 0 | 1 | 1 | 5 |
| **Ivarsson et al. (2014)** | **1** | **1** | **1** | **1** | **1** | **1** | **1** | **1** | **8** |
| **Jacob et al. (2014)** | **1** | **1** | **1** | **1** | **1** | **1** | **1** | **1** | **8** |
| Jalal et al. (2020) | 1 | 1 | 0 | 0 | 0 | 0 | 0 | 1 | 3 |
| Jensen et al. (1994) | 1 | 1 | 1 | 0 | 0 | 0 | 0 | 0 | 3 |
| Johnson et al. (2011) | 0 | 1 | 1 | 1 | 0 | 1 | 1 | 0 | 5 |
| Johnson et al. (2016) post-treatment | 0 | 1 | 1 | 1 | 0 | 1 | 1 | 1 | 6 |
| Johnson et al. (2016) FU2 | 0 | 1 | 1 | 1 | 0 | 0 | 1 | 1 | 5 |
| Johnson et al. (2020) | 1 | 1 | 1 | 1 | 0 | 1 | 0 | 1 | 6 |
| **Karatzias et al. (2011)** | **1** | **1** | **1** | **1** | **1** | **0** | **1** | **1** | **7** |
| Keane et al. (1989) | 1 | 1 | 0 | 0 | 0 | 0 | 0 | 0 | 2 |
| **Kearney et al. (2021)** | **1** | **1** | **1** | **1** | **1** | **1** | **1** | **1** | **8** |
| Kelly et al. (2021) post-treatment | 1 | 1 | 1 | 0 | 0 | 1 | 1 | 0 | 5 |
| Kelly et al. (2021) FU | 1 | 1 | 1 | 0 | 0 | 0 | 1 | 0 | 4 |
| Kent et al. (2011) | 1 | 0 | 1 | 0 | 1 | 0 | 0 | 1 | 4 |
| Krakow et al. (2000) | 0 | 1 | 0 | 0 | 0 | 1 | 0 | 1 | 3 |
| Krakow et al. (2001) | 0 | 1 | 0 | 0 | 0 | 1 | 1 | 1 | 4 |
| Krupnick et al. (2008) | 1 | 1 | 1 | 1 | 1 | 0 | 0 | 0 | 5 |
| Kubany et al. (2003) | 1 | 1 | 1 | 0 | 1 | 0 | 0 | 1 | 5 |
| **Kubany et al. (2004)** | **1** | **1** | **1** | **1** | **1** | **1** | **0** | **1** | **7** |
| Lang et al. (2019) | 1 | 1 | 1 | 0 | 0 | 0 | 1 | 1 | 5 |
| **Langkaas et al. (2017)** | **1** | **1** | **1** | **1** | **1** | **1** | **1** | **1** | **8** |
| **Latif et al. (2021)** | **1** | **1** | **0** | **1** | **1** | **1** | **1** | **1** | **7** |
| **Laugharne et al. (2016)** | **1** | **1** | **1** | **1** | **1** | **0** | **1** | **1** | **7** |
| Lee et al. (2002) | 1 | 1 | 0 | 1 | 0 | 0 | 1 | 0 | 4 |
| **Lehavot et al. (2021)** | **1** | **0** | **1** | **1** | **1** | **1** | **0** | **1** | **7** |
| **Lely et al. (2019)** | **1** | **1** | **1** | **1** | **1** | **0** | **1** | **1** | **7** |
| **Lewis et al. (2017)** | **1** | **1** | **1** | **1** | **1** | **0** | **1** | **1** | **7** |
| **Lindauer et al. (2005)** | **1** | **1** | **1** | **1** | **1** | **0** | **1** | **1** | **7** |
| Littleton et al. (2016) post-treatment | 1 | 1 | 1 | 1 | 0 | 1 | 1 | 0 | 6 |
| Littleton et al. (2016) FU | 1 | 1 | 1 | 1 | 0 | 0 | 1 | 0 | 5 |
| Litz et al. (2007) | 1 | 1 | 1 | 1 | 0 | 0 | 0 | 1 | 5 |
| Litz et al. (2021) | 1 | 1 | 0 | 1 | 0 | 1 | 1 | 1 | 6 |
| Maguen et al. (2017) | 0 | 0 | 1 | 1 | 0 | 0 | 1 | 1 | 4 |
| Marcus et al. (1997) | 1 | 1 | 1 | 0 | 0 | 1 | 1 | 1 | 6 |
| **Markowitz et al. (2015)** | **1** | **0** | **1** | **1** | **1** | **1** | **1** | **1** | **7** |
| Marks et al. (1998) | 1 | 0 | 1 | 1 | 0 | 0 | 0 | 1 | 4 |
| McDonagh et al. (2005) | 1 | 1 | 1 | 1 | 1 | 0 | 0 | 1 | 6 |
| McGuire Stanbury et al. (2020) | 1 | 1 | 1 | 1 | 0 | 0 | 0 | 1 | 5 |
| McLean et al. (2020) | 1 | 1 | 1 | 1 | 0 | 0 | 0 | 1 | 5 |
| Mitchell et al. (2014) | 0 | 1 | 1 | 0 | 1 | 0 | 1 | 1 | 5 |
| **Monson et al. (2006)** | **1** | **1** | **1** | **1** | **1** | **1** | **0** | **1** | **7** |
| **Monson et al. (2012)** | **1** | **1** | **1** | **1** | **1** | **0** | **1** | **1** | **7** |
| Morath et al. (2014) | 1 | 1 | 1 | 1 | 0 | 0 | 1 | 1 | 6 |
| Mueser et al. (2008) | 1 | 0 | 0 | 1 | 0 | 1 | 1 | 1 | 5 |
| **Nacasch et al. (2011)** | **1** | **1** | **1** | **1** | **1** | **0** | **1** | **1** | **7** |
| NCT00607815 | 1 | 1 | 0 | 0 | 1 | 1 | 0 | 1 | 5 |
| Neuner et al. (2004) | 1 | 1 | 1 | 1 | 0 | 0 | 1 | 1 | 6 |
| **Neuner et al. (2008)** | **1** | **1** | **1** | **1** | **1** | **1** | **0** | **1** | **7** |
| **Neuner et al. (2010)** | **1** | **1** | **1** | **1** | **1** | **0** | **1** | **1** | **7** |
| **Nidich et al. (2018)** | **1** | **1** | **0** | **1** | **1** | **1** | **1** | **1** | **7** |
| **Nijdam et al. (2012)** | **1** | **1** | **1** | **1** | **0** | **1** | **1** | **1** | **7** |
| Niles et al. (2012) | 1 | 1 | 1 | 1 | 0 | 0 | 1 | 0 | 5 |
| Orang et al. (2018) | 1 | 1 | 1 | 0 | 0 | 0 | 1 | 1 | 5 |
| **Pacella et al. (2012)** | **1** | **1** | **1** | **1** | **1** | **1** | **1** | **1** | **8** |
| Paunovic (2011) | 1 | 0 | 1 | 0 | 0 | 0 | 0 | 0 | 2 |
| Power et al. (2002) EMDR vs.WL | 1 | 1 | 1 | 1 | 0 | 1 | 1 | 0 | 6 |
| Power et al. (2002) other comparisons | 1 | 1 | 1 | 1 | 0 | 0 | 1 | 0 | 5 |
| Rauch et al. (2014) | 1 | 1 | 1 | 0 | 0 | 0 | 0 | 1 | 4 |
| Ready et al. (2018) | 1 | 0 | 1 | 1 | 0 | 1 | 0 | 1 | 5 |
| **Reger et al. (2016)** | **1** | **1** | **1** | **1** | **1** | **1** | **1** | **1** | **8** |
| Resick et al. (2002) | 1 | 1 | 1 | 1 | 1 | 1 | 0 | 0 | 6 |
| Resick et al. (2015) | 1 | 0 | 1 | 1 | 1 | 1 | 0 | 1 | 6 |
| **Robjant et al. (2019)** | **1** | **1** | **1** | **1** | **0** | **1** | **1** | **1** | **7** |
| Rothbaum et al. (2005) | 1 | 1 | 1 | 1 | 0 | 0 | 0 | 1 | 5 |
| **Sautter et al. (2015)** | **1** | **1** | **1** | **1** | **1** | **1** | **0** | **1** | **7** |
| **Schaal et al. (2009)** | **1** | **1** | **1** | **1** | **1** | **0** | **1** | **1** | **7** |
| Scheck et al. (1998) | 0 | 1 | 1 | 0 | 0 | 1 | 1 | 1 | 5 |
| Schnurr et al. (2003) | 1 | 0 | 1 | 1 | 0 | 1 | 1 | 1 | 6 |
| **Schnurr et al. (2007)** | **1** | **0** | **1** | **1** | **1** | **1** | **1** | **1** | **7** |
| Sloan et al. (2011) | 1 | 1 | 1 | 1 | 0 | 0 | 0 | 1 | 5 |
| Sloan et al. (2012) | 1 | 0 | 1 | 1 | 1 | 0 | 1 | 1 | 6 |
| **Sloan et al. (2018)** | **1** | **1** | **1** | **1** | **1** | **1** | **1** | **1** | **8** |
| **Spence et al. (2011)** | **1** | **1** | **1** | **1** | **1** | **0** | **1** | **1** | **7** |
| **Stenmark et al. (2013)** | **1** | **1** | **1** | **1** | **0** | **1** | **1** | **1** | **7** |
| **Suris et al. (2013)** | **1** | **1** | **1** | **1** | **0** | **1** | **1** | **1** | **7** |
| Taylor et al. (2003) | 1 | 1 | 1 | 1 | 1 | 0 | 0 | 1 | 6 |
| **ter Heide et al. (2016)** | **1** | **1** | **1** | **1** | **0** | **1** | **1** | **1** | **7** |
| Thorp et al. (2019) post-treatment | 1 | 1 | 1 | 1 | 0 | 1 | 0 | 1 | 6 |
| Thorp et al. (2019) FU | 1 | 1 | 1 | 1 | 0 | 0 | 0 | 1 | 5 |
| Tylee et al. (2017) | 1 | 1 | 0 | 1 | 1 | 0 | 0 | 1 | 5 |
| **van den Berg et al. (2015)** | **1** | **1** | **1** | **1** | **1** | **1** | **1** | **1** | **8** |
| **van der Kolk et al. (2007)** | **1** | **1** | **1** | **1** | **1** | **1** | **1** | **1** | **8** |
| van der Kolk et al. (2014) | 1 | 1 | 0 | 0 | 1 | 1 | 0 | 1 | 5 |
| **van Gelderen et al. (2020)** | **1** | **1** | **1** | **1** | **1** | **0** | **1** | **1** | **7** |
| Vaughan et al. (1994) | 0 | 1 | 1 | 0 | 0 | 0 | 0 | 1 | 3 |
| Vera et al. (2021) | 1 | 1 | 0 | 1 | 0 | 1 | 1 | 1 | 6 |
| Wagner et al. (2019) | 1 | 1 | 1 | 1 | 0 | 1 | 0 | 1 | 6 |
| Wahbeh et al. (2016) | 1 | 1 | 0 | 0 | 0 | 0 | 1 | 0 | 3 |
| Wells et al. (2012) | 1 | 1 | 0 | 1 | 1 | 0 | 1 | 1 | 6 |
| Wells et al. (2015) | 1 | 1 | 1 | 1 | 0 | 0 | 1 | 1 | 6 |
| **Yehuda et al. (2014)** | **1** | **1** | **1** | **1** | **1** | **1** | **0** | **1** | **7** |
| Yurtsever et al. (2018) | 1 | 1 | 1 | 0 | 0 | 0 | 1 | 1 | 5 |
| **Zang et al. (2013)** | **1** | **1** | **1** | **1** | **1** | **0** | **1** | **1** | **7** |
| **Zang et al. (2014)** | **1** | **1** | **1** | **1** | **1** | **0** | **1** | **1** | **7** |
| **Zemansti et al. (2022)** | **1** | **1** | **1** | **1** | **1** | **0** | **1** | **1** | **7** |
| Zlotnick et al. (1997) | 1 | 0 | 1 | 0 | 0 | 0 | 0 | 1 | 3 |
| Zoellner et al. (2017) | 1 | 1 | 1 | 1 | 1 | 0 | 1 | 0 | 6 |

Note: Bold font indicates high quality of trial (i.e., sum score ≥ 7 out of 8).

| Appendix F: Trial characteristics of included trials | | | | | | | | | | | |
| --- | --- | --- | --- | --- | --- | --- | --- | --- | --- | --- | --- |
| Publication, conditions/category,  (number & length of sessions) | N at post-treatment assessment | % fulfilling PTSD diagnosis at baseline | Outcome measure | Country | Mean age (SD or range) | Longest included follow-up assessment in months | Treatment format | Statistical analysis | % female | Type of trauma | Study quality at post-treatment assessment |
| Acarturk et al., 2016  EMDR/EMDR (7 sessions, 90 min.)  WL/PCC | 49  49 | 100 | IES-R | Turkey (Syrian refugees) | 33.32 (11.09)  34.04 (10.00) | 1 | Individual | ITT | 74 | Mass conflict | 8 |
| Akbarian et al., 2015  CBT/TF-CBT (10 sessions, 60 – 90 min.)  TAU/ACC (n.r.) | 14  14 | 100 | IES-R | Iran | 32.07 (5.76)  31.21 (6.10) | n.a. | Group  Individual | ITT | 79 | Various types | 5 |
| Andersen et al., 2021  TF-CBT + exercise/TF-CBT (10 sessions, 60 –   90 min.)  SPT + exercise/non-TF-PIs (10 sessions, 60   min.) | 43  46 | 100 | CAPS | Australia and Denmark | 39.71 (13.3)  44.49 (11.6) | 12 | Individual Individual | Compl. | 74  72 | Motor vehicle accident | 7 |
| Asukai et al., 2010  PE/TF-CBT (8-15 sessions, 90 min.)  TAU/ACC (n.r.) | 12  12 | 100 | CAPS | Japan | 27.10 (5.40)  31.40 (8.80) | r.b.i. | Individual  Individual | ITT | 88 | Various types | 7 |
| Basoglu et al., 2005  SSBT/TF-CBT (1 session, 60 min.)  WL/PCC | 31  28 | 100 | CAPS | Turkey | 36.30 (11.50) | r.b.i. | Individual | Compl. | 84 | Disaster | 6 |
| Basoglu et al., 2007  SSBT/TF-CBT (1 session, 69 – 130 min.)  WL/PCC | 16  15 | 100 | CAPS | Turkey | 34.00 (11.00) | r.b.i. | Individual | Compl. | 87 | Disaster | 6 |
| Beck et al., 2009  CBT/TF-CBT (14 sessions, 120 min.)  MA/PCC | 17  16 | 100 | CAPS | USA | 43.30 (12.80) | r.b.i. | Group  Individual | Compl. | 82 | Motor vehicle accident | 5 |
| Bellehsen et al., 2021  TMed/non-TF-PIs (16 sessions, 60 min.)  TAU/ACC (n.r.) | 20  20 | 100 | CAPS | USA | 52.9 (10.7)  50.3 (12.2) | r.b.i. | Group | ITT | 20  10 | Combat and other types | 5 |
| Belleville et al., 2018  IRT/other-TF-PIs (5 sessions, 60 min.)  WL/PCC | 19  20 | 100 | MPSS | Canada | 29.45 (9.05)  31.45 (10.32) | r.b.i. | Individual | ITT | 88 | Sexual assault | 6 |
| Bisson et al., 2020  3MDR/iEMDR (9 sessions, average 63.3 min)  WL/PCC | 16  19 | 100 | CAPS | UK | 40.2 (10.13)  44.0 (11.97) | r.b.i. | Individual | Compl. | 0 | Various types | 6 |
| Blanchard et al., 2003  CBT/TF-CBT (10 sessions, n.r.)  SPT/non-TF-PIs (10 sessions, n.r.)  WL/PCC | 37  36  25 | 77.78  77.78  87.50 | CAPS | USA | 40.60 (11.00)  40.60 (13.10)  42.10 (10.90) | 3 | Individual  Individual | ITT | 78  78  63 | Motor vehicle accident | 6 |
| Bohus et al., 2013  DBT-PTSD/TF-CBT (23 sessions, 45 min.)   TAU/ACC (n.r.) | 36  38 | 100 | CAPS | Germany | 35.14 (10.60)  36.71 (9.84) | 3 | Individual  Individual | ITT | 100 | Childhood sexual abuse | 7 |
| Bormann et al., 2008  Mantram interv./non-TF-PIs   (6 sessions, 90 min.)  WL/PCC | 14  15 | 100 | CAPS | USA | 56.00 (6.57) | n.r. | Group | Compl. | 0 | Combat | 5 |
| Bormann et al., 2013  Mantram interv./non-TF-PIs   (6 sessions, 90 min.)  TAU/ACC (n.r.) | 71  75 | 100 | CAPS | USA | 57.00 (10.10) | r.b.i. | Group  Individual | ITT | 3 | Combat | 6 |
| Boterhoven de Haan et al., 2020  EMDR/EMDR (12 sessions, 90 min.)  IR/other-TF-PIs (12 sessions, 90 min.) | 68  66 | 93 | CAPS | Australia  Germany  NL | 38.96 (11.51)  38.08 (10.85) | 12 | Individual  Individual | ITT | 80  73 | Childhood trauma, Various types | 7 |
| Brom et al., 1989  Trauma desensitization/TF-CBT (n.r.)  Psychodynamic Therapy/non-TF-PIs (n.r.)  WL/PCC | 28 26  20 | 100 | IES | NL | 42.00 (n.r.) | 3 | Individual | 0 | 79 | Various types | 4 |
| Brom et al., 2017  SE/non-TF-PIs (15 sessions, 60 min.)  WL/PCC | 32  28 | 100 | CAPS | Israel | 40.51 (13.05) | r.b.i. | Individual | Compl. | 46  57 | Various types | 7 |
| Bryant et al., 2003  IE/TF-CBT (8 sessions, 90 min.)  SC/ACC (8 sessions, 90 min.) | 20  18 | 100 | CAPS | Australia | 37.05 (12.31)  36.28 (8.41) | 6 | Individual  Individual | ITT | 52  52 | Nonsexual assault or motor vehicle accident | 6 |
| Bryant et al., 2011  CBT/TF-CBT (8 sessions, 60 min.)   TAU/ACC (8 sessions, n.r.) | 16  12 | 100 | PSS-I | Thailand | 42.30 (6.30)  43.9 (11.9) | 3 | Individual  Individual | ITT | 100  91 | Terror | 5 |
| Bryant et al., 2019  CBT long/TF-CBT (12 sessions, 90 min.)  WL/PCC | 33  34 | 100 | CAPS | Australia | 44.7 (10.7)  43.4 (7.8) | r.b.i. | Individual | Compl. | 12  29 | Various types | 4 |
| Buhmann et al., 2016  TF-CBT/TF-CBT (16 sessions, n.r.)  WL/PCC | 52 48 | 100 | HTQ | Denmark | 46.00 (8.00) 47.00 (8.00) | n.a. | Individual | ITT | 42 39 | Mass conflict | 7 |
| Butollo et al., 2016  DET/other-TF-PIs (24 sessions, n.r.)   CPT/TF-CBT (24 sessions, n.r.) | 74  67 | 100 | PDS | Germany | 37.99 (12.10)  33.67 (10.30) | 6 | Individual  Individual | ITT | 65  68 | Various types | 7 |
| Capezzani et al., 2013  EMDR/EMDR (8 sessions, n.r.)   CBT/TF-CBT (8 sessions, n.r.) | 21  11 | 100 | IES-R | Italy | 50.82 (7.64)  52.70 (8.68) | n.a. | Individual  Individual | ITT | 91 | Cancer | 4 |
| Carletto et al., 2016  EMDR/EMDR (10 sessions, 60 min.)  RT/ACC (n.r.) | 20  22 | 100 | CAPS | Italy | 39.52 (11.68)  40.66 (10.03) | n.a. | Individual | Compl. | 75  86 | MS | 6 |
| Carlson et al., 1998  EMDR/EMDR (12 sessions, 60-75 min.)   TAU/ACC (n.r.) | 10  12 | 100 | M-PTSD | USA | 52.70 (8.60)  46.90 (4.00) | r.b.i. | Individual  Individual | Compl. | 0  0 | Combat | 4 |
| Carlsson et al., 2018  CR/TF-CBT (16 sessions, 45-60 min.)  SIT/ACC (16 sessions, 45-60 min.) | 64  62 | 100 | HTQ | Denmark | 43.30 (9.50) | n.a. | Individual | ITT | 44 | Various types | 7 |
| Carter et al., 2013  SKY/non-TF-PIs (5 sessions, 22 hours total)  WL/PCC | 14  11 | 100 | CAPS | Australia | 58.50 (3.80)  58.40 (4.80) | r.b.i. | Group | Compl. | 0 | Combat | 5 |
| Castillo et al., 2016  CBT/TF-CBT (16 sessions, 90 min.)   WL/PCC | 42  42 | 100 | CAPS | USA | 35.90 (11.00) | r.b.i. | Group | ITT | 100 | Various types | 6 |
| Chard et al., 2005  CPT/TF-CBT (17 sessions à 90 min. group +   10 sessions à 60 min. individual)   MA/PCC (17 phone-calls à 5-10 min.) | 28  27 | 100 | CAPS | USA | 32.77 (8.87) | r.b.i. | Combination  Individual | Compl. | 100 | Childhood sexual abuse | 6 |
| Classen et al., 2020  TBG/non-TF-PIs (20 sessions, n.r.)  WL/PCC | 14  18 | 87.50 | PCL | Canada | 43.51 (10.01) | 6 | Group | Compl. | 100 | Childhood sexual and/or physical abuse | 1 |
| Cloitre et al., 2002  STAIR/TF-CBT (16 sessions, 8 à 60 min. + 8  à 90 min.)  WL/PCC | 31  27 | 100 | CAPS | USA | 34.00 (7.22) | r.b.i. | Individual | Compl. | 100 | Childhood sexual and/or physical abuse | 5 |
| Cloitre et al., 2010  STAIR/TF-CBT (16 sessions, 90 min.)  Skills comparator/ACC (n.r.) | 33  38 | 100 | CAPS | USA | 33.20 (n.r.)  37.10 (n.r.) | 6 | Individual  Individual | ITT | 100 | Childhood sexual and/or physical abuse | 8 |
| Cottraux et al., 2008  CBT/TF-CBT (10-16 sessions, 60-120 min.)  ST/non-TF-PIs (16 sessions, 60 min.) | 27  15 | 100 | PCL | France | 43.18 (10.60)  37.20 (9.20) | r.b.i. | Individual  Individual | Compl. | 70 | Various types | 6 |
| Davis et al., 2020  HYP/non-TF-PIs (16 sessions, 90 min.)  WLP/ACC (16 sessions, 90 min.) | 70  70 | 100 | CAPS | USA | 49.9 (12.6)  51.2 (13.3) | 7 | Group  Group | Compl. | 34 | Various types | 6 |
| Devilly et al., 1998  EMDR/EMDR (2 sessions, 90 min.)  TAU/ACC (n.r.) | 12  10 | 100 | M-PTSD | Australia | 50.10 (6.48) | r.b.i. | Individual n.r. | Compl. | 0 | Combat | 3 |
| Devilly et al., 1999  EMDR/EMDR (8 sessions, 90-165 min.)  TTP/TF-CBT (9 sessions, 90-165 min.) | 11  12 | 100 | PTSD-I | Australia | 37.96 (12.82) | r.b.i. | Individual  Individual | Compl. | 73  58 | Various types | 4 |
| Dorrepaal et al., 2012  SGT-ComplexPTSD/other-TF-PIs (20 sessions,   120 min.)  TAU/ACC (n.r.) | 38  33 | 100 | DTS | NL | 40.30 (10.70)  37.10 (10.30) | n.a. | Group  Individual | ITT | n.r. | Childhood sexual and/or physical abuse | 6 |
| Duffy et al., 2007  CBT/TF-CBT (12 sessions, n.r.)   WL/PCC | 29  29 | 100 | PDS | Northern Ireland | 44.1 (11.3)  43.7 (12.3) | r.b.i. | Individual | ITT | 34  45 | Terror | 7 |
| Dunne et al., 2012  CBT/TF-CBT (10 sessions, 60 min.)  WL/PCC | 13  13 | 100 | PDS | Australia | 32.54 (7.09) | r.b.i. | Individual | ITT | 50 | Chronic whiplash | 4 |
| Echeburua et al., 1997  EXP + CR/TF-CBT (6 sessions, 7h total)   PMR/ACC (6 sessions, 4,15h total) | 10  10 | 100 | Global Scale of PTSD | Spain | 20.00 (7.09) | 12 | Individual  Individual | ITT | 100 | Sexual assault | 3 |
| Ehlers et al., 2003  CT/TF-CBT (12 sessions, 60-90min.)  Self-help booklet/ACC (1 session, 40min.)   WL/PCC | 28  28  29 | 100 | PDS | UK | n.r. | 9 | Individual  Individual | ITT | n.r.  n.r.  n.r. | Motor vehicle accident | 7 |
| Ehlers et al., 2005  CT/TF-CBT (12 sessions, 60-90 min.)   WL/PCC | 14  14 | 100 | PDS | UK | 35.4 (10.9)  37.8 (11.2) | r.b.i. | Individual  Individual | ITT | 57  50 | Various types | 6 |
| Ehlers et al., 2014  Standard CT/TF-CBT (12 sessions, up to 20h total)   EFST/non-TF-PIs (12 sessions, up to 20h total)  WL/PCC | 31  30  30 | 100 | CAPS | UK | 41.50 (11.70)  37.80 (9.90)  36.80 (10.50) | 6 | Individual  Individual | ITT | 58  57  60 | Various types | 8 |
| Engel et al., 2015  DESTRESS-PC/i-non-TF-PIs (18 sessions, 15-30  min. + 30 min. homework)   TAU/ACC (3 sessions, 15 min.) | 29  29 | 100 | PCL | USA | 36.20 (7.75)  36.70 (9.75) | 2 | Individual | Compl. | 21  16 | Combat and military sexual trauma | 7 |
| Ertl et al., 2011  NET/TF-CBT (8 sessions, 90-120 min.)   SC/ACC (8 sessions, 90-120 min.)  WL/PCC | 26^a^  24^a^  28^a^ | 100 | CAPS | Uganda | 18.66 (3.77)  18.32 (4.30)  18.07 (3.55) | 12 | Individual  Individual | Compl. | 55  68  43 | Various types including childhood soldier victimization | 6 |
| Falsetti et al., 2008  TF-CBT/TF-CBT (12 sessions, 90 min.)   WL/PCC | 22  31 | 100 | MPSS | USA | 35.00 (9.82) | n.a. | Group | ITT | 100 100 | Various types | 7 |
| Fecteau et al., 1999  CBT/TF-CBT (4 sessions, 90-180 min.)   WL/PCC | 10  10 | 100 | CAPS | Canada | 41.30 (25–63) | r.b.i. | Individual | Compl. | 70  70 | Motor vehicle accident | 6 |
| Foa et al., 1991  PE/TF-CBT (9 sessions, 90 min.)   SC/ACC (9 sessions, 90 min.)  WL/PCC | 10  11  10 | 100 | SI-PTSD | USA | 32.70 (7.30)  34.20 (9.80)  32.00 (9.60) | r.b.i. | Individual  Individual | Compl. | 100 | Sexual assault | 5 |
| Foa et al., 1999  PE/TF-CBT (9 sessions, 2x120+7x90 min.)  SIT/ACC (9 sessions, 2x120+7x90 min.)  WL/PCC | 23  19  15 | 100 | PSS-I | USA | 34.90 (10.60) | 12 | Individual  Individual | Compl. | 100 | Sexual or non-sexual assault | 5 |
| Foa et al., 2005  PE/TF-CBT (12 sessions, 90-120 min.)  WL/PCC | 79  26 | 100 | PSS-I | USA | 31.30 (9.89) | r.b.i. | Individual | ITT | 100 | Sexual or non-sexual assault | 7 |
| Foa et al., 2018  spaced PE/TF-CBT (10 sessions, 90 min.)  PCT/non-TF-PIs (10 sessions, 90 min.)  MA/PCC (4 sessions, 10-15 min.) | 109  107  40 | 100 | PSS-I | USA | 32.89 (7.05)  32.54 (7.45)  32.70 (7.68) | 6 | Individual  Individual | ITT | 9  15  5 | Combat | 8 |
| Fonzo et al., 2017  PE/TF-CBT (9-12 sessions, 90 min.)  WL/PCC | 36  30 | 100 | CAPS | USA | 34.42 (10.23)  39.03 (10.35) | n.a. | Individual | ITT | 65 | Various types | 6 |
| Forbes et al., 2012  CPT/TF-CBT (12 sessions, 60 min.)  TAU/ACC (n.r.) | 30  29 | 100 | CAPS | Australia | 53.13 (13.97)  53.62 (13.33) | 3 | Individual  Individual | ITT | 7  0 | Combat | 7 |
| Ford et al., 2011  TARGET/other-TF-PIs (12 sessions, 50 min.)   PCT/non-TF-PIs (12 sessions, n.r.)   WL/PCC | 48  53  45 | 80  74  87 | CAPS | USA | 30.70 (6.90) | r.b.i. | Individual  Individual | ITT | 100  100  100 | Various types | 6 |
| Ford et al., 2013  TARGET/other-TF-PIs (12 sessions, 75 min.)   SGT/non-TF-PIs (12 sessions, 75 min.) | 38  34 | 82  74 | CAPS | USA | 34.60 (8.60)  38.00 (7.80) | n.a. | Individual  Group | Compl. | 100  100 | Various types | 6 |
| Galovski et al., 2012  MCPT/TF-CBT (4-18 sessions, n.r.)  SMDT/PCC (n.r.) | 38  37 | 100 | CAPS | USA | 39.80 (11.74) | r.b.i. | Individual | ITT | 69 | Sexual or physical assault | 8 |
| Gersons et al., 2000  BEP/other-TF-PIs (16 sessions, 60 min.)  WL/PCC | 22  20 | 100 | SI-PTSD | NL | 35.00 (6.00)  38.00 (7.00) | 3 | Individual | ITT | 18  5 | Various types | 7 |
| Ghafoori et al., 2017  PE/TF-CBT (12 sessions, 60-90 min.)  PCT/non-TF-PIs (12 sessions, 60-90 min.) | 47  24 | 100 | PCL-5 | USA | 35.10 (12.80)  35.30 (10.40) | n.a. | Individual  Individual | ITT | 83  83 | Physical assault and other types | 6 |
| Goldstein et al., 2017  IN EXC/non-TF-PIs (36 sessions, 60 min.)   WL/PCC | 21  26 | 89.36 | CAPS | USA | 46.80 (14.93) | n.a. | Group | ITT | 19 | Combat | 4 |
| Gray et al., 2019  RTM/TF-CBT (3 sessions, 120 min.)  WL/PCC | 37  37 | 100 | PSS-I | USA | 48.60 (13.30) | r.b.i. | Individual | ITT | 0 | Combat and other types | 7 |
| Gray et al., 2021  RTM/TF-CBT (3 sessions, ≤120 min.)  WL/PCC | 15  15 | 100 | PSS-I | USA | n.r.  n.r. | r.b.i. | Individual | ITT | 100 | Various types | 6 |
| Heffner et al., 2016 – Texas subsample only  Mantram/non-TF-PIs (8sessions, 60 min.)  PSY-EDU/ACC (10 sessions, 90 min.) | 18  15 | 82 | CAPS | USA | n.r. | n.a. | Individual  Group | Compl. | n.r. | Combat | 1 |
| Hensel-Dittmann et al., 2011  NET/TF-CBT (10 sessions, 90 min.)  SIT/ACC (10 sessions, 90 min.) | 11  10 | 100 | CAPS | Germany | n.r. | 6 | Individual  Individual | Compl. | n.r. | Various types | 6 |
| Hinton et al., 2011  CA-CBT/non-TF-PIs (14 sessions, 60 min.)  AMR/ACC (14 sessions, 60 min.) | 12  12 | 100 | PCL | USA (Cambod. refugees) | 47.60 (8.20)  51.40 (5.90) | 3 | Group  Group | ITT | 100 | n.r. | 5 |
| Hollifield et al., 2007  CBT/TF-CBT (12 sessions, 120 min.)   WL/PCC | 25  24 | 100 | PSS-SR | USA | 40.90 (13.40) 43.40 (13.50) | 3 | Group | ITT | 79  63 | Various types | 5 |
| Ivarsson et al., 2014  I-CBT/iTF-CBT (8 sessions, 28 min. of contact   to therapists on average)  SC/ACC (n.r.) | 31  31 | 100 | IES-R | Sweden | 44.80 (11.20)  47.20 (12.20) | r.b.i. | Individual | ITT | 77  87 | Various types | 8 |
| Jacob et al., 2014  NET/TF-CBT (8 sessions, 90-150 min.)  WL/PCC | 38 (17 orphans)  38 (16 orphans) | 100 | CAPS | Rwanda | widows  48.29 (13.40) orphans  25.06 (4.31)  widows  46.86 (11.73) orphans  24.00 (4.40) | r.b.i. | Individual | ITT | 89  95 | Various types | 8 |
| Jalal et al., 2020  CA-CBT/non-TF-PIs (14 sessions, 60 min.)  AMR/ACC (14 sessions, 60 min.) | 10 10 | 100 | PCL-C | South Africa | 28.2 (15 - n.r.) | n.a. | Individual | Compl. | 75 | Various types | 3 |
| Jensen, 1994  EMDR/EMDR (3 sessions, n.r.)  TAU/ACC (n.r.) | 13  12 | 100 | SI-PTSD | USA | 43.10 (2.84) | n.a. | Individual  Individual | Compl. | 0  0 | Combat | 3 |
| Johnson et al., 2011  HOPE/TF-CBT (12 sessions, 60-90 min.)  TAU/ACC (n.r.) | 34  34 | 88.60  85.70 | CAPS | USA | 32.55 (8.00) | 6 | Individual  Group | Compl. | 100 | IPV | 5 |
| Johnson et al., 2016  HOPE/TF-CBT (16 sessions, 60 min.)  TAU/ACC (n.r.) | 26  25 | 93.30  96.70 | CAPS | USA | 33.30 (10.48) 33.20 (10.39) | 6 | Individual  Group | Compl. | 100 | IPV and other types | 6 (Post + FU1) 5 (FU2) |
| Johnson et al., 2020  HOPE/TF-CBT (16 sessions, 50– 60 min.)  PCT+/non-TF-PIs (16 sessions, 50– 60 min.) | 83  89 | 100 | CAPS | USA | 34.34 (9.46)  35.87 (8.78) | 12 | Individual  Individual | Compl. | 100 | IPV | 6 |
| Karatzias et al., 2011  EMDR/EMDR (8 sessions, 60 min.)   EFT/non-TF-PIs (8 sessions, 60 min.) | 23 23 | 100 | CAPS | UK | 41.5 (10.8) 39.7 (10.9) | 3 | Individual Individual | ITT | 61 52 | Various types | 7 |
| Keane et al., 1989  IT/TF-CBT (14 sessions, 90 min.)   WL/PCC | 11  13 | 100 | MMPI  PTSD | USA | 34.70 (4.30)  34.50 (2.10) | r.b.i. | Individual | Compl. | 0  0 | Combat | 2 |
| Kearney et al., 2021  CPT-C/TF-CBT (12 sessions, 90 min.)  LKM/non-TF-PIs (12 sessions, 90 min.) | 93  90 | 100 | CAPS | USA | 56.1 (13.7)  58.2 (12.5) | 6 | Group  Group | ITT | 16 | Various types | 8 |
| Kelly et al., 2021  CPT/TF-CBT (12 sessions, 90 min.)  Trauma-Sensitive Yoga/non-TF-PIs (10   sessions, 60 min.) | 18  37 | 100 | CAPS | USA | 48.38 (11.1) | 3 | Group  Group | Compl. | 100 | Military sexual trauma | 5 (Post)  4 (FU) |
| Kent et al., 2011  ROT/non-TF-PIs (12 sessions, 90 min.)  WL/PCC | 23  19 | 100 | PDS | USA | 54.00 (8.34) | n.a. | Group | ITT | 33 | Various types | 4 |
| Krakow et al., 2000  IRT/other-TF-PIs (3 sessions, 2x 180+1x 60 min.)   WL/PCC | 42  48 | 95  95 | PSS-SR | USA | Compl.  40.10 (11.30)  36.00 (9.80) | r.b.i. | Group | Compl. | 100 | Sexual assault and other types | 3 |
| Krakow et al., 2001  IRT/other-TF-PIs (3 sessions, 2x 180+1x 60 min.)   WL/PCC | 45  52 | 95  95 | CAPS | USA | Compl.  40.00 (11.20)  36.00 (9.30) | r.b.i. | Group | Compl. | 100 | Sexual assault and other types | 4 |
| Krupnick et al., 2008  IPT/non-TF-PIs (16 sessions, 120 min.)  WL/PCC | 32  16 | 100 | CAPS | USA | 32.00 (10.20) | 4 | Group | ITT | 100 | Various types | 5 |
| Kubany et al., 2003  CTT-BW/TF-CBT (8-11 sessions, 90 min.)  WL/PCC | 19  18 | 100 | CAPS | USA | Compl.  36.80 (9.50) | r.b.i. | Individual | ITT | 100 | IPV and other types | 5 |
| Kubany et al., 2004  CTT-BW/TF-CBT (8-11 sessions, 90 min.)   WL/PCC | 63  62 | 100 | CAPS | USA | 42.20 (10.10) | r.b.i. | Individual | ITT | 100  100 | IPV and other types | 7 |
| Lang et al., 2019  CM/non-TF-PIs (10 sessions, 90 min.)  VC/ACC (10 sessions, 90 min.) | 14  14 | 100 | CAPS | USA | 49.10 (14.50) | n.a. | Group  Group | Compl. | 25 | Combat | 5 |
| Langkaas et al., 2017  PE/TF-CBT (10 sessions, 90-120 min.)  IR/other-TF-PIs (10 sessions, 90-120 min.) | 34  31 | 100 | PSS-I | Norway | 45.20 (9.70) | 12 | Individual  Individual | ITT | 58 | Various types | 8 |
| Latif et al., 2021  CatCBT GSH/non-TF-PIs (9 sessions, n.r.)  WL/PCC | 25  25 | 100 | IES-R | Pakistan | 27.4 (4.6)  26.3 (3.7) | n.a. | Individual | ITT | 100 | Domestic violence | 7 |
| Laugharne et al., 2016  PE/TF-CBT (12 sessions, n.r.)  EMDR/EMDR (12 sessions, n.r.) | 10  10 | 100 | CAPS | Australia | 40.10 (9.90) | n.a. | Individual  Individual | ITT | 70 | Various types | 7 |
| Lee et al., 2002  EMDR/EMDR (7 sessions, 90 min.)   PE + SIT/TF-CBT (7 sessions, 90 min.) | 12  12 | 100 | SI-PTSD | Australia | 35.30 (n.r.) | 3 | Individual  Individual | Compl. | 46  46 | Various types | 4 |
| Lehavot et al., 2021  DESTRESS-WV/iTF-CBT (16 sessions, n.r.)  Phone monitoring/ACC (9 sessions, 15 min.) | 51  51 | 100 | PCL-5 | USA | 49.9 (11.3)  48.9 (12.2) | 6 | Individual | ITT | 100 | Various types | 7 |
| Lely et al., 2019  NET/TF-CBT (11 sessions, 90 min.)  PCT/non-TF-PIs (11 sessions, 90 min.) | 18  15 | 100 | CAPS | NL | 62.65 (5.89)  62.47 (6.24) | 4 | Individual  Individual | ITT | 28  27 | Various types | 7 |
| Lewis et al., 2017  I-CBT/iTF-CBT (n.r.)  WL/PCC | 21  21 | 100 | CAPS | UK | 39.29 (12.70) | 3 | Individual | ITT | 60 | Various types | 7 |
| Lindauer et al., 2005  BEP/other-TF-PIs (16 sessions, 45-60 min.)  WL/PCC | 12  12 | 100 | SI-PTSD | NL | 37.60 (10.20)  40.30 (8.90) | n.a. | Individual | ITT | 42  67 | Various types | 7 |
| Littleton et al., 2016  FSTTP/i-non-TF-PIs (n.r.)  PESHW/ACC (n.r.) | 23 28 | 100 | PSS-I | USA | 22 (18-42) | 3 | Individual Individual | Compl. | 100 | Rape and other IPV | 6 (Post) 5 (FU1) |
| Litz et al., 2007  I-CBT/iTF-CBT (n.r.)  I-SC/iACC (n.r.) | 14  17 | 100 | PSS-I | USA | 39.86 (7.72)  38.63 (9.41) | 3 | Individual | Compl. | 19  25 | Combat or terror | 5 |
| Litz et al., 2021  CPT-C/TF-CBT (12 sessions, 60 min.)  AD/non-TF-PIs (6-8 sessions, 90 min.) | 33  37 | 100 | CAPS | USA | 30.30 (6.43)  29.80 (6.39) | r.b.i. | Individual  Individual | Compl. | 8 | Combat | 6 |
| Maguen et al., 2017  IOK/TF-CBT (6-8 sessions, 60-90min.)  TAU/ACC (n.r.) | 15  15 | n.r. | PCL-M | USA | 61.20 (13.00) | n.a. | Individual  Individual | Compl. | 0  0 | Combat | 4 |
| Marcus et al., 1997  EMDR/EMDR (8 sessions, 50 min.)  TAU/ACC (n.r., individual therapy: 50  min., group therapy: 90 min.) | 33  33 | 100 | MPSS | USA | Women 39.98 (18.00-73.00)  Men 44.78 (23.00-67.00) | n.a. | Individual  Combination | Compl. | 79  79 | Various types | 6 |
| Markowitz et al., 2015  PE/TF-CBT (10 sessions, 90 min.)  IPT/non-TF-PIs (14 sessions, 50 min.)   RT/ACC (10 sessions, 90 min.) | 38  40  32 | 100 | CAPS | USA | 47.50 (10.60)  41.00 (9.10)  34.80 (5.10) | 3 | Individual  Individual  Individual | ITT | 55  70  88 | Various types | 7 |
| Marks et al., 1998  EXP/TF-CBT (10 sessions, 90 min.)  RT/ACC (10 sessions, 90 min.) | 20  18 | 100 | CAPS | UK | 39.00 (11.00)  36.00 (10.00) | 3 | Individual  Individual | Compl. | 39  48 | Various types | 4 |
| McDonagh et al., 2005  CBT/TF-CBT (14 sessions, 7x120+7x90 min.)  PCT/non-TF-PIs (14 sessions, 7x120+7x90   min.)  WL/PCC | 29  22  23 | 100 | CAPS | USA | 39.80 (9.90)  39.60 (9.60)  42.00 (9.80) | 6 | Individual  Individual | ITT | 100 | Childhood sexual abuse | 6 |
| McGuire Stanbury et al., 2020  PE/TF-CBT (12 sessions, 90 min.)  EMDR/EMDR (12 sessions, 90 min.) | 10 10 | 100 | CAPS | Australia | 44.60 (12.18) 39.70 (9.55) | 3 | Individual Individual | Compl. | n.r. | Various types | 5 |
| McLean et al., 2020  Web-PE/TF-CBT (10 sessions, 60 min.)  PCT/non-TF-PIs (10 sessions, 60 min.) | 10^a^  11^a^ | 100 | PCL-5 | USA | 38.7 (8.9)  41.5 (6.5) | 3 | Individual | Compl. | 25 | Combat | 5 |
| Mitchell et al., 2014  YI/non-TF-PIs (12 sessions, 75 min.)   WL/PCC | 20  18 | 71  71 | PCL | USA | 44.37 (12.37) | 1 | Group | ITT | 100  100 | Various types | 5 |
| Monson et al., 2006  CPT/TF-CBT (12 sessions, n.r.)  WL/PCC | 30  30 | 100 | CAPS | USA | 54.00 (6.30) | 1 | Individual | ITT | 7  13 | Combat | 7 |
| Monson et al., 2012  CBCT/TF-CBT (15 sessions, n.r.)  WL/PCC | 20  20 | 100 | CAPS | USA & Canada | 40.40 (11.30)  33.80 (10.50) | r.b.i. | Couple | ITT | 65  85 | Various types | 7 |
| Morath et al., 2014  NET/TF-CBT (12 sessions, n.r.)   WL/PCC | 19  19 | 100 | CAPS | Germany | 28.70 (9.54)  30.10 (8.21) | r.b.i. | Individual | Compl. | 32  32 | Various types | 6 |
| Mueser et al., 2008  CBT/TF-CBT (16 sessions, n.r.)  TAU/ACC(n.r.) | 32  27 | 100 | CAPS | USA | 45.13 (9.83)  43.30 (11.41) | 6 | Individual  Individual | Compl. | 76  82 | Various types | 5 |
| Nacasch et al., 2011  PE/TF-CBT (9-15 sessions, 90-120 min.)  TAU/ACC (n.r., 60 min.) | 15  15 | 100 | PSS-I | Israel | 34.80 (11.40)  33.70 (11.90) | 12 | Individual  Individual | ITT | n.r. | Combat and terror | 7 |
| NCT00607815  CPT/TF-CBT (12 sessions, n.r.)  PCT/non-TF-PIs (12 sessions, n.r.) | 43  36 | 100 | CAPS | USA | 29.50 (7.11)  32.11 (7.85) | 12 | Individual  Individual | ITT | 0  0 | Combat | 5 |
| Neuner et al., 2004  NET/TF-CBT (4 sessions, 90-120 min.)   SC/ACC (4 sessions, 90-120 min.)   PSY-EDU/PCC (1 session, n.r.) | 17  14  12 | 100 | PDS | Uganda (Sundanese refugees) | 31.90 (6.70)  33.80 (7.90)  34.20 (6.90) | 12 | Individual  Individual  Individual | Compl. | 53  57  75 | Various types | 6 |
| Neuner et al., 2008  NET/TF-CBT (6 sessions, n.r.)   TC/ACC (6 sessions, n.r.)   WL/PCC | 111  111  55 | 100 | PDS | Uganda (Rwandan & Somal. refugees) | 34.40 (12.20)  35.20 (12.80)  35.60 (14.00) | 9 | Individual  Individual | ITT | 51  53  49 | Various types | 7 |
| Neuner et al., 2010  NET/TF-CBT (5-17 sessions, 120 min.)  TAU/ACC (n.r.) | 14^a^  16^a^ | 100 | PDS | Germany (refugees) | 31.10 (7.80)  31.60 (7.70) | 6 | Individual Individual | ITT | 31  31 | Physical torture and other types | 7 |
| Nidich et al., 2018  PE/TF-CBT (12 sessions, 90 min.)  TMed/non-TF-PIs (12 sessions, 90 min.)  HE/ACC (12 sessions, 90 min.) | 68  68  66 | 100 | CAPS | USA | 48.50 (15.60)  46.40 (14.30)  46.20 (16.40) | n.a. | Individual  Individual  Individual | ITT | 18  18  15 | Combat | 7 |
| Nijdam et al., 2012  EMDR/EMDR (17 sessions, 90 min.)  BEP/other-TF-PIs (16 sessions, 45-60 min.) | 70  70 | 100 | SI-PTSD | NL | 38.30 (12.20)  37.30 (10.60) | n.a. | Individual  Individual | Compl. | 51  61 | Assault and other types | 7 |
| Niles et al., 2012  teleMM/i-non-TF-PIs (8 sessions; 2 f2f à 45 min., 6   tele à 20 min.)  telePSY-EDU/ACC (8 sessions; 2 f2f à 45 min., 6   tele à 20 min.) | 13  14 | 100 | CAPS (post) PCL (FU) | USA | 52.00 (13.00) | 1.50 | Individual  Individual | Compl. | 0 | Combat or mass violence (as peacekeepers) | 5 |
| Orang et al., 2018  NET/TF-CBT (12 sessions, 120-150 min.)  TAU/ACC (12 sessions, 90–120 min.) | 17^a^  17^a^ | 100 | PSS-I | Iran | 38.04 (9.69)  37.28 (7.92) | 6 | Individual Combinat. | Compl. | 100 | IPV | 5 |
| Pacella et al., 2012  PE/TF-CBT (10 sessions, 90-120 min.)  WL/PCC | 34  24 | 100 | PSS-I | USA | 46.00 (5.80)  48.00 (7.00) | 3 | Individual | ITT | 37 | HIV-related and others | 8 |
| Paunovic, 2011  EIT/TF-CBT (3-9 sessions, 60-120 min.)   WL/PCC | 14  15 | 100 | CAPS | Sweden | 37.10 (13.80)  37.30 (10.20) | r.b.i. | Individual | Compl. | 50  47 | Sexual assault and other types | 2 |
| Power et al., 2002  EXP + CR/TF-CBT (10 sessions, 90 min.)  EMDR/EMDR (10 sessions, 90 min.)  WL/PCC | 21  27  24 | 100 | SI-PTSD | UK | 43.20 (11.00)  38.60 (11.80)  36.50 (11.60) | r.b.i. | Individual  Individual | Compl. | 38  44  42 | Various types | 6 (EMDR vs WL)  5 (other comparisons) |
| Rauch et al., 2015  PE/TF-CBT (10-12 sessions, 80 min.)   PCT/non-TF-PIs (10-12 sessions, 80 min.) | 11  15 | 100 | CAPS | USA | 31.90 (7.60) | n.a. | Individual  Individual | Compl. | 8 | Combat | 4 |
| Ready et al., 2018  GBET/TF-CBT (32 sessions, 240 min.)  GPCT/non-TF-PIs (37 sessions, 90 min.) | 40  41 | 100 | CAPS | USA | 61.40 (2.60) | 12 | Group  Group | Compl. | n.r. | Combat | 5 |
| Reger et al., 2016  PE/TF-CBT (10 sessions, 90-120 min.)  WL/PCC | 54  54 | 100 | CAPS | USA | 30.89 (7.09)  30.39 (6.45) | r.b.i. | Individual | ITT | 4  2 | Nonsexual  assault | 8 |
| Resick et al., 2002  CPT/TF-CBT (12 sessions, 13 hours total)  WL/PCC | 62  47 | 100 | SI-PTSD | USA | 32.00 (9.90) | r.b.i. | Individual | ITT | 100  100 | Sexual assault | 6 |
| Resick et al., 2015  GCPT/TF-CBT (12 sessions, 90 min.)  GPCT/non-TF-PIs (12 sessions, 90 min.) | 56  52 | 100 | PSS-I | USA | 31.80 (7.30)  32.40 (7.90) | 12 | Group  Group | ITT | 7  8 | Combat and other types | 6 |
| Robjant et al., 2019  NET/TF-CBT (6 sessions á 90-120 min.   individual, 6 sessions á 90-120 group)  TAU/ACC (n.r.) | 45^a^ 43^a^ | 100 | PSS-I | DRC | 18 (16-25)  18 (16-25) | 9 | Combination  Individual | Compl. | 100 | Former child soldier victimization | 7 |
| Rothbaum et al., 2005  EMDR/EMDR (9 sessions, 90 min.)   PE/TF-CBT (9 sessions, 90 min.)   WL/PCC | 20  20  20 | 100 | CAPS | USA | 33.80 (11.00) | 6 | Individual  Individual | Compl. | 100 | Sexual assault | 5 |
| Sautter et al., 2015  SAT/TF-CBT (12 sessions, 60 min.)  PFE/ACC (12 sessions, 60 min.) | 29  28 | 100 | CAPS | USA | 32.55 (6.16)  33.71 (7.01) | 3 | Couple  Couple | ITT | 0  4 | Combat | 7 |
| Schaal et al., 2009  NET/TF-CBT (4 sessions, 120-150 min.)  IPT/non-TF-PIs (4 sessions, 120-150 min.) | 12 14 | 100 | CAPS | Rwanda | 19.42 (3.59) | 6 | Individual  Group | ITT | 62 | Mass conflict | 7 |
| Scheck et al., 1998  EMDR/EMDR (2 sessions, 90 min.)  AL/ACC (2 sessions, 90 min.) | 28  29 | 77 | IES | USA | 20.93  (16.00-25.00) | r.b.i. | Individual  Individual | Compl. | 100 | Various types | 5 |
| Schnurr et al., 2003  TFGT/TFCBT (30 sessions, 90-120 min.)  GPCT/non-TF-PIs (30 sessions, 90-120 min.) | 162  160 | 100 | CAPS | USA | 50.60 (3.70)  50.80 (3.80) | 12 | Group  Group | Compl. | 0 | Combat | 6 |
| Schnurr et al., 2007  PE/TF-CBT (10 sessions, 90 min.)  PCT/non-TF-PIs (10 sessions, 90 min.) | 141 143 | 100 | CAPS | USA | 44.60 (9.39)  44.90 (9.47) | 6 | Individual  Individual | ITT | 100 | Sexual trauma and other types | 7 |
| Sloan et al., 2012  WET/TF-CBT (3 sessions, 20 min.)   Neutral Writing/PCC (3 sessions, 20 min.) | 21^a^  21^a^ | 100 | PSS-I | USA | 18.90 (1.10) | 1 | Individual  Individual | Compl. | n.r. | Various types | 5 |
| Sloan et al., 2012  WET/TF-CBT (5 sessions, 1x60+4x40 min.)   WL/PCC | 22  24 | 100 | CAPS | USA | 40.65  (13.10) | 3 | Individual | ITT | 65  65 | Motor vehicle accident | 6 |
| Sloan et al., 2018  GCBT/TF-CBT (14 sessions, 120 min.)  GPCT/non-TF-PIs (14 sessions, 120 min.) | 98  100 | 100 | CAPS | USA | 54.40 (11.44)  57.22 (12.51) | 12 | Group  Group | ITT | 0 | Combat and other types | 8 |
| Spence et al., 2013  iCBT/iTF-CBT (10 sessions, 90 min.)  WL/PCC (10 sessions, 90 min.) | 23  21 | 100 | PCL-C | Australia | 43.00 (15.20)  42.00 (10.40) | n.a. | Individual | ITT | 74  89 | Various types | 7 |
| Stenmark et al., 2011  NET/TF-CBT (10 sessions, 90 min.)  TAU/ACC (10 sessions, 90 min.) | 33  21 | 100 | CAPS | Norway  (refugees) | 34.50 (11.10)  36.60 (11.00) | 6 | Individual  Individual | Compl. | 33  27 | Various types | 7 |
| Suris et al., 2013  CPT/TF-CBT (12 sessions, n.r.)  PCT/non-TF-PIs (12 sessions, n.r.) | 52  34 | 100 | CAPS | USA | 44.60 (10.50)  48.40 (8.20) | 6 | Individual  Individual | Compl. | 83  88 | Military sexual trauma | 7 |
| Taylor et al., 2003  PE/TF-CBT (8 sessions, 90 min.)  EMDR/EMDR (8 sessions, 90 min.)  RT/ACC (8 sessions, 90 min.) | 15  15  15 | 100 | CAPS | Canada | 37.00 (10.00) | 3 | Individual  Individual  Individual | ITT | 75 | Various types | 6 |
| ter Heide et al., 2016  EMDR/EMDR (9 sessions, 3x60+6x90 min.) TAU/ACC (12 sessions, 60 min.) | 32  29 | 100 | CAPS | NL | 43.10 (10.70)  39.80 (11.90) | 3 | Individual  Individual | Compl | 17  39 | Various types | 7 |
| Thorp et al., 2019  PE/TF-CBT (12 sessions, 90 min.)  RT/ACC (12 sessions, 90 min.) | 29  38 | 100 | CAPS | USA | 66.51 (6.21)  64.43 (4.49) | 6 | Individual  Individual | ITT | 0 | Combat | 6 (Post) 5 (FU2) |
| Tylee et al., 2017  RTM/TF-CBT (3 sessions, 120 min.)  WL/PCC | 15 15 | 100 | PSS-I | USA | 49.0 (19.5) 42.6 (15.9) | r.b.i. | Individual | Compl. | 0 | Combat and other types | 5 |
| van den Berg et al., 2015  EMDR/EMDR (8 sessions, 90 min.)  PE/TF-CBT (8 sessions, 90 min.)  WL/PCC | 55  55  47 | 100 | CAPS | NL | 40.40 (11.30)  42.60 (10.30)  40.30 (9.70) | 6 | Individual  Individual | ITT | 55  57  51 | Various types | 8 |
| van der Kolk et al., 2007  EMDR/EMDR (8 sessions, 90 min.)  Placebo/ACC (8 sessions, 20-30 min.) | 29  29 | 100 | CAPS | USA | 38.70 (14.30)  35.70 (13.40) | r.b.i. | Individual Individual | ITT | 76  86 | Various types | 8 |
| van der Kolk et al., 2014  YI/non-TF-PIs (10 sessions, 60 min.)  HE/ACC (10 sessions, 60 min.) | 32  32 | 100 | CAPS | USA | 41.50 (12.20)  44.30 (11.90) | n.a. | Group  Group | ITT | 100  100 | Various types | 5 |
| van Gelderen et al., 2020  3MDR/iEMDR (6 sessions, 70-90 min.)  NTCC/ACC | 22  21 | 100 | CAPS | NL | 42.41 (9.80) 41.93 (9.12) | r.b.i. | Individual Individual | ITT | 4.5  0 | Various types | 7 |
| Vaughan et al., 1994  EMDR/EMDR (3-5 sessions, 50 min.)  IHT/TF-CBT (3-5 sessions, 50 min.)   AMR/ACC  WL/PCC | 12  13  11  11 | 78 | SI-PTSD | Australia | 32.00 (14.70) | 3 | Individual  Individual  Individual | Compl. | 64 | Various types | 3 |
| Vera et al., 2022  PE/TF-CBT (12-15 sessions, 90 min.)  AMR/ACC (12-15 sessions, 90 min.) | 39  37 | 100 | CAPS | Puerto Rico | 44.08 (11.53)  43.16 (12.73) | 3 | Individual  Individual | ITT | 74  90 | Various types | 6 |
| Wagner et al., 2019  BA/non-TF-PIs (8 sessions, 45 min.)  TAU/ACC (6 sessions, n.r.) | 30 24 | 100 | CAPS | USA | 30.2 (6.4) 29.9 (7.1) | 3 | Individual  Combination | Compl. | 7 5 | Military | 6 |
| Wahbeh et al., 2016  MM/non-TF-PIs (6 sessions, 60 min.)  SQ/PCC (6 sessions, n.r.) | 24  22 | 100 | PCL | Canada | 53.30 (12.60)  53.00 (11.80) | n.a. | Individual  Individual | Compl. | 7  4 | Combat | 3 |
| Wells et al., 2012  MCT/non-TF-PIs (8 sessions, n.r.)  WL/PCC | 10  10 | 100 | PDS | UK | 33.40 (13.40)  41.30 (13.70) | r.b.i. | Individual | ITT | 60  50 | Various types | 6 |
| Wells et al., 2015  MCT/non-TF-PIs (8 sessions, 60 min.)  PE/TF-CBT (8 sessions, 60 min.)  WL/PCC | 10  10  10 | 100 | PDS | UK | 40.60 (11.90)  40.50 (10.90)  42.70 (18.50) | 3 | Individual  Individual | Compl. | 36  36  40 | Various types | 6 |
| Yehuda et al., 2014  PE/TF-CBT (12 sessions, 90 min.)  MA/PCC | 25  12 | 100 | CAPS | UA | 48,86 | r.b.i. | Individual | Compl. | 11 | Combat | 7 |
| Yurtsever et al., 2018  EMDR G-TEP/EMDR (2 sessions, 240 min.) WL/PCC | 18  29 | 100 | IES-R | Turkey (Syrian refugees) | 39.89 (10.96)  35.93 (11.10) | 1 | Group | Compl. | 72  79 | Various types | 5 |
| Zang et al., 2013  NET/TF-CBT (4 sessions, 60-90 min.)  WL/PCC | 11  11 | 100 | IES-R | China | 56.64 (12.22)  54.82 (11.59) | r.b.i. | Individual | ITT | 73  82 | Disaster | 7 |
| Zang et al., 2014  NET/TF-CBT (4 sessions, 60-90 min.)  WL/PCC | 10  10 | 100 | IES-R | China | 53.50 (1.24)  50.90 (1.23) | r.b.i. | Individual | ITT | 90  100 | Disaster | 7 |
| Zemestani et al., 2022  TF-CBT/TF-CBT (12 sessions, 90 min.)  WL/PCC | 24  24 | 100 | PCL-5 | Iraq | 33.45 (5.46)  32.37 (5.27) | 1 | Individual | ITT | 100  100 | Various types | 7 |
| Zlotnick et al., 1997  AM/Non-TF-PIs (15 sessions, 120 min.) TAU/ACC (n.r.) | 16  17 | 100 | DTS | USA | 39.00 (9.59) | n.a. | Group  Individual | Compl. | 100 | Childhood sexual abuse | 3 |
| Zoellner et al., 2017  IE + Placebo/TF-CBT (5 sessions, 50 min.)  WL/PCC | 16  11 | 100 | PSS-I | USA | 37.50 (12.40) | 1 | Individual | ITT | 71 | Various types | 6 |

^a^no posttreatment assessment, hence, FU sample sizes reported. ACC – Active Control Condition, AD – Adaptive Disclosure, AL – Active Listening, AM – Affect-Management, AMR – Applied Muscle Relaxation, BA – Behavioral Activation, BEP – Brief Elective Psychotherapy, CA-CBT – Culturally Adapted Cognitive Behavioral Therapy, CAPS – Clinician-Administered PTSD Scale, CatCBT GSH – culturally adapted CBT-based guided self-help, CBCT – Cognitive-Behavior Couple Therapy, CBT – Cognitive-Behavioral Therapy, CM – Compassion Meditation, Compl. – Completer analysis, CPT – Cognitive-Processing Therapy, CR – Cognitive Restructuring, CPT-C – Cognitive-Processing Therapy (cognitive only), CT – Cognitive Therapy, CTT-BW – Cognitive Trauma-Therapy for Battered Women, DBT-PTSD – Dialectic Behavior Therapy for PTSD, DESTRESS-PC – Delivery of Self Training and Education for Stressful Situations-Primary Care version, DESTRESS-WV – Delivery of Self Training and Education for Stressful Situations – Women Veterans version, DET – Dialogical Exposure Therapy, DRC – Democratic Republic of the Congo, DTS – Davidson Trauma Scale, EFST – Emotion Focused Supportive Therapy, EFT – Emotional Freedom Techniques, EIT – Exposure Inhibition Therapy, EMDR – Eye Movement Desensitization and Reprocessing, EMDR G-TEP – EMDR Group Traumatic Episodic Protocol, EXP – Exposure, EXP + CR – Exposure plus Cognitive Restructuring, f2f – face to face, FSTTP – From Survivor to Thriver Program, GBET – Group-Based Exposure Therapy, GCBT – Group Cognitive Behavioral Therapy, GCPT – Group Cognitive-Processing Therapy, GPCT – Group Present-Centered Therapy, HE – Health Education, HOPE – Helping to Overcome PTSD through Empowerment, HTQ – Harvard Trauma Questionnaire, HYP – Holistic Yoga Program, I-CBT – Internet-based Cognitive Behavioral Therapy, IE – Imaginal Exposure, iEMDR – internet-delivered/technology-delivered Eye Movement Desensitization and Reprocessing, IE + Placebo – Imaginal Exposure plus pill placebo, IES – Impact of Event Scale, IES-R – Impact of Event Scale - Revised, IHT – Image Habituation Training, IN EXC – Integrative Exercise, i-non-TF-PIs – internet-delivered/technology-delivered non-trauma focused psychological interventions, IOK – Impact Of Killing, IPT – Inter-Personal Therapy, IPV – Intimate Partner Violence, IR – Imagery Rescripting, IRT – Imagery Rehearsal Therapy, I-SC – Internet-based Supportive Counseling, IT – Implosive Therapy, iTF-CBT – (mainly or completely) internet-delivered/technology-delivered Trauma-Focused Cognitive Behavioral Therapy, ITT – Intent-To-Treat analysis, LKM – Loving-Kindness Meditation, MA – Minimal Attention, MCPT – Modified Cognitive-Processing Therapy, MCT – Meta-Cognitive Therapy, min. – minutes, MM – Mindfulness Meditation, MMPI – Minnesota Multiphasic Personality Inventory, MPSS – Modified PTSD Symptom Scale, M-PTSD – Mississippi scale for combat-related PTSD, MS – Multiple Sclerosis, n.a. – not applicable, NET – Narrative Exposure Therapy, NET-R – Narrative Exposure Therapy Revised, NL – the Netherlands, non-TF-PIs – non-trauma focused psychological interventions, n.r. – not reported, NTCC – Non-specific Treatment Component Control, other-TF-PIs – other trauma focused psychological interventions (i.e., non-TF-CBT & non-EMDR interventions), PCC – Passive Control Condition, PCL-5 – PTSD Checklist for DSM-5, PCL – PTSD Check-List – Civilian Version, PCL-C – PTSD Check-List - Civilian Version, PCL-M – PTSD Check-List - Military Version, PCT – Present-Centered Therapy, PCT+ – adapted version of Present-Centered Therapy, PDS – Posttraumatic Diagnostic Scale, PE – Prolonged Exposure, PE + CR – Prolonged Exposure + Cognitive Restructuring, PE + SIT – Prolonged Exposure plus Stress Inoculation Training, PESHW – Psycho-educational self-help website, PFE – PTSD Family Education, Placebo – pill placebo control group, PMR – Progressive Muscle Relaxation, PSS-I – PTSD Symptom Scale – Interview, PSS-SR – PTSD Symptom Scale – Self-Report, PSY-EDU – Psychoeducation, PTSD-I – PTSD Interview, r.b.i. – reported but irrelevant (i.e., no meaningful group comparison possible at follow-up), ROT – Resilience Oriented Treatment, RT – Relaxation Therapy/Training, RTM – Reconsolidation of Traumatic Memories, SAT – Structured Approach Therapy, SC – Supportive Counselling, SE – Somatic Experiencing, SGT – Supportive Group Therapy, SGT-ComplexPTSD – Stabilizing Group Treatment for Complex PTSD, SI-PTSD – Structured Interview for PTSD, SIT – Stress Inoculation Training, SKY – Sudarshan Kriya Yoga, SMDT – Symptom-Monitoring Delayed Treatment group, SPT – Supportive Psychotherapy, SQ – Sitting Quietly, SSBT – Single-Session Behavioral Treatment, STAIR – Skills Training in Affective and Interpersonal Regulation, TARGET – Trauma Affect Regulation Guide for Education and Therapy, TAU – Treatment-As-Usual, TBG – Trauma and the Body Group, TC – Trauma Counselling, teleMM – telehealth Mindfulness Meditation, telePSY-EDU – telehealth Psychoeducation, TF-CBT – Trauma-Focused Cognitive Behavioral Therapy, TFGT – Trauma-Focused Group Psychotherapy, TMed – Transcendental Meditation, TTP – Trauma Treatment Protocol, UK – United Kingdom, USA – United States of America, VC – Veteran.Calm, WL – Wait-List control condition, WLP – Wellness Lifestyle Program, YI – Yoga Intervention

**Appendix G:** **References of trials included in the present network and pairwise meta-analysis**

1. Acarturk C, Konuk E, Cetinkaya M, et al. The efficacy of eye movement desensitization and reprocessing for post-traumatic stress disorder and depression among Syrian refugees: results of a randomized controlled trial. *Psychol Med*. 2016;46(12):2583-2593. doi:10.1017/S0033291716001070.
2. Akbarian F, Bajoghli H, Haghighi M, Kalak N, Holsboer-Trachsler E, Brand S. The effectiveness of cognitive behavioral therapy with respect to psychological symptoms and recovering autobiographical memory in patients suffering from post-traumatic stress disorder. *Neuropsychiatr Dis Treat*. 2015;11(19):395-404. doi:10.2147/NDT.S79581.
3. Andersen TE, Ravn SL, Armfield N, Maujean A, Requena SS, Sterling M. Trauma-focused cognitive behavioural therapy and exercise for chronic whiplash with comorbid posttraumatic stress disorder: a randomised controlled trial. *Pain*. 2021;162(4):1221-1232.
4. Asukai N, Saito A, Tsuruta N, Kishimoto J, Nishikawa T. Efficacy of exposure therapy for Japanese patients with posttraumatic stress disorder due to mixed traumatic events: A randomized controlled study. *J Trauma Stress*. 2010;23(6):744-750. doi:10.1002/jts.20589.
5. Başoglu M, Salcioglu E, Livanou M. A randomized controlled study of single-session behavioural treatment of earthquake-related post-traumatic stress disorder using an earthquake simulator. *Psychol Med*. 2007;37(2):203-213. doi:10.1017/S0033291706009123.
6. Başoğlu M, Salcioğlu E, Livanou M, Kalender D, Acar G. Single-session behavioral treatment of earthquake-related posttraumatic stress disorder: a randomized waiting list controlled trial. *J Trauma Stress*. 2005;18(1):1-11. doi:10.1002/jts.20011.
7. Beck JG, Coffey SF, Foy DW, Keane TM, Blanchard EB. Group cognitive behavior therapy for chronic posttraumatic stress disorder: an initial randomized pilot study. *Behav Ther*. 2009;40(1):82-92. doi:10.1016/j.beth.2008.01.003.
8. Bellehsen M, Stoycheva V, Cohen BH, Nidich S. A Pilot Randomized Controlled Trial of Transcendental Meditation as Treatment for Posttraumatic Stress Disorder in Veterans. *J Trauma Stress*. 2022;35(1):22-31.
9. Belleville G, Dubé-Frenette M, Rousseau A. Efficacy of Imagery Rehearsal Therapy and Cognitive Behavioral Therapy in Sexual Assault Victims With Posttraumatic Stress Disorder: A Randomized Controlled Trial. *J Trauma Stress*. 2018;31(4):591-601. doi:10.1002/jts.22306.
10. Bisson JI, van Deursen R, Hannigan B, et al. Randomized controlled trial of multi-modular motion-assisted memory desensitization and reconsolidation (3MDR) for male military veterans with treatment-resistant post-traumatic stress disorder. *Acta Psychiatr Scand*. 2020;142(2):141-151. doi:10.1111/acps.13200.
11. Blanchard EB, Hickling EJ, Devineni T, et al. A controlled evaluation of cognitive behaviorial therapy for posttraumatic stress in motor vehicle accident survivors. *Behav Res Ther*. 2003;41(1):79-96. doi:10.1016/S0005-7967(01)00131-0.
12. Bohus M, Dyer AS, Priebe K, et al. Dialectical behaviour therapy for post-traumatic stress disorder after childhood sexual abuse in patients with and without borderline personality disorder: a randomised controlled trial. *Psychother Psychosom*. 2013;82(4):221-233. doi:10.1159/000348451.
13. Bormann JE, Thorp S, Wetherell JL, Golshan S. A spiritually based group intervention for combat veterans with posttraumatic stress disorder: feasibility study. *Journal of Holistic Nursing*. 2008;26(2):109-116. doi:10.1177/0898010107311276.
14. Bormann JE, Thorp SR, Wetherell JL, Golshan S, Lang AJ. Meditation-based mantram intervention for veterans with posttraumatic stress disorder: A randomized trial. *Psychol Trauma*. 2013;5(3):259-267. doi:10.1037/a0027522.
15. Boterhoven de Haan, Katrina L., Lee CW, Fassbinder E, et al. Imagery rescripting and eye movement desensitisation and reprocessing as treatment for adults with post-traumatic stress disorder from childhood trauma: randomised clinical trial. *Br J Psychiatry*. 2020;217(5):609-615. doi:10.1192/bjp.2020.158.
16. Brom D, Kleber RJ, Defares PB. Brief psychotherapy for posttraumatic stress disorders. *J Consult Clin Psychol*. 1989;57(5):607-612. doi:10.1037//0022-006x.57.5.607.
17. Brom D, Stokar Y, Lawi C, et al. Somatic Experiencing for Posttraumatic Stress Disorder: A Randomized Controlled Outcome Study. *J Trauma Stress*. 2017;30(3):304-312. doi:10.1002/jts.22189.
18. Bryant RA, Moulds ML, Guthrie RM, Dang ST, Nixon RDV. Imaginal exposure alone and imaginal exposure with cognitive restructuring in treatment of posttraumatic stress disorder. *J Consult Clin Psychol*. 2003;71(4):706-712. doi:10.1037/0022-006x.71.4.706.
19. Bryant RA, Ekasawin S, Chakrabhand S, Suwanmitri S, Duangchun O, Chantaluckwong T. A randomized controlled effectiveness trial of cognitive behavior therapy for post-traumatic stress disorder in terrorist-affected people in Thailand. *World Psychiatry*. 2011;10(3):205-209. doi:10.1002/j.2051-5545.2011.tb00058.x.
20. Bryant RA, Kenny L, Rawson N, et al. Efficacy of exposure-based cognitive behaviour therapy for post-traumatic stress disorder in emergency service personnel: a randomised clinical trial. *Psychol Med*. 2019;49(9):1565-1573. doi:10.1017/S0033291718002234.
21. Buhmann CB, Nordentoft M, Ekstroem M, Carlsson J, Mortensen EL. The effect of flexible cognitive-behavioural therapy and medical treatment, including antidepressants on post-traumatic stress disorder and depression in traumatised refugees: pragmatic randomised controlled clinical trial. *Br J Psychiatry*. 2016;208(3):252-259. doi:10.1192/bjp.bp.114.150961.
22. Butollo W, Karl R, König J, Rosner R. A Randomized Controlled Clinical Trial of Dialogical Exposure Therapy versus Cognitive Processing Therapy for Adult Outpatients Suffering from PTSD after Type I Trauma in Adulthood. *Psychother Psychosom*. 2016;85(1):16-26. doi:10.1159/000440726.
23. Capezzani L, Ostacoli L, Cavallo M, et al. EMDR and CBT for Cancer Patients: Comparative Study of Effects on PTSD, Anxiety, and Depression. *J EMDR Prac Res*. 2013;7(3):134-143. doi:10.1891/1933-3196.7.3.134.
24. Carletto S, Borghi M, Bertino G, et al. Treating Post-traumatic Stress Disorder in Patients with Multiple Sclerosis: A Randomized Controlled Trial Comparing the Efficacy of Eye Movement Desensitization and Reprocessing and Relaxation Therapy. *Front Psychol*. 2016;7(21):526. doi:10.3389/fpsyg.2016.00526.
25. Carlson JG, Chemtob CM, Rusnak K, Hedlund NL, Muraoka MY. Eye movement desensitization and reprocessing (EDMR) treatment for combat-related posttraumatic stress disorder. *J Trauma Stress*. 1998;11(1):3-24. doi:10.1023/A:1024448814268.
26. Carlsson J, Sonne C, Vindbjerg E, Mortensen EL. Stress management versus cognitive restructuring in trauma-affected refugees-A pragmatic randomised study. *Psychiatry Res*. 2018;266:116-123. doi:10.1016/j.psychres.2018.05.015.
27. Carter JJ, Gerbarg PL, Brown R, et al. Multi-Component Yoga Breath Program for Vietnam Veteran Post Traumatic Stress Disorder: Randomized Controlled Trial. *J Trauma Stress Disor Treat*. 2013;02(03). doi:10.4172/2324-8947.1000108.
28. Castillo DT, Chee CL, Nason E, et al. Group-delivered cognitive/exposure therapy for PTSD in women veterans: A randomized controlled trial. *Psychol Trauma*. 2016;8(3):404-412. doi:10.1037/tra0000111.
29. Chard KM. An evaluation of cognitive processing therapy for the treatment of posttraumatic stress disorder related to childhood sexual abuse. *J Consult Clin Psychol*. 2005;73(5):965-971. doi:10.1037/0022-006X.73.5.965.
30. Classen CC, Hughes L, Clark C, Hill Mohammed B, Woods P, Beckett B. A Pilot RCT of A Body-Oriented Group Therapy For Complex Trauma Survivors: An Adaptation of Sensorimotor Psychotherapy. *J Trauma Dissociation*. 2021;22(1):52-68. doi:10.1080/15299732.2020.1760173.
31. Cloitre M, Koenen KC, Cohen LR, Han H. Skills training in affective and interpersonal regulation followed by exposure: A phase-based treatment for PTSD related to childhood abuse. *J Consult Clin Psychol*. 2002;70(5):1067-1074. doi:10.1037//0022-006X.70.5.1067.
32. Cloitre M, Stovall-McClough KC, Nooner K, et al. Treatment for PTSD related to childhood abuse: a randomized controlled trial. *Am J Psychiatry*. 2010;167(8):915-924. doi:10.1176/appi.ajp.2010.09081247.
33. Cottraux J, Note I, Yao SN, et al. Randomized controlled comparison of cognitive behavior therapy with Rogerian supportive therapy in chronic post-traumatic stress disorder: a 2-year follow-up. *Psychother Psychosom*. 2008;77(2):101-110. doi:10.1159/000112887.
34. Davis LW, Schmid AA, Daggy JK, et al. Symptoms improve after a yoga program designed for PTSD in a randomized controlled trial with veterans and civilians. *Psychol Trauma*. 2020;12(8):904-912. doi:10.1037/tra0000564.
35. Devilly GJ, Spence SH, Rapee RM. Statistical and reliable change with eye movement desensitization and reprocessing: Treating trauma within a veteran population. *Behav Ther*. 1998;29(3):435-455. doi:10.1016/S0005-7894(98)80042-7.
36. Devilly GJ, Spence SH. The Relative Efficacy and Treatment Distress of EMDR and a Cognitive-Behavior Trauma Treatment Protocol in the Amelioration of Posttraumatic Stress Disorder. *J Anxiety Disord*. 1999;13(1-2):131-157. doi:10.1016/s0887-6185(98)00044-9.
37. Dorrepaal E, Thomaes K, Smit JH, et al. Stabilizing group treatment for complex posttraumatic stress disorder related to child abuse based on psychoeducation and cognitive behavioural therapy: a multisite randomized controlled trial. *Psychother Psychosom*. 2012;81(4):217-225. doi:10.1159/000335044.
38. Duffy M, Gillespie K, Clark DM. Post-traumatic stress disorder in the context of terrorism and other civil conflict in Northern Ireland: randomised controlled trial. *BMJ*. 2007;334(7604):1147. doi:10.1136/bmj.39021.846852.BE.
39. Dunne RL, Kenardy J, Sterling M. A randomized controlled trial of cognitive-behavioral therapy for the treatment of PTSD in the context of chronic whiplash. *Clinical Journal of Pain*. 2012;28(9):755-765. doi:10.1097/AJP.0b013e318243e16b.
40. Echeburúa E, Corral P, Zubizarreta I, Sarasua B. Psychological treatment of chronic posttraumatic stress disorder in victims of sexual aggression. *Behavior Modification*. 1997;21(4):433-456. doi:10.1177/01454455970214003.
41. Ehlers A, Clark DM, Hackmann A, et al. A randomized controlled trial of cognitive therapy, a self-help booklet, and repeated assessments as early interventions for posttraumatic stress disorder. *Arch Gen Psychiatry*. 2003;60(10):1024-1032. doi:10.1001/archpsyc.60.10.1024.
42. Ehlers A, Clark DM, Hackmann A, McManus F, Fennell M. Cognitive therapy for post-traumatic stress disorder: development and evaluation. *Behav Res Ther*. 2005;43(4):413-431. doi:10.1016/j.brat.2004.03.006.
43. Ehlers A, Hackmann A, Grey N, et al. A randomized controlled trial of 7-day intensive and standard weekly cognitive therapy for PTSD and emotion-focused supportive therapy. *Am J Psychiatry*. 2014;171(3):294-304. doi:10.1176/appi.ajp.2013.13040552.
44. Engel CC, Litz B, Magruder KM, et al. Delivery of self training and education for stressful situations (DESTRESS-PC): a randomized trial of nurse assisted online self-management for PTSD in primary care. *General Hospital Psychiatry*. 2015;37(4):323-328. doi:10.1016/j.genhosppsych.2015.04.007.
45. Ertl, V, Pfeiffer, A, Schauer, E, Elbert, T, Neuner, F. Community-implemented trauma therapy for former child soldiers in Northern Uganda: a randomized controlled trial. *JAMA*. 2011;306(5):503-512.
46. Falsetti SA, Resnick HS, Davis JL. Multiple channel exposure therapy for women with PTSD and comorbid panic attacks. *Cognitive Behaviour Therapy*. 2008;37(2):117-130. doi:10.1080/16506070801969088.
47. Fecteau G, Nicki R. Cognitive behavioural treatment of post traumatic stress disorder after motor vehicle accident. *Behav Cogn Psychother*. 1999;27(3):201-214. doi:10.1017/S135246589927302X.
48. Foa EB, Rothbaum BO, Riggs DS, Murdock TB. Treatment of posttraumatic stress disorder in rape victims: A comparison between cognitive-behavioral procedures and counseling. *J Consult Clin Psychol*. 1991;59(5):715-723. doi:10.1037//0022-006x.59.5.715.
49. Foa EB, Dancu CV, Hembree EA, Jaycox LH, Meadows EA, Street GP. A comparison of exposure therapy, stress inoculation training, and their combination for reducing posttraumatic stress disorder in female assault victims. *J Consult Clin Psychol*. 1999;67(2):194-200. doi:10.1037//0022-006x.67.2.194.
50. Foa EB, Hembree EA, Cahill SP, et al. Randomized trial of prolonged exposure for posttraumatic stress disorder with and without cognitive restructuring: outcome at academic and community clinics. *J Consult Clin Psychol*. 2005;73(5):953-964. doi:10.1037/0022-006X.73.5.953.
51. Foa EB, McLean CP, Zang Y, et al. Effect of Prolonged Exposure Therapy Delivered Over 2 Weeks vs 8 Weeks vs Present-Centered Therapy on PTSD Symptom Severity in Military Personnel: A Randomized Clinical Trial. *JAMA*. 2018;319(4):354-364. doi:10.1001/jama.2017.21242.
52. Fonzo GA, Goodkind MS, Oathes DJ, et al. Selective Effects of Psychotherapy on Frontopolar Cortical Function in PTSD. *Am J Psychiatry*. 2017;174(12):1175-1184. doi:10.1176/appi.ajp.2017.16091073.
53. Forbes D, Lloyd D, Nixon RDV, et al. A multisite randomized controlled effectiveness trial of cognitive processing therapy for military-related posttraumatic stress disorder. *J Anxiety Disord*. 2012;26(3):442-452. doi:10.1016/j.janxdis.2012.01.006.
54. Ford JD, Steinberg KL, Zhang W. A randomized clinical trial comparing affect regulation and social problem-solving psychotherapies for mothers with victimization-related PTSD. *Behav Ther*. 2011;42(4):560-578. doi:10.1016/j.beth.2010.12.005.
55. Ford JD, Chang R, Levine J, Zhang W. Randomized clinical trial comparing affect regulation and supportive group therapies for victimization-related PTSD with incarcerated women. *Behav Ther*. 2013;44(2):262-276. doi:10.1016/j.beth.2012.10.003.
56. Galovski TE, Blain LM, Mott JM, Elwood L, Houle T. Manualized therapy for PTSD: flexing the structure of cognitive processing therapy. *J Consult Clin Psychol*. 2012;80(6):968-981. doi:10.1037/a0030600.
57. Gersons BP, Carlier IV, Lamberts RD, van der Kolk BA. Randomized clinical trial of brief eclectic psychotherapy for police officers with posttraumatic stress disorder. *J Trauma Stress*. 2000;13(2):333-347. doi:10.1023/A:1007793803627.
58. Ghafoori B, Hansen MC, Garibay E, Korosteleva O. Feasibility of Training Frontline Therapists in Prolonged Exposure: A Randomized Controlled Pilot Study of Treatment of Complex Trauma in Diverse Victims of Crime and Violence. *J Nerv Ment Dis*. 2017;205(4):283-293. doi:10.1097/NMD.0000000000000659.
59. Goldstein LA, Mehling WE, Metzler TJ, et al. Veterans Group Exercise: A randomized pilot trial of an Integrative Exercise program for veterans with posttraumatic stress. *J Affect Disord*. 2018;227:345-352. doi:10.1016/j.jad.2017.11.002.
60. Gray R, Budden-Potts D, Bourke F. Reconsolidation of Traumatic Memories for PTSD: A randomized controlled trial of 74 male veterans. *Psychother Res*. 2019;29(5):621-639. doi:10.1080/10503307.2017.1408973.
61. Gray RM, Budden-Potts D, Schwall RJ, Bourke FF. An open-label, randomized controlled trial of the reconsolidation of traumatic memories protocol (RTM) in military women. *Psychol Trauma: Theory Res Pract Policy*. 2020. doi:10.1037/tra0000986.
62. Heffner, KL, Crean, HF, & Kemp, JE (2016). Meditation programs for veterans with posttraumatic stress disorder: Aggregate findings from a multi-site evaluation. *Psychol Trauma*. 2016;8(3):365-374.
63. Hensel-Dittmann D, Schauer M, Ruf M, et al. Treatment of traumatized victims of war and torture: a randomized controlled comparison of narrative exposure therapy and stress inoculation training. *Psychother Psychosom*. 2011;80(6):345-352. doi:10.1159/000327253.
64. Hinton DE, Hofmann SG, Rivera E, Otto MW, Pollack MH. Culturally adapted CBT (CA-CBT) for Latino women with treatment-resistant PTSD: a pilot study comparing CA-CBT to applied muscle relaxation. *Behav Res Ther*. 2011;49(4):275-280. doi:10.1016/j.brat.2011.01.005.
65. Hollifield M, Sinclair-Lian N, Warner TD, Hammerschlag R. Acupuncture for posttraumatic stress disorder: a randomized controlled pilot trial. *J Nerv Ment Dis*. 2007;195(6):504-513. doi:10.1097/NMD.0b013e31803044f8.
66. Ivarsson D, Blom M, Hesser H, et al. Guided internet-delivered cognitive behavior therapy for post-traumatic stress disorder: A randomized controlled trial. *Internet Interventions*. 2014;1(1):33-40. doi:10.1016/j.invent.2014.03.002.
67. Jacob N, Neuner F, Maedl A, Schaal S, Elbert T. Dissemination of psychotherapy for trauma spectrum disorders in postconflict settings: a randomized controlled trial in Rwanda. *Psychother Psychosom*. 2014;83(6):354-363. doi:10.1159/000365114.
68. Jalal B, Kruger Q, Hinton DE. Culturally adapted CBT (CA-CBT) for traumatised indigenous South Africans (Sepedi): a randomised pilot trial comparing CA-CBT to applied muscle relaxation. *Intervention*. 2020;(18):61-65. https://www.interventionjournal.org/text.asp?2020/18/1/61/285315.
69. Jensen JA. An investigation of eye movement desensitization and reprocessing (EMD/R) as a treatment for posttraumatic stress disorder (PTSD) symptoms of Vietnam combat veterans. *Behav Ther*. 1994;25(2):311-325. doi:10.1016/S0005-7894(05)80290-4.
70. Johnson DM, Zlotnick C, Perez S. Cognitive behavioral treatment of PTSD in residents of battered women's shelters: results of a randomized clinical trial. *J Consult Clin Psychol*. 2011;79(4):542-551. doi:10.1037/a0023822.
71. Johnson DM, Johnson NL, Perez SK, Palmieri PA, Zlotnick C. Comparison of Adding Treatment of PTSD During and After Shelter Stay to Standard Care in Residents of Battered Women's Shelters: Results of a Randomized Clinical Trial. *J Trauma Stress*. 2016;29(4):365-373. doi:10.1002/jts.22117.
72. Johnson DM, Palmieri PA, Zlotnick C, et al. A Randomized Controlled Trial Comparing HOPE Treatment and Present-Centered Therapy in Women Residing in Shelter with PTSD from Intimate Partner Violence. *Psychol Women Q*. 2020;44(4):539-553. doi:10.1177/0361684320953120.
73. Karatzias T, Power K, Brown K, et al. A controlled comparison of the effectiveness and efficiency of two psychological therapies for posttraumatic stress disorder: eye movement desensitization and reprocessing vs. emotional freedom techniques. *J Nerv Ment Dis*. 2011;199(6):372-378. doi:10.1097/NMD.0b013e31821cd262.
74. Keane TM, Fairbank JA, Caddell JM, Zimering RT. Implosive (flooding) therapy reduces symptoms of PTSD in Vietnam combat veterans. *Behav Ther*. 1989;20(2):245-260. doi:10.1016/S0005-7894(89)80072-3.
75. Kearney DJ, Malte CA, Storms M, Simpson TL. Loving-Kindness Meditation vs Cognitive Processing Therapy for Posttraumatic Stress Disorder Among Veterans: A Randomized Clinical Trial. *JAMA Netw Open*. 2021;4(4):e216604. doi:10.1001/jamanetworkopen.2021.6604.
76. Kelly U, Haywood T, Segell E, Higgins M. Trauma-sensitive yoga for post-traumatic stress disorder in women veterans who experienced military sexual trauma: interim results from a randomized controlled trial. *J Altern Complement Med*. 2021;27(S1):S45-S59.
77. Kent M, Davis MC, Stark SL, Stewart LA. A resilience-oriented treatment for posttraumatic stress disorder: results of a preliminary randomized clinical trial. *J Trauma Stress*. 2011;24(5):591-595. doi:10.1002/jts.20685.
78. Krakow B, Hollifield M, Schrader R, et al. A controlled study of imagery rehearsal for chronic nightmares in sexual assault survivors with PTSD: a preliminary report. *J Trauma Stress*. 2000;13(4):589-609. doi:10.1023/A:1007854015481.
79. Krakow B, Hollifield M, Johnston L, et al. Imagery rehearsal therapy for chronic nightmares in sexual assault survivors with posttraumatic stress disorder: a randomized controlled trial. *JAMA*. 2001;286(5):537-545. doi:10.1001/jama.286.5.537.
80. Krupnick JL, Green BL, Stockton P, Miranda J, Krause E, Mete M. Group interpersonal psychotherapy for low-income women with posttraumatic stress disorder. *Psychother Res*. 2008;18(5):497-507. doi:10.1080/10503300802183678.
81. Kubany ES, Hill EE, Owens JA. Cognitive trauma therapy for battered women with PTSD: preliminary findings. *J Trauma Stress*. 2003;16(1):81-91. doi:10.1023/A:1022019629803.
82. Kubany ES, Hill EE, Owens JA, et al. Cognitive trauma therapy for battered women with PTSD (CTT-BW). *J Consult Clin Psychol*. 2004;72(1):3-18. doi:10.1037/0022-006X.72.1.3.
83. Lang AJ, Malaktaris AL, Casmar P, et al. Compassion Meditation for Posttraumatic Stress Disorder in Veterans: A Randomized Proof of Concept Study. *J Trauma Stress*. 2019;32(2):299-309. doi:10.1002/jts.22397.
84. Langkaas TF, Hoffart A, Øktedalen T, Ulvenes PG, Hembree EA, Smucker M. Exposure and non-fear emotions: A randomized controlled study of exposure-based and rescripting-based imagery in PTSD treatment. *Behav Res Ther*. 2017;97:33-42. doi:10.1016/j.brat.2017.06.007.
85. Latif M, Husain MI, Gul M, et al. Culturally adapted trauma-focused CBT-based guided self-help (CatCBT GSH) for female victims of domestic violence in Pakistan: feasibility randomized controlled trial. *Behav Cogn Psychother*. 2021;49(1):50-61. doi:10.1017/S1352465820000685.
86. Laugharne J, Kullack C, Lee CW, et al. Amygdala Volumetric Change Following Psychotherapy for Posttraumatic Stress Disorder. *Journal of Neuropsychiatry and Clinical Neurosciences*. 2016;28(4):312-318. doi:10.1176/appi.neuropsych.16010006.
87. Lee C, Gavriel H, Drummond P, Richards J, Greenwald R. Treatment of PTSD: stress inoculation training with prolonged exposure compared to EMDR. *J Clin Psychol*. 2002;58(9):1071-1089. doi:10.1002/jclp.10039.
88. Lehavot K, Millard SP, Thomas RM, et al. A randomized trial of an online, coach-assisted self-management PTSD intervention tailored for women veterans. *J Consult Clin Psychol*. 2021;89(2):134-142. doi:10.1037/ccp0000556.
89. Lely JCG, Knipscheer JW, Moerbeek M, Heide FJJ ter, van den Bout J, Kleber RJ. Randomised controlled trial comparing narrative exposure therapy with present-centred therapy for older patients with post-traumatic stress disorder. *Br J Psychiatry*. 2019;214(6):369-377. doi:10.1192/bjp.2019.59.
90. Lewis CE, Farewell D, Groves V, et al. Internet-based guided self-help for posttraumatic stress disorder (PTSD): Randomized controlled trial. *Depress Anxiety*. 2017;34(6):555-565. doi:10.1002/da.22645.
91. Lindauer RJL, Gersons BPR, van Meijel EPM, et al. Effects of brief eclectic psychotherapy in patients with posttraumatic stress disorder: randomized clinical trial. *J Trauma Stress*. 2005;18(3):205-212. doi:10.1002/jts.20029.
92. Littleton H, Grills AE, Kline KD, Schoemann AM, Dodd JC. The From Survivor to Thriver program: RCT of an online therapist-facilitated program for rape-related PTSD. *J Anxiety Disord*. 2016;43:41-51.
93. Litz BT, Engel CC, Bryant RA, Papa A. A randomized, controlled proof-of-concept trial of an Internet-based, therapist-assisted self-management treatment for posttraumatic stress disorder. *Am J Psychiatry*. 2007;164(11):1676-1683. doi:10.1176/appi.ajp.2007.06122057.
94. Litz BT, Rusowicz-Orazem L, Doros G, et al. Adaptive disclosure, a combat-specific PTSD treatment, versus cognitive-processing therapy, in deployed marines and sailors: A randomized controlled non-inferiority trial. *Psychiatry Res*. 2021;297:113761. doi:10.1016/j.psychres.2021.113761.
95. Maguen S, Burkman K, Madden E, et al. Impact of Killing in War: A Randomized, Controlled Pilot Trial. *J Clin Psychol*. 2017;73(9):997-1012. doi:10.1002/jclp.22471.
96. Marcus SV, Marquis P, Sakai C. Controlled study of treatment of PTSD using EMDR in an HMO setting. *Psychotherapy: Theory, Research, Practice, Training*. 1997;34(3):307-315. doi:10.1037/h0087791.
97. Markowitz JC, Petkova E, Neria Y, et al. Is Exposure Necessary? A Randomized Clinical Trial of Interpersonal Psychotherapy for PTSD. *Am J Psychiatry*. 2015;172(5):430-440. doi:10.1176/appi.ajp.2014.14070908.
98. Marks I, Lovell K, Noshirvani H, Livanou M, Thrasher S. Treatment of posttraumatic stress disorder by exposure and/or cognitive restructuring: a controlled study. *Arch Gen Psychiatry*. 1998;55(4):317-325. doi:10.1001/archpsyc.55.4.317.
99. McDonagh A, Friedman M, McHugo G, et al. Randomized trial of cognitive-behavioral therapy for chronic posttraumatic stress disorder in adult female survivors of childhood sexual abuse. *J Consult Clin Psychol*. 2005;73(3):515-524. doi:10.1037/0022-006X.73.3.515.
100. McGuire Stanbury TM, Drummond PD, Laugharne J, Kullack C, Lee CW. Comparative Efficiency of EMDR and Prolonged Exposure in Treating Posttraumatic Stress Disorder: A Randomized Trial. *J EMDR Prac Res*. 2020;14(1):2-12. doi:10.1891/1933-3196.14.1.2.
101. McLean CP, Foa EB, Dondanville KA, et al. The effects of web-prolonged exposure among military personnel and veterans with posttraumatic stress disorder. *Psychol Trauma: Theory Res Pract Policy*. 2020. doi:10.1037/tra0000978.
102. Mitchell KS, Dick AM, DiMartino DM, et al. A pilot study of a randomized controlled trial of yoga as an intervention for PTSD symptoms in women. *J Trauma Stress*. 2014;27(2):121-128. doi:10.1002/jts.21903.
103. Monson CM, Schnurr PP, Resick PA, Friedman MJ, Young-Xu Y, Stevens SP. Cognitive processing therapy for veterans with military-related posttraumatic stress disorder. *J Consult Clin Psychol*. 2006;74(5):898-907. doi:10.1037/0022-006X.74.5.898.
104. Monson CM, Fredman SJ, Macdonald A, Pukay-Martin ND, Resick PA, Schnurr PP. Effect of cognitive-behavioral couple therapy for PTSD: a randomized controlled trial. *JAMA*. 2012;308(7):700-709. doi:10.1001/jama.2012.9307.
105. Morath J, Moreno-Villanueva M, Hamuni G, et al. Effects of psychotherapy on DNA strand break accumulation originating from traumatic stress. *Psychother Psychosom*. 2014;83(5):289-297. doi:10.1159/000362739.
106. Mueser KT, Rosenberg SD, Xie H, et al. A randomized controlled trial of cognitive-behavioral treatment for posttraumatic stress disorder in severe mental illness. *J Consult Clin Psychol*. 2008;76(2):259-271. doi:10.1037/0022-006X.76.2.259.
107. Nacasch N, Foa EB, Huppert JD, et al. Prolonged exposure therapy for combat- and terror-related posttraumatic stress disorder: a randomized control comparison with treatment as usual. *J Clin Psychiatry*. 2011;72(9):1174-1180. doi:10.4088/JCP.09m05682blu.
108. NCT00607815 PI: Chard, KM. A Comparison of Cognitive Processing Therapy (CPT) Versus Present Centered Therapy (PCT) for Veterans. Unpublished trial. Data retrieved from: https://clinicaltrials.gov/ct2/show/study/NCT00607815
109. Neuner F, Schauer M, Klaschik C, Karunakara U, Elbert T. A comparison of narrative exposure therapy, supportive counseling, and psychoeducation for treating posttraumatic stress disorder in an african refugee settlement. *J Consult Clin Psychol*. 2004;72(4):579-587. doi:10.1037/0022-006X.72.4.579.
110. Neuner F, Onyut PL, Ertl V, Odenwald M, Schauer E, Elbert T. Treatment of posttraumatic stress disorder by trained lay counselors in an African refugee settlement: a randomized controlled trial. *J Consult Clin Psychol*. 2008;76(4):686-694. doi:10.1037/0022-006X.76.4.686.
111. Neuner F, Kurreck S, Ruf M, Odenwald M, Elbert T, Schauer M. Can asylum-seekers with posttraumatic stress disorder be successfully treated? A randomized controlled pilot study. *Cogn Behav Ther*. 2010;39(2):81-91. doi:10.1080/16506070903121042.
112. Nidich S, Mills PJ, Rainforth M, et al. Non-trauma-focused meditation versus exposure therapy in veterans with post-traumatic stress disorder: A randomised controlled trial. *Lancet Psychiatry*. 2018;5(12):975-986. doi:10.1016/S2215-0366(18)30384-5.
113. Nijdam MJ, Gersons BPR, Reitsma JB, Jongh A, Olff M. Brief eclectic psychotherapy v. eye movement desensitisation and reprocessing therapy for post-traumatic stress disorder: randomised controlled trial. *Br J Psychiatry*. 2012;200(3):224-231. doi:10.1192/bjp.bp.111.099234.
114. Niles BL, Klunk-Gillis J, Ryngala DJ, Silberbogen AK, Paysnick A, Wolf EJ. Comparing mindfulness and psychoeducation treatments for combat-related PTSD using a telehealth approach. *Psychol Trauma*. 2012;4(5):538-547. doi:10.1037/a0026161.
115. Orang T, Ayoughi S, Moran JK, et al. The efficacy of narrative exposure therapy in a sample of Iranian women exposed to ongoing intimate partner violence-A randomized controlled trial. *Clin Psychol Psychother*. 2018;25(6):827-841. doi:10.1002/cpp.2318.
116. Pacella ML, Armelie A, Boarts J, et al. The impact of prolonged exposure on PTSD symptoms and associated psychopathology in people living with HIV: a randomized test of concept. *AIDS and Behavior*. 2012;16(5):1327-1340. doi:10.1007/s10461-011-0076-y.
117. Paunović N. Exposure Inhibition Therapy as a Treatment for Chronic Posttraumatic Stress Disorder: A Controlled Pilot Study. *Psychology*. 2011;02(06):605-614. doi:10.4236/psych.2011.26093.
118. Power K, McGoldrick T, Brown K, et al. A controlled comparison of eye movement desensitization and reprocessing versus exposure plus cognitive restructuring versus waiting list in the treatment of post-traumatic stress disorder. *Clin Psychol Psychother*. 2002;9(5):299-318. doi:10.1002/cpp.341.
119. Rauch SAM, King AP, Abelson J, et al. Biological and symptom changes in posttraumatic stress disorder treatment: a randomized clinical trial. *Depress Anxiety*. 2015;32(3):204-212. doi:10.1002/da.22331.
120. Ready DJ, Mascaro N, Wattenberg MS, Sylvers P, Worley V, Bradley-Davino B. A Controlled Study of Group-Based Exposure Therapy with Vietnam-Era Veterans. *Journal of Loss and Trauma*. 2018;23(6):439-457. doi:10.1080/15325024.2018.1485268.
121. Reger GM, Koenen-Woods P, Zetocha K, et al. Randomized controlled trial of prolonged exposure using imaginal exposure vs. virtual reality exposure in active duty soldiers with deployment-related posttraumatic stress disorder (PTSD). *J Consult Clin Psychol*. 2016;84(11):946-959. doi:10.1037/ccp0000134.
122. Resick PA, Nishith P, Weaver TL, Astin MC, Feuer CA. A comparison of cognitive-processing therapy with prolonged exposure and a waiting condition for the treatment of chronic posttraumatic stress disorder in female rape victims. *J Consult Clin Psychol*. 2002;70(4):867-879. doi:10.1037//0022-006X.70.4.867.
123. Resick PA, Wachen JS, Mintz J, et al. A randomized clinical trial of group cognitive processing therapy compared with group present-centered therapy for PTSD among active duty military personnel. *J Consult Clin Psychol*. 2015;83(6):1058-1068. doi:10.1037/ccp0000016.
124. Robjant K, Koebach A, Schmitt S, Chibashimba A, Carleial S, Elbert T. The treatment of posttraumatic stress symptoms and aggression in female former child soldiers using adapted Narrative Exposure therapy - a RCT in Eastern Democratic Republic of Congo. *Behav Res Ther*. 2019;123:103482. doi:10.1016/j.brat.2019.103482.
125. Rothbaum BO, Astin MC, Marsteller F. Prolonged Exposure versus Eye Movement Desensitization and Reprocessing (EMDR) for PTSD rape victims. *J Trauma Stress*. 2005;18(6):607-616. doi:10.1002/jts.20069.
126. Sautter FJ, Glynn SM, Cretu JB, Senturk D, Vaught AS. Efficacy of structured approach therapy in reducing PTSD in returning veterans: A randomized clinical trial. *Psychological Services*. 2015;12(3):199-212. doi:10.1037/ser0000032.
127. Schaal S, Elbert T, Neuner F. Narrative exposure therapy versus interpersonal psychotherapy. A pilot randomized controlled trial with Rwandan genocide orphans. *Psychother Psychosom*. 2009;78(5):298-306. doi:10.1159/000229768.
128. Scheck MM, Schaeffer JA, Gillette C. Brief psychological intervention with traumatized young women: the efficacy of eye movement desensitization and reprocessing. *J Trauma Stress*. 1998;11(1):25-44. doi:10.1023/A:1024400931106.
129. Schnurr PP, Friedman MJ, Foy DW, et al. Randomized trial of trauma-focused group therapy for posttraumatic stress disorder: results from a department of veterans affairs cooperative study. *Arch Gen Psychiatry*. 2003;60(5):481-489. doi:10.1001/archpsyc.60.5.481.
130. Schnurr PP, Friedman MJ, Engel CC, et al. Cognitive behavioral therapy for posttraumatic stress disorder in women: a randomized controlled trial. *JAMA*. 2007;297(8):820-830. doi:10.1001/jama.297.8.820.
131. Sloan, DM, Marx, BP, Greenberg, EM. A test of written emotional disclosure as an intervention for posttraumatic stress disorder. *Behav Res Ther*. 2011;49(4):299-304.
132. Sloan DM, Marx BP, Bovin MJ, Feinstein BA, Gallagher MW. Written exposure as an intervention for PTSD: a randomized clinical trial with motor vehicle accident survivors. *Behav Res Ther*. 2012;50(10):627-635. doi:10.1016/j.brat.2012.07.001.
133. Sloan DM, Unger W, Lee DJ, Beck JG. A Randomized Controlled Trial of Group Cognitive Behavioral Treatment for Veterans Diagnosed With Chronic Posttraumatic Stress Disorder. *J Trauma Stress*. 2018;31(6):886-898. doi:10.1002/jts.22338.
134. Spence, J, Titov, N, Dear, BF, et al. Randomized controlled trial of Internet‐delivered cognitive behavioral therapy for posttraumatic stress disorder. *Depress Anxiety*. 2011;28(7):541-550.
135. Stenmark H, Catani C, Neuner F, Elbert T, Holen A. Treating PTSD in refugees and asylum seekers within the general health care system. A randomized controlled multicenter study. *Behav Res Ther*. 2013;51(10):641-647. doi:10.1016/j.brat.2013.07.002.
136. Surís A, Link-Malcolm J, Chard K, Ahn C, North C. A randomized clinical trial of cognitive processing therapy for veterans with PTSD related to military sexual trauma. *J Trauma Stress*. 2013;26(1):28-37. doi:10.1002/jts.21765.
137. Taylor S, Thordarson DS, Maxfield L, Fedoroff IC, Lovell K, Ogrodniczuk J. Comparative efficacy, speed, and adverse effects of three PTSD treatments: exposure therapy, EMDR, and relaxation training. *J Consult Clin Psychol*. 2003;71(2):330-338. doi:10.1037/0022-006x.71.2.330.
138. terHeide FJJ, Mooren TM, van de Schoot R, Jongh A, Kleber RJ. Eye movement desensitisation and reprocessing therapy v. stabilisation as usual for refugees: randomised controlled trial. *Br J Psychiatry*. 2016;209(4):311-318. doi:10.1192/bjp.bp.115.167775.
139. Thorp SR, Glassman LH, Wells SY, et al. A randomized controlled trial of prolonged exposure therapy versus relaxation training for older veterans with military-related PTSD. *J Anxiety Disord*. 2019;64:45-54. doi:10.1016/j.janxdis.2019.02.003.
140. Tylee DS, Gray R, Glatt SJ, Bourke F. Evaluation of the reconsolidation of traumatic memories protocol for the treatment of PTSD: a randomized, wait-list-controlled trial. *Journal of Military, Veteran and Family Health*. 2017;3(1):21-33. doi:10.3138/jmvfh.4120.
141. van den Berg DPG, Bont PAJM de, van der Vleugel BM, et al. Prolonged exposure vs eye movement desensitization and reprocessing vs waiting list for posttraumatic stress disorder in patients with a psychotic disorder: a randomized clinical trial. *JAMA Psychiatry*. 2015;72(3):259-267. doi:10.1001/jamapsychiatry.2014.2637.
142. van der Kolk BA, Spinazzola J, Blaustein ME, et al. A randomized clinical trial of eye movement desensitization and reprocessing (EMDR), fluoxetine, and pill placebo in the treatment of posttraumatic stress disorder: treatment effects and long-term maintenance. *J Clin Psychiatry*. 2007;68(1):37-46. doi:10.4088/jcp.v68n0105.
143. van der Kolk BA, Stone L, West J, et al. Yoga as an adjunctive treatment for posttraumatic stress disorder: a randomized controlled trial. *J Clin Psychiatry*. 2014;75(6):e559-65. doi:10.4088/JCP.13m08561.
144. van Gelderen MJ, Nijdam MJ, Haagen JFG, Vermetten E. Interactive Motion-Assisted Exposure Therapy for Veterans with Treatment-Resistant Posttraumatic Stress Disorder: A Randomized Controlled Trial. *Psychother Psychosom*. 2020;89(4):215-227. doi:10.1159/000505977.
145. Vaughan K, Armstrong MS, Gold R, O'Connor N, Jenneke W, Tarrier N. A trial of eye movement desensitization compared to image habituation training and applied muscle relaxation in post-traumatic stress disorder. *J Behav Ther Exp Psychiatry*. 1994;25(4):283-291. doi:10.1016/0005-7916(94)90036-1.
146. Vera, M, Obén, A, Juarbe, D, Hernández, N, Kichic, R, Hembree, EA. A randomized clinical trial of prolonged exposure and applied relaxation for the treatment of Latinos with posttraumatic stress disorder. *J Trauma Stress*. 2022;35(2),:593-604.
147. Wagner AW, Jakupcak M, Kowalski HM, Bittinger JN, Golshan S. Behavioral Activation as a Treatment for Posttraumatic Stress Disorder Among Returning Veterans: A Randomized Trial. *Psychiatr Serv*. 2019;70(10):867-873. doi:10.1176/appi.ps.201800572.
148. Wahbeh H, Goodrich E, Goy E, Oken BS. Mechanistic Pathways of Mindfulness Meditation in Combat Veterans With Posttraumatic Stress Disorder. *J Clin Psychol*. 2016;72(4):365-383. doi:10.1002/jclp.22255.
149. Wells A, Walton D, Lovell K, Proctor D. Metacognitive Therapy Versus Prolonged Exposure in Adults with Chronic Post-traumatic Stress Disorder: A Parallel Randomized Controlled Trial. *Cogn Ther Res*. 2015;39(1):70-80. doi:10.1007/s10608-014-9636-6.
150. Wells A, Colbear JS. Treating posttraumatic stress disorder with metacognitive therapy: a preliminary controlled trial. *J Clin Psychol*. 2012;68(4):373-381. doi:10.1002/jclp.20871.
151. Yehuda R, Pratchett LC, Elmes MW, et al. Glucocorticoid-related predictors and correlates of post-traumatic stress disorder treatment response in combat veterans. *Interface Focus*. 2014;4(5):20140048. doi:10.1098/rsfs.2014.0048.
152. Yurtsever A, Konuk E, Akyüz T, et al. An Eye Movement Desensitization and Reprocessing (EMDR) Group Intervention for Syrian Refugees With Post-traumatic Stress Symptoms: Results of a Randomized Controlled Trial. *Front Psychol*. 2018;9:493. doi:10.3389/fpsyg.2018.00493.
153. Zang Y, Hunt N, Cox T. A randomised controlled pilot study: the effectiveness of narrative exposure therapy with adult survivors of the Sichuan earthquake. *BMC Psychiatry*. 2013;13:41. doi:10.1186/1471-244X-13-41.
154. Zang Y, Hunt N, Cox T. Adapting narrative exposure therapy for Chinese earthquake survivors: a pilot randomised controlled feasibility study. *BMC Psychiatry*. 2014;14:262. doi:10.1186/s12888-014-0262-3.
155. Zemestani, M, Mohammed, AF, Ismail, AA, Vujanovic, AA. A Pilot Randomized Clinical Trial of a Novel, Culturally Adapted, Trauma-Focused Cognitive-Behavioral Intervention for War-Related PTSD in Iraqi Women. *Behav Ther*. Accepted. [doi:10.1016/j.beth.2022.01.009](https://doi.org/10.1016/j.beth.2022.01.009)
156. Zlotnick C, Shea TM, Rosen K, et al. An affect-management group for women with posttraumatic stress disorder and histories of childhood sexual abuse. *J Trauma Stress*. 1997;10(3):425-436. doi:10.1023/a:1024841321156.
157. Zoellner LA, Telch M, Foa EB, et al. Enhancing Extinction Learning in Posttraumatic Stress Disorder With Brief Daily Imaginal Exposure and Methylene Blue: A Randomized Controlled Trial. *J Clin Psychiatry*. 2017;78(7):e782-e789. doi:10.4088/JCP.16m10936.

**Appendix H: Network graph for FU1 (i.e., ≤ 5 months follow-up)**

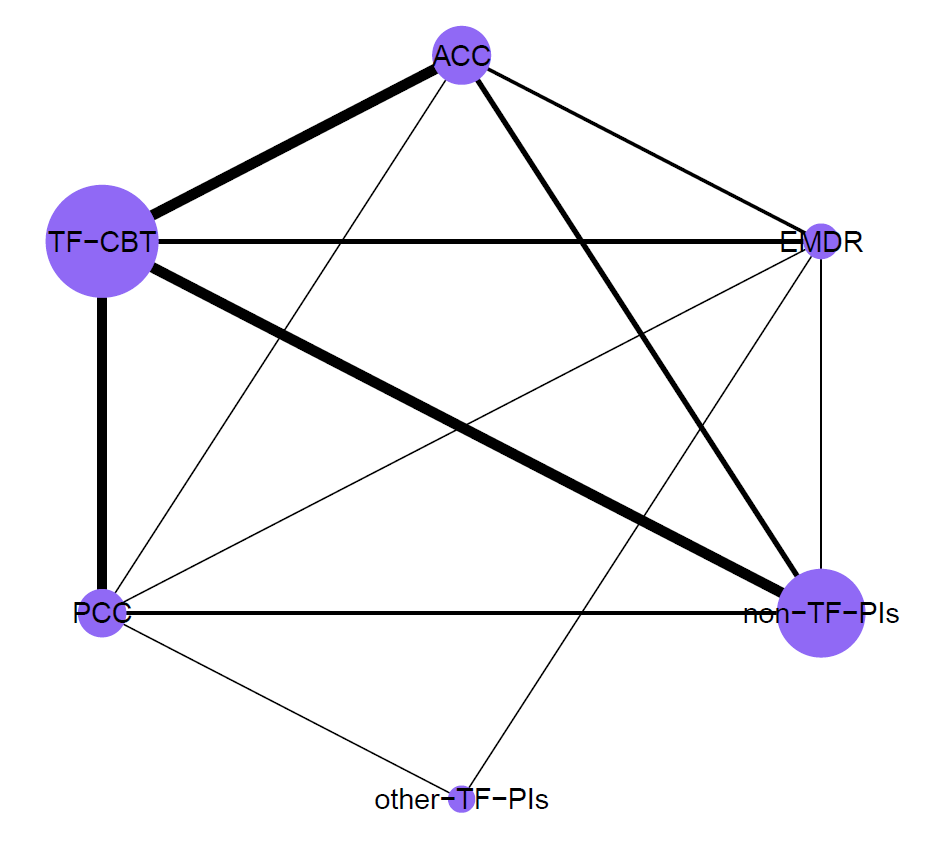

Nodes sizes are proportional to the number of included participants per dyad and the thickness of lines is proportional to the number of direct comparisons. ACC – active control conditions (e.g., treatment-as-usual), EMDR – eye movement desensitization and reprocessing, non-TF-PIs – non-trauma-focused psychological interventions, other-TF-PIs – other trauma-focused psychological interventions (i.e., other than TF-CBT and EMDR), PCC – passive control conditions (e.g., waitlist), TF-CBT – trauma-focused cognitive behaviour therapy

**Appendix I: Network Graph for FU2 (i.e., > 5 months follow-up)**

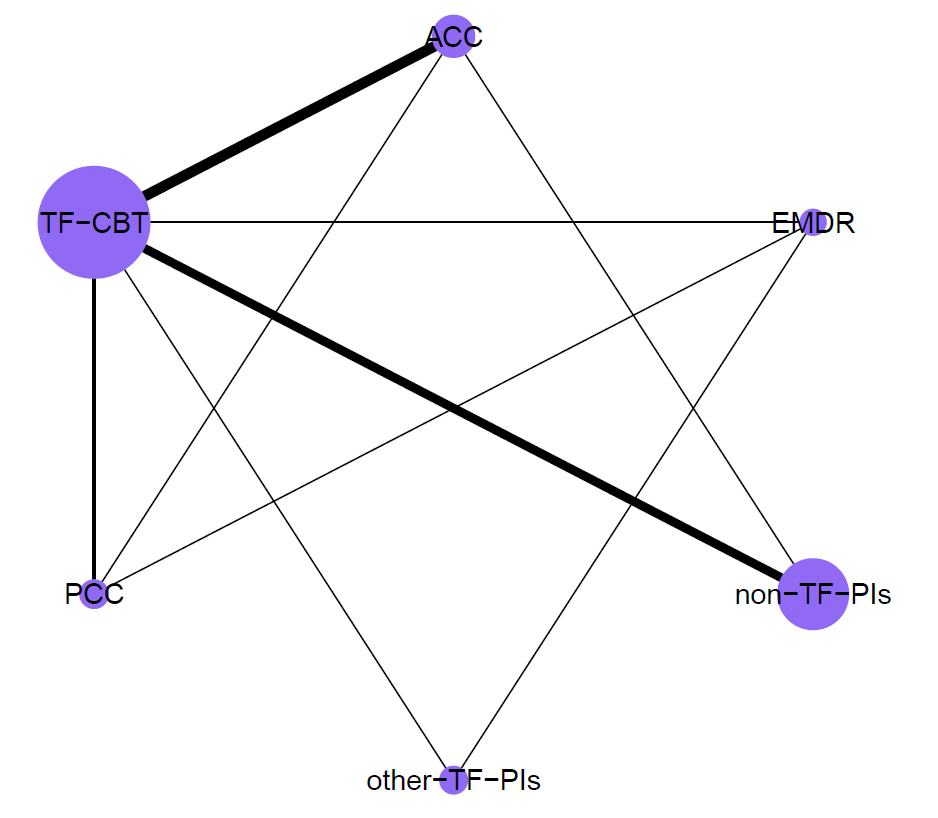

Nodes sizes are proportional to the number of included participants per dyad and the thickness of lines is proportional to the number of direct comparisons. ACC – active control conditions (e.g., treatment-as-usual), EMDR – eye movement desensitization and reprocessing, non-TF-PIs – non-trauma-focused psychological interventions, other-TF-PIs – other trauma-focused psychological interventions (i.e., other than TF-CBT and EMDR), PCC – passive control conditions (e.g., waitlist), TF-CBT – trauma-focused cognitive behaviour therapy

**Appendix J: Number of trials per dyad for all NMAs (incl. sensitivity analyses)**

| *Post-treatment Assessment (main analysis)* | | | | | |
| --- | --- | --- | --- | --- | --- |
|  | EMDR | other-TF-PIs | non-TF-PIs | ACC | PCC |
| TF-CBT | 10 (9#) | 2 | 25 | 31 | 53 (47#) |
| EMDR |  | 2 | 1 | 11 | 7 |
| other-TF-PIs |  |  | 2 | 1 | 6 |
| non-TF-PIs |  |  |  | 16 | 18 (17#) |
| ACC |  |  |  |  | 5 |
| *Post-treatment Assessment (sensitivity analysis – high quality trials only)* | | | | | |
|  | EMDR | other-TF-PIs | non-TF-PIs | ACC | PCC |
| TF-CBT | 2 | 2 | 11 | 13 | 22 (20#) |
| EMDR |  | 2 | 1 | 3 | 2 (1#) |
| other-TF-PIs |  |  | 0 | 0 | 2 |
| non-TF-PIs |  |  |  | 4 | 4 (3#) |
| ACC |  |  |  |  | 1 |
| *Post-treatment Assessment (sensitivity analysis – trials with individual f2f delivery only)* | | | | | |
|  | EMDR | other-TF-PIs | non-TF-PIs | ACC | PCC |
| TF-CBT | 10 (9#) | 2 | 19 | 27 | 45 (39#) |
| EMDR |  | 2 | 1 | 11 | 6 |
| other-TF-PIs |  |  | 2 | 0 | 4 |
| non-TF-PIs |  |  |  | 4 (3#) | 11 (10#) |
| ACC |  |  |  |  | 5 |
| *Follow-Up 1 Assessment (i.e., ≤ 5 months)* | | | | | |
|  | EMDR | other-TF-PIs | non-TF-PIs | ACC | PCC |
| TF-CBT | 4 | 0 | 17 | 18 | 14 (13#) |
| EMDR |  | 1 | 1 | 3 | 2 |
| other-TF-PIs |  |  | 0 | 0 | 1 |
| non-TF-PIs |  |  |  | 7 (6#) | 4 |
| ACC |  |  |  |  | 1 |
| *Follow-Up 1 Assessment (sensitivity analysis – trials with individual f2f delivery only)* | | | | | |
|  | EMDR | other-TF-PIs | non-TF-PIs | ACC | PCC |
| TF-CBT | 4 | 0 | 12 | 16 | 12 (11#) |
| EMDR |  | 1 | 1 | 3 | 1 |
| other-TF-PIs |  |  | 0 | 0 | 1 |
| non-TF-PIs |  |  |  | 1 | 1 |
| ACC |  |  |  |  | 1 |
| *Follow-Up 2 Assessment (i.e., > 5 months)* | | | | | |
|  | EMDR | other-TF-PIs | non-TF-PIs | ACC | PCC |
| TF-CBT | 2 | 2 | 13 | 15 (14#) | 5 |
| EMDR |  | 1 | 0 | 0 | 1 |
| other-TF-PIs |  |  | 0 | 0 | 0 |
| non-TF-PIs |  |  |  | 1 | 0 |
| ACC |  |  |  |  | 1 |

#number of trials per dyad for outlier-corrected analysis. ACC – active control conditions (e.g., treatment-as-usual), EMDR – eye movement desensitization and reprocessing, non-TF-PIs – non-trauma-focused psychological interventions, other-TF-PIs – other trauma-focused psychological interventions (i.e., other than TF-CBT and EMDR), PCC – passive control conditions (e.g., waitlist), TF-CBT – trauma-focused cognitive behaviour therapy

**Appendix K: Bean plot for the distribution of participants’ sexes (i.e., % females)**

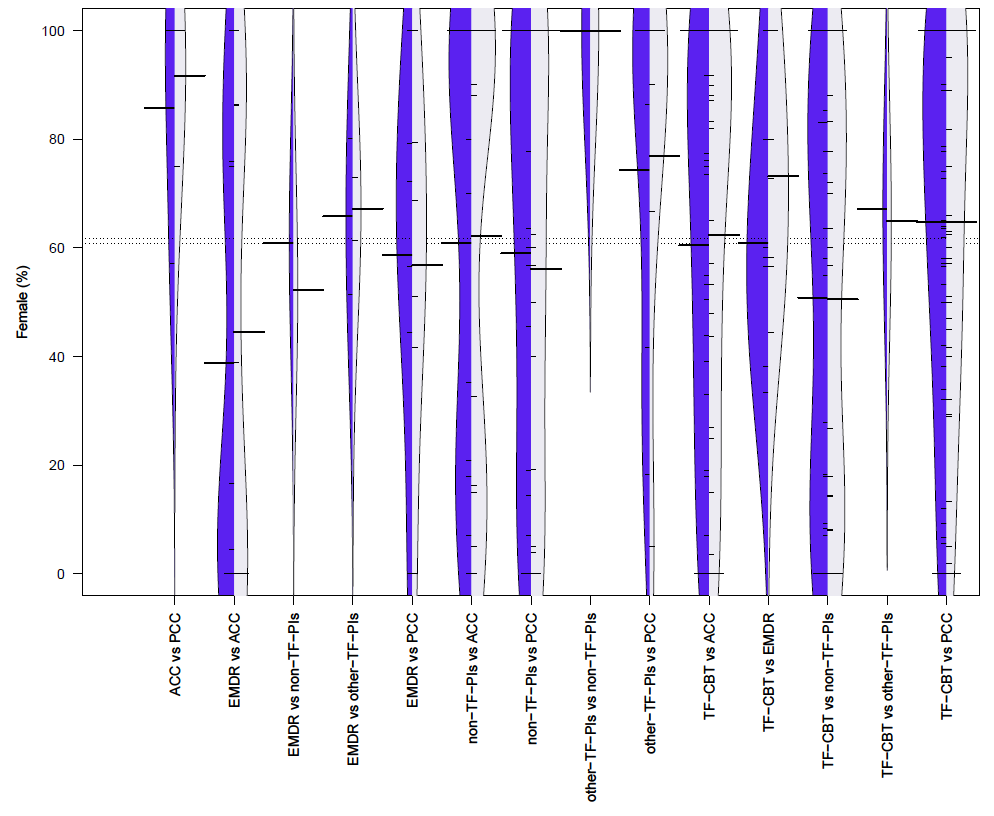

ACC – active control conditions (e.g., treatment-as-usual), EMDR – eye movement desensitization and reprocessing, non-TF-PIs – non-trauma-focused psychological interventions, other-TF-PIs – other trauma-focused psychological interventions (i.e., other than TF-CBT and EMDR), PCC – passive control conditions (e.g., waitlist), TF-CBT – trauma-focused cognitive behaviour therapy

**Appendix L: Comparison of participant and trial characteristics across comparison dyads**

|  | *100% PTSD* | | *Sample age* | | *% females* | | *Western* | | *HQ^a^* | | *utilized CAPS* | | *Individual delivery^b^* | | *Treatment length^c^* | |
| --- | --- | --- | --- | --- | --- | --- | --- | --- | --- | --- | --- | --- | --- | --- | --- | --- |
| *Comparison* | *k* | *%* | *mean* | *SD* | *mean* | *SD* | *k* | *%* | *k* | *%* | *k* | *%* | *k* | *%* | *mean* | *SD* |
| TF-CBT vs PCC | 54 | 96.43 | 38.17 | 7.15 | 64.21 | 31.79 | 48 | 85.71 | 23 | 41.07 | 27 | 48.21 | 47 | 83.93 | 817.33 | 456.29 |
| EMDR vs PCC | 6 | 85.71 | 36.72 | 4.02 | 58.81 | 31.61 | 5 | 71.43 | 2 | 28.57 | 3 | 42.86 | 6 | 85.71 | 615.71 | 232.01 |
| other-TF-PIs vs PCC | 3 | 50 | 35.38 | 3.84 | 75.70 | 35.94 | 6 | 100 | 2 | 33.33 | 2 | 33.33 | 4 | 66.67 | 590 | 261.30 |
| non-TF-PIs vs PCC | 13 | 72.22 | 42.19 | 8.94 | 57.52 | 38.36 | 16 | 88.89 | 4 | 22.22 | 9 | 50 | 11 | 61.11 | 1063.85 | 546.98 |
| TF-CBT vs ACC | 31 | 88.57 | 37.28 | 10.17 | 62.88 | 34.77 | 26 | 74.29 | 14 | 40 | 18 | 51.43 | 30 | 85.71 | 870.91 | 323.81 |
| EMDR vs ACC | 9 | 81.82 | 39.43 | 8.18 | 46.33 | 40.29 | 11 | 100 | 3 | 27.27 | 5 | 45.45 | 11 | 100 | 501 | 248.66 |
| other-TF-PIs vs ACC | 1 | 100 | 38.80 | 0 | n.r. | n.r. | 1 | 100 | 0 | 0 | 0 | 0 | 0 | 0 | 2400 | 0 |
| non-TF-PIs vs ACC | 15 | 93.75 | 42.82 | 10.17 | 56.09 | 41.57 | 15 | 93.75 | 4 | 25 | 10 | 62.50 | 4 | 25 | 798.93 | 431.61 |
| TF-CBT vs non-TF-PIs | 25 | 96.15 | 41.98 | 10.14 | 49.67 | 37.20 | 25 | 96.15 | 11 | 42.31 | 19 | 73.08 | 20 | 76.92 | 1368.64 | 1510.04 |
| EMDR vs non-TF-PIs | 1 | 100 | 40.60 | 0 | 56.50 | 0 | 1 | 100 | 1 | 100 | 1 | 100 | 1 | 100 | 480 | 0 |
| other-TF-PIs vs non-TF-PIs | 0 | 0 | 33.45 | 3.89 | 100 | 0 | 2 | 100 | 0 | 0 | 2 | 100 | 2 | 100 | 750 | 212.13 |
| TF-CBT vs other-TF-PIs | 2 | 100 | 40.57 | 6.55 | 62 | 5.66 | 2 | 100 | 2 | 100 | 0 | 0 | 2 | 100 | 1050 | 0 |
| EMDR vs other-TF-PIs | 2 | 100 | 38.17 | 0.52 | 66.60 | 14.42 | 2 | 100 | 2 | 100 | 1 | 50 | 2 | 100 | 1305 | 318.20 |
| TF-CBT vs EMDR | 9 | 90 | 39.38 | 5.80 | 67.63 | 19.11 | 10 | 100 | 2 | 20 | 5 | 50 | 10 | 100 | 760 | 274.07 |

*^a^*Meeting at least seven of eight trial quality criteria (based on Cuijpers et al., 2010). *^b^*Treatment delivered individually and face-to-face. *^c^*Total treatment length in minutes ([mean] number of sessions multiplied by [mean] session length). ACC – active control conditions (e.g., treatment-as-usual), CAPS – Clinician Administered PTSD Scale, EMDR – eye movement desensitization and reprocessing, non-TF-PIs – non-trauma-focused psychological interventions, other-TF-PIs – other trauma-focused psychological interventions (i.e., other than TF-CBT and EMDR), PCC – passive control conditions (e.g., waitlist), TF-CBT – trauma-focused cognitive behaviour therapy

**Appendix M: Forest plot for NMA on short-term efficacy with passive control conditions as reference group**

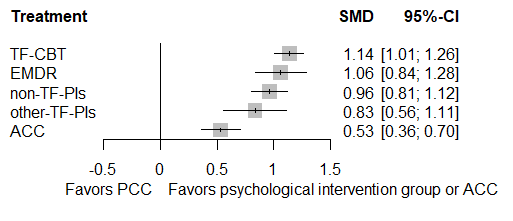

ACC – active control conditions (e.g., treatment-as-usual), EMDR – eye movement desensitization and reprocessing, non-TF-PIs – non-trauma-focused psychological interventions, other-TF-PIs – other trauma-focused psychological interventions (i.e., other than TF-CBT and EMDR), PCC – passive control conditions (e.g., waitlist), TF-CBT – trauma-focused cognitive behaviour therapy

**Appendix N: Forest plot for NMA on short-term efficacy with non-TF-PIs as reference group**

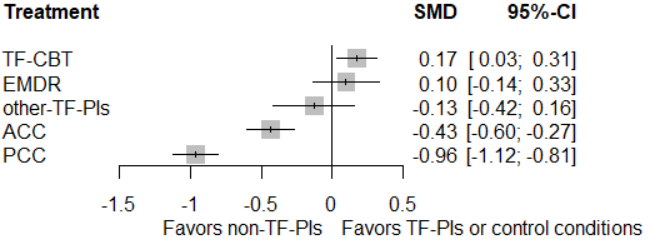

ACC – active control conditions (e.g., treatment-as-usual), EMDR – eye movement desensitization and reprocessing, non-TF-PIs – non-trauma-focused psychological interventions, other-TF-PIs – other trauma-focused psychological interventions (i.e., other than TF-CBT and EMDR), PCC – passive control conditions (e.g., waitlist), TF-CBT – trauma-focused cognitive behaviour therapy

**Appendix O: Funnel plot for NMA on short-term efficacy**

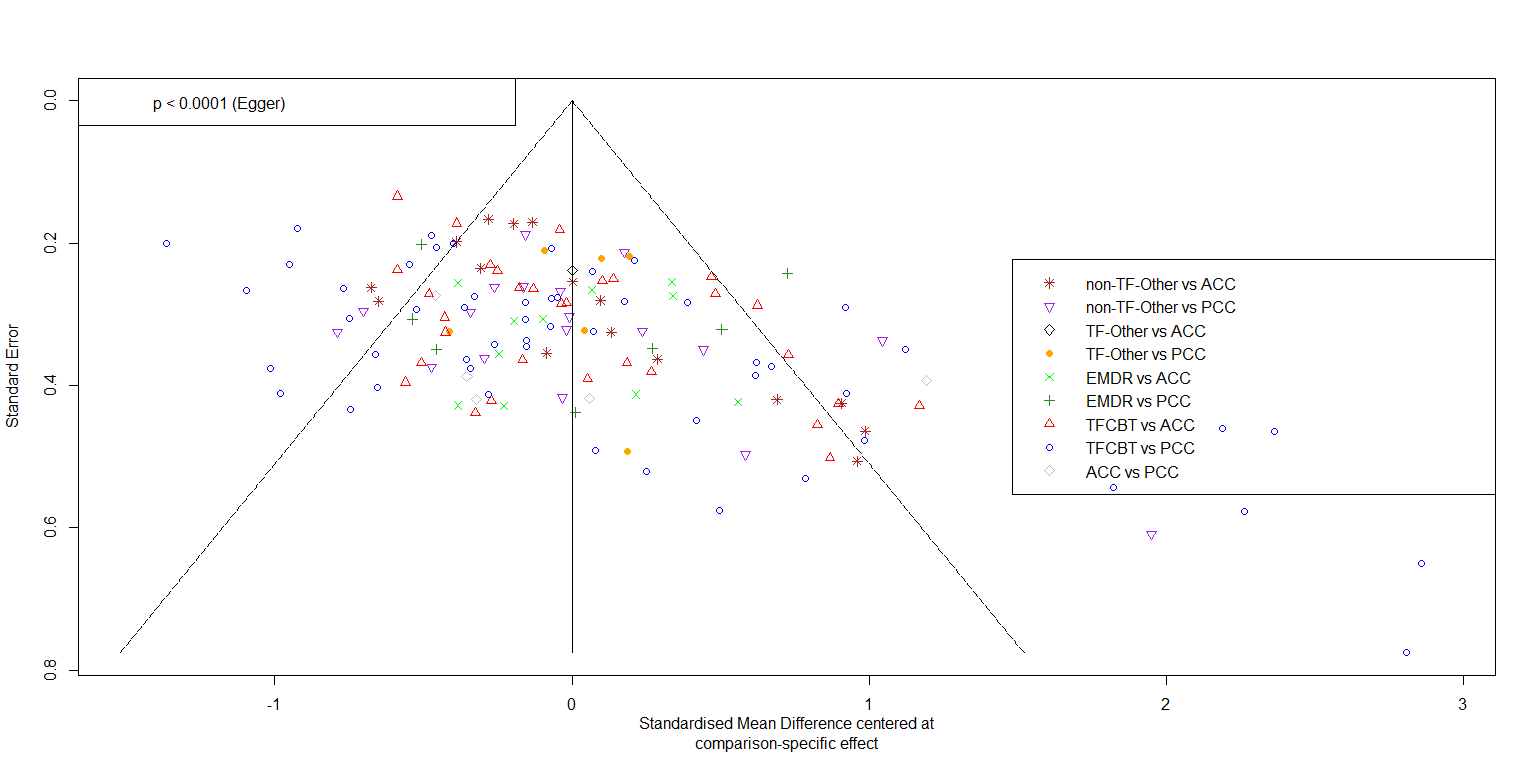

ACC – active control conditions (e.g., treatment-as-usual), EMDR – eye movement desensitization and reprocessing, non-TF-PIs – non-trauma-focused psychological interventions, other-TF-PIs – other trauma-focused psychological interventions (i.e., other than TF-CBT and EMDR), PCC – passive control conditions (e.g., waitlist), TF-CBT – trauma-focused cognitive behaviour therapy

**Appendix P: Netheat plot for NMA on short-term efficacy**

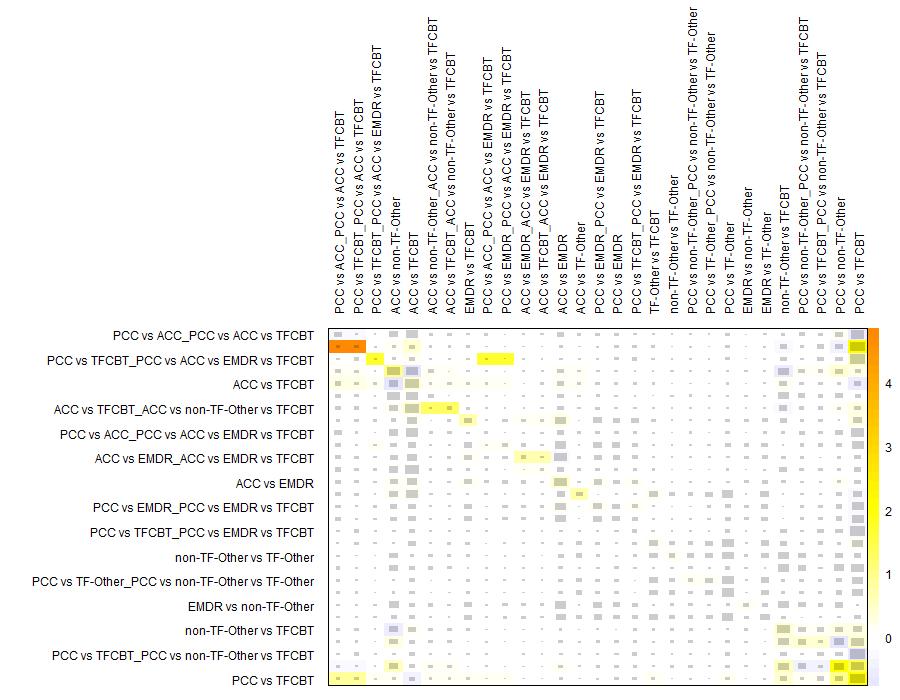

ACC – active control conditions (e.g., treatment-as-usual), EMDR – eye movement desensitization and reprocessing, non-TF-PIs – non-trauma-focused psychological interventions, other-TF-PIs – other trauma-focused psychological interventions (i.e., other than TF-CBT and EMDR), PCC – passive control conditions (e.g., waitlist), TF-CBT – trauma-focused cognitive behaviour therapy

**Appendix Q: Outlier-corrected comparative short-term efficacy of psychological interventions (PIs)**

| *Reference group PIs* | | | *k  (N)* | *g* | *95% CI* | *p* | | *I^2^ (τ^2^)* |
| --- | --- | --- | --- | --- | --- | --- | --- | --- |
|  | ***Outlier-corrected main analysis*** | | | | | | |  |
| relative to PCC | | TF-CBT# | 46 (2,642) | **1.00** | **0.88 – 1.11** | **< .001** | | 59.40 *** (0.10) |
|  |  | EMDR | 7 (396) | **0.93** | **0.74 – 1.13** | **< .001** | |  |
|  |  | other-TF-PIs | 6 (385) | **0.76** | **0.53 – 1.00** | **< .001** | |  |
|  |  | non-TF-PIs# | 17 (891) | **0.85** | **0.71 – 0.99** | **< .001** | |  |
|  | | ACC | 5 (154) | **0.42** | **0.27 – 0.57** | **< .001** | |  |
| relative to ACC | | TF-CBT | 31 (1,736) | **0.58** | **0.45 – 0.79** | **< .001** | |  |
|  |  | EMDR | 11 (449) | **0.51** | **0.32 – 0.70** | **< .001** | |  |
|  |  | other-TF-PIs | 1 (71) | **0.34** | **0.09 – 0.59** | **.008** | |  |
|  |  | non-TF-PIs | 16 (1,026) | **0.43** | **0.28 – 0.57** | **< .001** | |  |
| relative to EMDR | | TF-CBT# | 9 (338) | 0.06 | -0.12 – 0.25 | .512 | |  |
|  |  | other-TF-PIs | 2 (224) | -0.17 | -0.44 – 0.09 | .203 | |  |
|  |  | non-TF-PIs | 1 (46) | -0.09 | -0.29 – 0.12 | .408 | |  |
| relative to other-TF-PIs | | TF-CBT | 2 (204) | **0.24** | **-0.00 – 0.47** | **.050** | |  |
|  |  | non-TF-PIs | 2 (173) | 0.09 | -0.16 – 0.33 | .487 | |  |
| relative to non-TF-PIs | | TF-CBT | 25 (2,587) | **0.15** | **0.03 – 0.27** | **.016** | |  |
|  | ***Outlier-corrected sensitivity analysis: high quality trials only*** | | | | | | |  |
| relative to PCC | | TF-CBT# | 20 (1,467) | **0.83** | **0.66 – 1.00** | **< .001** | | 65.60 *** (0.10) |
|  |  | EMDR# | 1 (102) | **0.69** | **0.36 – 1.02** | **< .001** | |  |
|  |  | other-TF-PIs | 2 (66) | **0.64** | **0.27 – 1.00** | **< .001** | |  |
|  |  | non-TF-PIs# | 3 (267) | **0.69** | **0.45 – 0.93** | **< .001** | |  |
|  | | ACC | 1 (52) | **0.33** | **0.08 – 0.57** | **.009** | |  |
| relative to ACC | | TF-CBT | 13 (1,036) | **0.51** | **0.31 – 0.70** | **< .001** | |  |
|  |  | EMDR | 3 (162) | **0.36** | **0.04 – 0.68** | **.026** | |  |
|  |  | other-TF-PIs | 0 (0) | **0.31** | **-0.07 – 0.69** | **.113** | |  |
|  |  | non-TF-PIs | 4 (366) | **0.36** | **0.12 – 0.60** | **.003** | |  |
| relative to EMDR | | TF-CBT | 2 (128) | 0.14 | -0.16 – 0.45 | .359 | |  |
|  |  | other-TF-PIs | 2 (224) | -0.05 | -0.42 – 0.32 | .786 | |  |
|  |  | non-TF-PIs | 1 (46) | 0.00 | -0.34 – 0.34 | .993 | |  |
| relative to other-TF-PIs | | TF-CBT | 2 (204) | 0.20 | -0.15 – 0.55 | .273 | |  |
|  |  | non-TF-PIs | 0 (0) | 0.05 | -0.34 – 0.44 | .789 | |  |
| relative to non-TF-PIs | | TF-CBT | 11 (1,390) | 0.14 | -0.05 – 0.34 | .150 | |  |
| ***Outlier-corrected sensitivity analysis: face-to-face delivery of PIs only*** | | | | | | |  | |
| relative to PCC | | TF-CBT# | 39 (2,242) | **1.00** | **0.87 – 1.13** | **< .001** | | 60.00 ***  (0.11) |
|  |  | EMDR | 6 (349) | **0.99** | **0.77 – 1.20** | **< .001** | |  |
|  |  | other-TF-PIs | 4 (198) | **0.81** | **0.53 – 1.09** | **< .001** | |  |
|  |  | non-TF-PIs# | 10 (633) | **0.84** | **0.66 – 1.02** | **< .001** | |  |
|  | | ACC | 5 (154) | **0.50** | **0.32 – 0.68** | **< .001** | |  |
| relative to ACC | | TF-CBT | 27 (1,558) | **0.50** | **0.35 – 0.65** | **< .001** | |  |
|  |  | EMDR | 11 (449) | **0.49** | **0.28 – 0.69** | **< .001** | |  |
|  |  | other-TF-PIs | 0 (0) | **0.31** | **0.01 – 0.61** | **.046** | |  |
|  |  | non-TF-PIs# | 3 (260) | **0.34** | **0.13 – 0.54** | **.001** | |  |
| relative to EMDR | | TF-CBT# | 9 (338) | 0.02 | -0.18 – 0.22 | .855 | |  |
|  |  | other-TF-PIs | 2 (224) | -0.18 | -0.48 – 0.13 | .255 | |  |
|  |  | non-TF-PIs | 1 (46) | -0.15 | -0.39 – 0.09 | .215 | |  |
| relative to other-TF-PIs | | TF-CBT | 2 (204) | 0.19 | -0.08 – 0.47 | .172 | |  |
|  |  | non-TF-PIs | 2 (173) | 0.03 | -0.27 – 0.32 | .861 | |  |
| relative to non-TF-PIs | | TF-CBT | 19 (1,637) | **0.17** | **0.01 – 0.33** | **.039** | |  |

#indicates that ≥ 1 outlier(s) was(were) detected and deleted for the outlier-corrected analyses. Bold prints highlight significant differences. *** p < .001, ** p < .01, * p < .05, correspond to the respective Q-statistic. ACC – active control conditions (e.g., treatment-as-usual), EMDR – eye movement desensitization and reprocessing, k – number of trials for the given comparison, N – number of participants for the given comparison, non-TF-PIs – non-trauma-focused psychological interventions, other-TF-PIs – other trauma-focused psychological interventions (i.e., other than TF-CBT and EMDR), PCC – passive control conditions (e.g., waitlist), PIs – psychological interventions, SMD – standardized mean differences (i.e., Hedges’ g), TF-CBT – trauma-focused cognitive behaviour therapy

**Appendix R: Outlier-adjusted funnel plot for NMA on short-term efficacy**

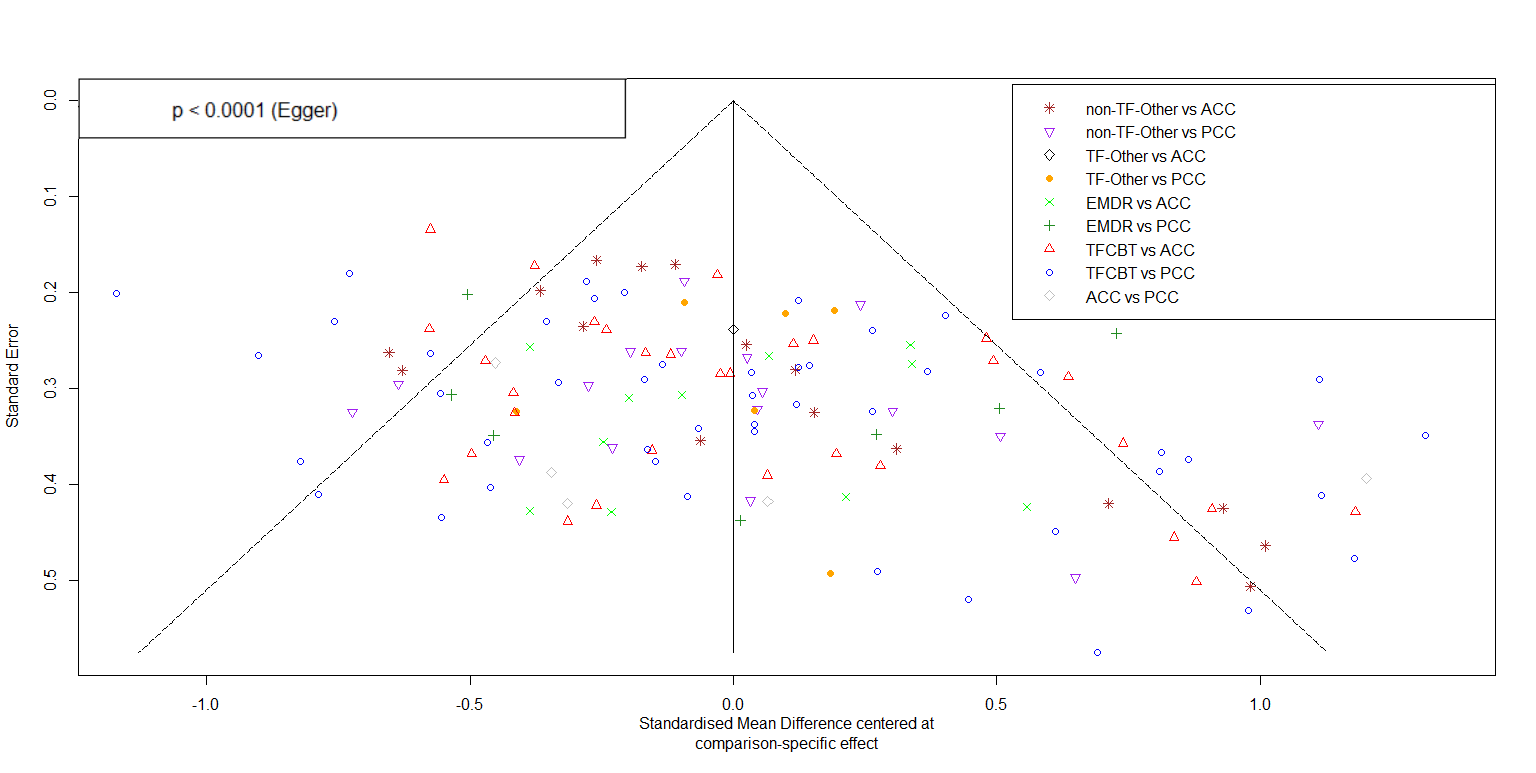

ACC – active control conditions (e.g., treatment-as-usual), EMDR – eye movement desensitization and reprocessing, non-TF-PIs – non-trauma-focused psychological interventions, other-TF-PIs – other trauma-focused psychological interventions (i.e., other than TF-CBT and EMDR), PCC – passive control conditions (e.g., waitlist), TF-CBT – trauma-focused cognitive behaviour therapy

**Appendix S: Forest plot for NMA at FU1 with passive control conditions as reference group**

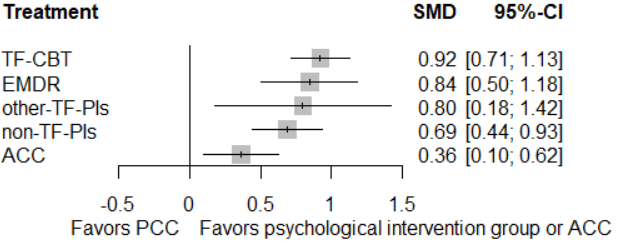

ACC – active control conditions (e.g., treatment-as-usual), EMDR – eye movement desensitization and reprocessing, FU1 – follow-up 1 (i.e., ≤ 5 months follow-up), non-TF-PIs – non-trauma-focused psychological interventions, other-TF-PIs – other trauma-focused psychological interventions (i.e., other than TF-CBT and EMDR), PCC – passive control conditions (e.g., waitlist), TF-CBT – trauma-focused cognitive behaviour therapy

**Appendix T: Forest plot for NMA at FU1 (i.e., ≤ 5 months follow-up) with non-TF-PIs as reference group**

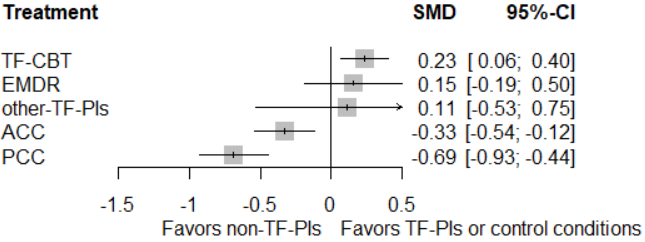

ACC – active control conditions (e.g., treatment-as-usual), EMDR – eye movement desensitization and reprocessing, FU1 – follow-up 1 (i.e., ≤ 5 months follow-up), non-TF-PIs – non-trauma-focused psychological interventions, other-TF-PIs – other trauma-focused psychological interventions (i.e., other than TF-CBT and EMDR), PCC – passive control conditions (e.g., waitlist), TF-CBT – trauma-focused cognitive behaviour therapy

**Appendix U: Funnel plot for NMA on efficacy at FU1 (i.e., ≤ 5 months follow-up)**

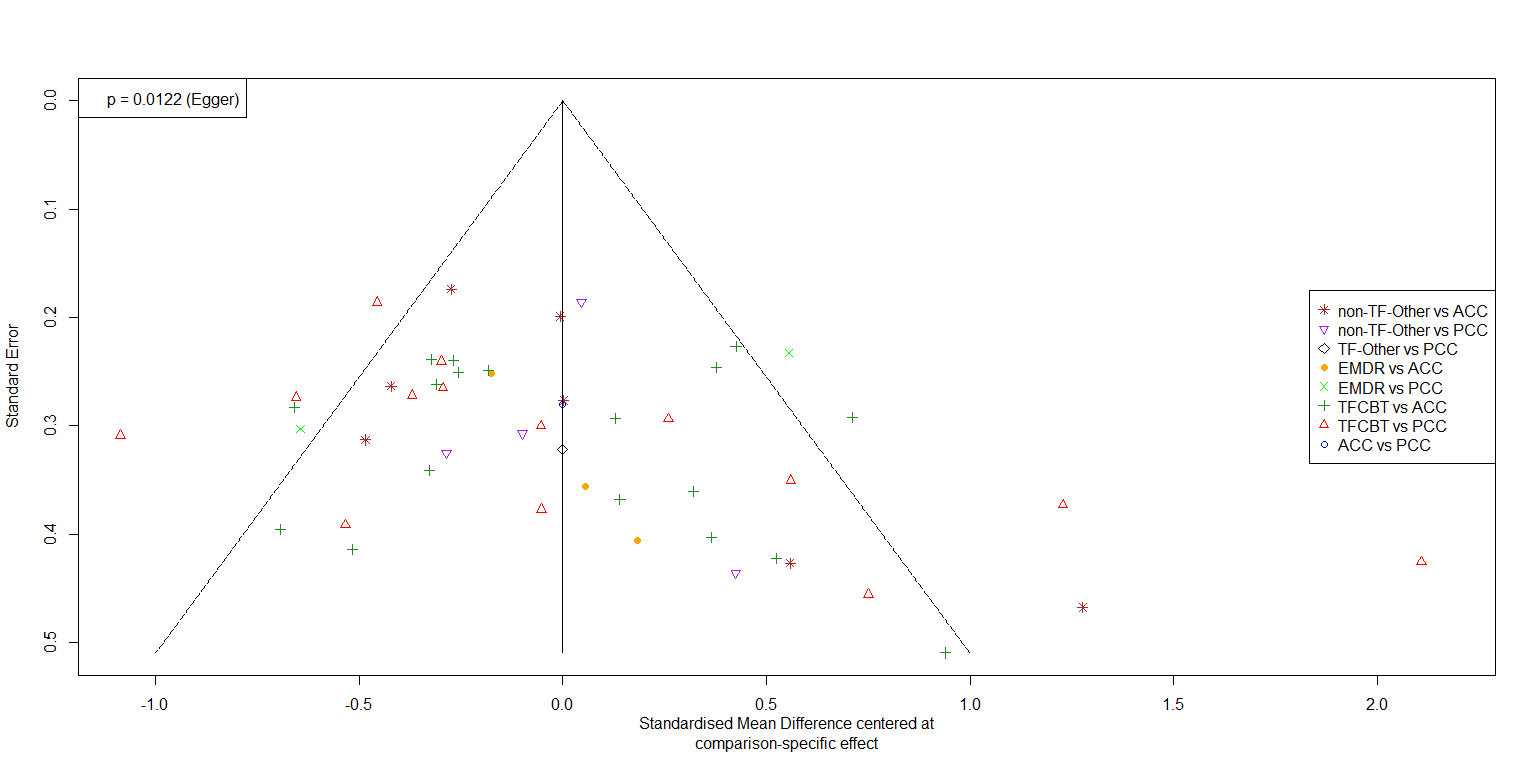

ACC – active control conditions (e.g., treatment-as-usual), EMDR – eye movement desensitization and reprocessing, FU1 – follow-up 1 (i.e., ≤ 5 months follow-up), non-TF-PIs – non-trauma-focused psychological interventions, other-TF-PIs – other trauma-focused psychological interventions (i.e., other than TF-CBT and EMDR), PCC – passive control conditions (e.g., waitlist), TF-CBT – trauma-focused cognitive behaviour therapy

**Appendix V. Outlier-adjusted funnel plot for NMA on efficacy at FU1 (i.e., ≤ 5 months follow-up)**

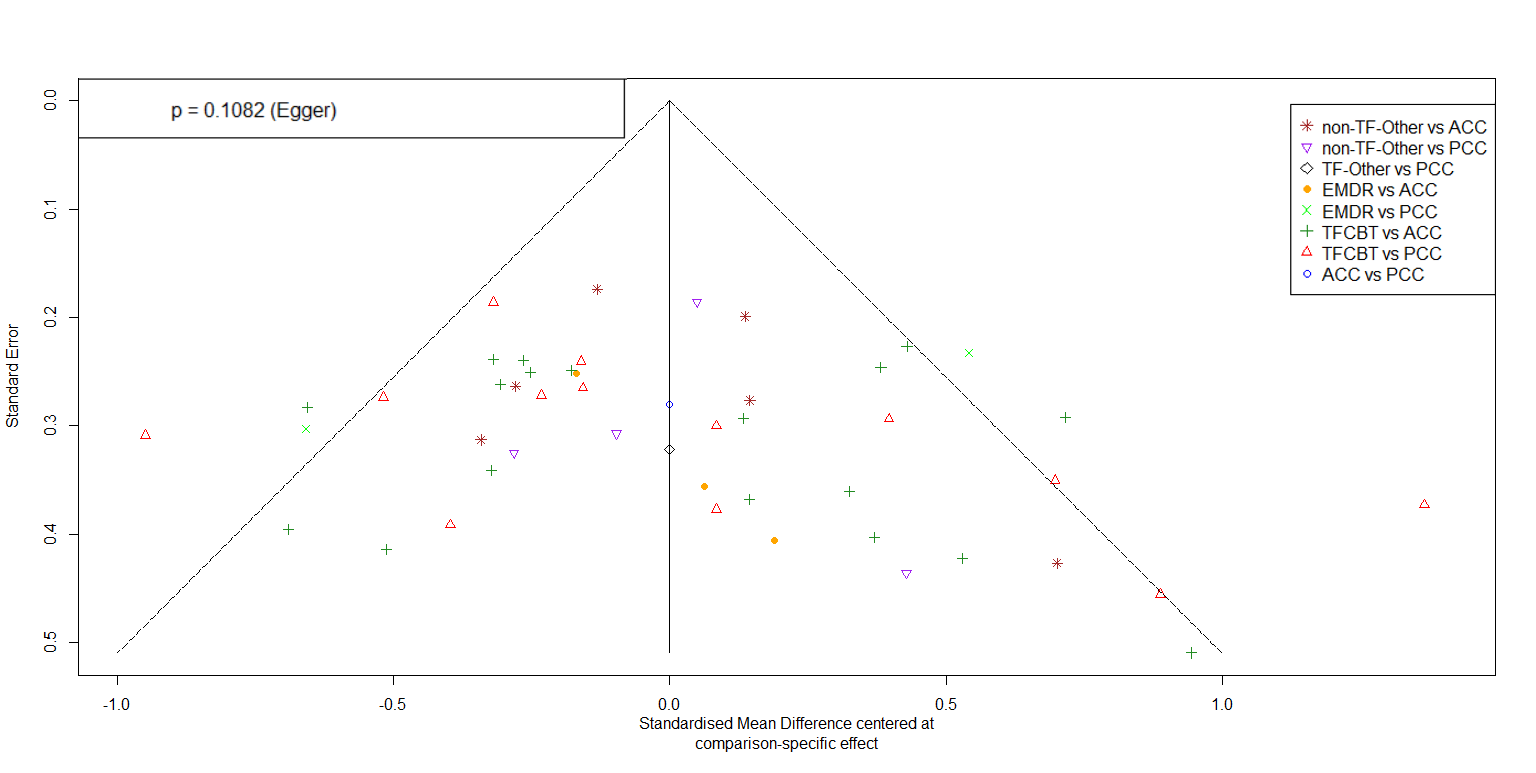

ACC – active control conditions (e.g., treatment-as-usual), EMDR – eye movement desensitization and reprocessing, FU1 – follow-up 1 (i.e., ≤ 5 months follow-up), non-TF-PIs – non-trauma-focused psychological interventions, other-TF-PIs – other trauma-focused psychological interventions (i.e., other than TF-CBT and EMDR), PCC – passive control conditions (e.g., waitlist), TF-CBT – trauma-focused cognitive behaviour therapy

**Appendix W: Netheat plot for NMA on efficacy at FU1 (i.e., ≤ 5 months follow-up)**

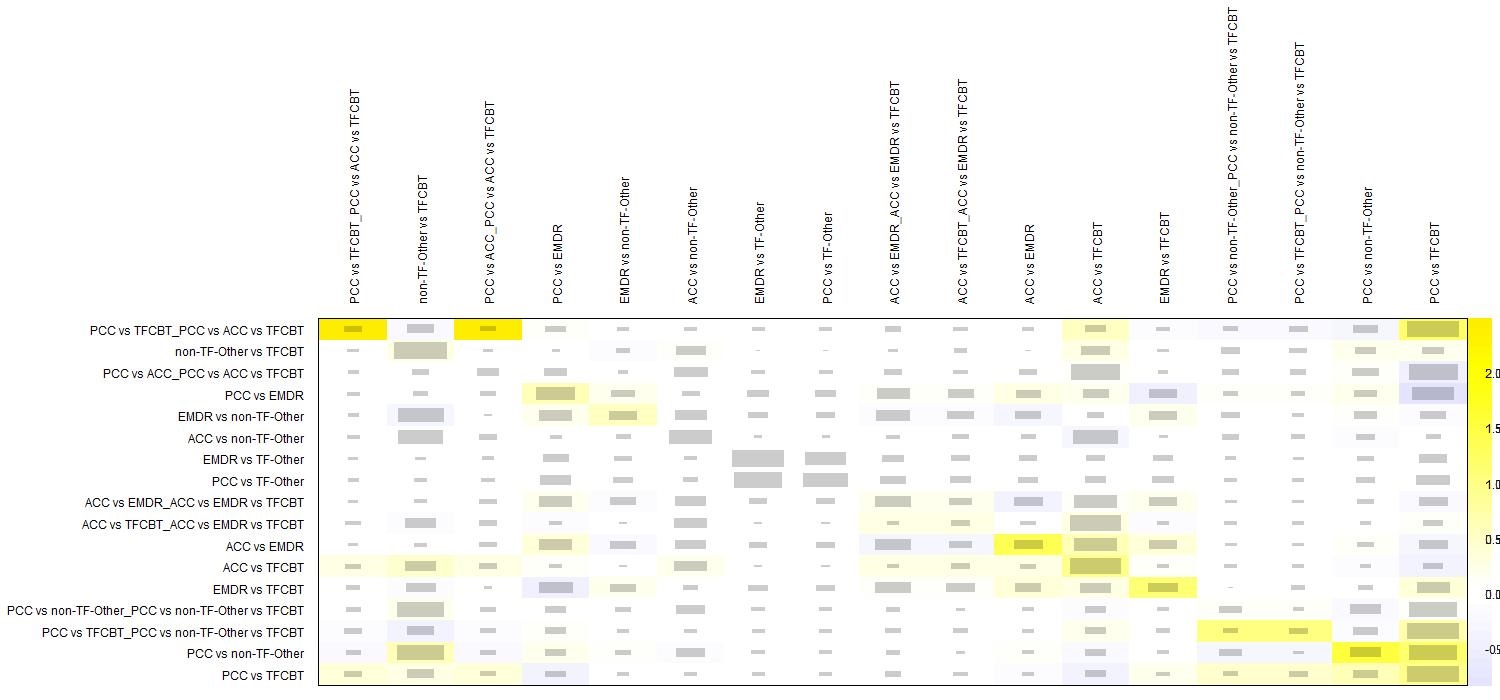

ACC – active control conditions (e.g., treatment-as-usual), EMDR – eye movement desensitization and reprocessing, FU1 – follow-up 1 (i.e., ≤ 5 months follow-up), non-TF-PIs – non-trauma-focused psychological interventions, other-TF-PIs – other trauma-focused psychological interventions (i.e., other than TF-CBT and EMDR), PCC – passive control conditions (e.g., waitlist), TF-CBT – trauma-focused cognitive behaviour therapy

**Appendix X: Outlier-corrected comparative long-term efficacy of psychological interventions (PIs)**

| *Reference group PIs* | | | *k (N)* | *g* | *95% CI* | *p* | *I^2^ (τ^2^)* |
| --- | --- | --- | --- | --- | --- | --- | --- |
|  | **FU1 (i.e., ≤ 5 months) - m*ain analysis*** | | | | | |  |
| relative to PCC | | TF-CBT# | 13 (713) | **0.83** | **0.63 – 1.03** | **< .001** | 56.50 *** (0.08) |
|  |  | EMDR | 2 (145) | **0.79** | **0.48 – 1.11** | **< .001** |  |
|  |  | other-TF-PIs | 1 (42) | **0.77** | **0.21 – 1.32** | **.007** |  |
|  |  | non-TF-PIs | 4 (256) | **0.59** | **0.37 – 0.82** | **< .001** |  |
|  | | ACC | 1 (52) | **0.31** | **0.07 – 0.55** | **.013** |  |
| relative to ACC | | TF-CBT | 18 (872) | **0.52** | **0.35 – 0.69** | **< .001** |  |
|  |  | EMDR | 3 (116) | **0.48** | **0.17 – 0.80** | **.002** |  |
|  |  | other-TF-PIs | 0 (0) | 0.46 | -0.12 – 1.03 | .122 |  |
|  |  | non-TF-PIs# | 6 (410) | 0.28 | 0.09 – 0.48 | **.004** |  |
| relative to EMDR | | TF-CBT | 4 (99) | 0.04 | -0.26 – 0.33 | .804 |  |
|  |  | other-TF-PIs | 1 (129) | -0.03 | -0.56 – 0.51 | .920 |  |
|  |  | non-TF-PIs | 1 (46) | -0.20 | -0.51 – 0.11 | .214 |  |
| relative to other-TF-PIs | | TF-CBT | 0 (0) | 0.07 | -0.50 – 0.63 | .822 |  |
|  |  | non-TF-PIs | 0 (0) | -0.17 | -0.75 – 0.40 | .559 |  |
| relative to non-TF-PIs | | TF-CBT | 17 (1,878) | **0.24** | **0.08 – 0.39** | **.002** |  |
|  | ***Sensitivity analysis: trials with individual face-to-face delivery of PIs only*** | | | | | |  |
| relative to PCC | | TF-CBT# | 11 (622) | **0.85** | **0.61 – 1.09** | **< .001** | 57.00 *** (0.10) |
|  |  | EMDR | 1 (102) | **0.87** | **0.51 – 1.24** | **< .001** |  |
|  |  | other-TF-PIs | 1 (42) | **0.81** | **0.22 – 1.41** | **.008** |  |
|  |  | non-TF-PIs | 1 (147) | **0.63** | **0.32 – 0.94** | **< .001** |  |
|  | | ACC | 1 (52) | **0.37** | **0.07 – 0.67** | **.017** |  |
| relative to ACC | | TF-CBT | 16 (727) | **0.48** | **0.28 – 0.69** | **< .001** |  |
|  |  | EMDR | 3 (116) | **0.50** | **0.15 – 0.85** | **.005** |  |
|  |  | other-TF-PIs | 0 (0) | 0.45 | -0.17 – 1.07 | .159 |  |
|  |  | non-TF-PIs | 1 (53) | 0.26 | -0.02 – 0.55 | **.071** |  |
| relative to EMDR | | TF-CBT | 4 (99) | -0.02 | -0.35 – 0.31 | .908 |  |
|  |  | other-TF-PIs | 1 (129) | -0.06 | -0.63 – 0.52 | .845 |  |
|  |  | non-TF-PIs | 1 (46) | -0.24 | -0.61 – 0.13 | .204 |  |
| relative to other-TF-PIs | | TF-CBT | 0 (0) | 0.04 | -0.57 – 0.64 | .902 |  |
|  |  | non-TF-PIs | 0 (0) | -0.18 | -0.81 – 0.45 | .568 |  |
| relative to non-TF-PIs | | TF-CBT | 11 (968) | **0.22** | **0.01 – 0.44** | **.045** |  |
|  | ***FU2 (i.e., > 5 months) - main analysis*** | | | | | |  |
| relative to PCC | | TF-CBT | 5 (399) | **0.69** | **0.44 – 0.95** | **< .001** | 37.00 *  (0.03) |
|  |  | EMDR | 1 (102) | **0.42** | **0.04 – 0.79** | **.030** |  |
|  |  | other-TF-PIs | 0 (0) | **0.61** | **0.21 – 1.00** | **.003** |  |
|  |  | non-TF-PIs | 0 (0) | **0.51** | **0.22 – 0.80** | **< .001** |  |
|  | | ACC | 1 (51) | 0.03 | -0.27 – 0.33 | .827 |  |
| relative to ACC | | TF-CBT# | 14 (647) | **0.66** | **0.48 – 0.84** | **< .001** |  |
|  |  | EMDR | 0 (0) | **0.38** | **0.01 – 0.76** | **.044** |  |
|  |  | other-TF-PIs | 0 (0) | **0.57** | **0.21 – 0.94** | **.002** |  |
|  |  | non-TF-PIs | 1 (102) | **0.47** | **0.26 – 0.69** | **< .001** |  |
| relative to EMDR | | TF-CBT | 2 (148) | 0.28 | -0.06 – 0.61 | .103 |  |
|  |  | other-TF-PIs | 1 (106) | 0.19 | -0.18 – 0.56 | .319 |  |
|  |  | non-TF-PIs | 0 (0) | 0.09 | -0.27 – 0.45 | .625 |  |
| relative to other-TF-PIs | | TF-CBT | 2 (204) | 0.09 | -0.24 – 0.41 | .598 |  |
|  |  | non-TF-PIs | 0 (0) | -0.10 | -0.45 – 0.25 | .580 |  |
| relative to non-TF-PIs | | TF-CBT | 13 (1,549) | **0.19** | **0.05 – 0.33** | **.009** |  |

#indicates that ≥ 1 outlier(s) was(were) detected and deleted for the outlier-corrected analyses. Bold prints highlight significant differences. *** p < .001, ** p < .01, * p < .05, correspond to the respective Q-statistic. ACC – active control conditions (e.g., treatment-as-usual), EMDR – eye movement desensitization and reprocessing, k – number of trials for the given comparison, N – number of participants for the given comparison, non-TF-PIs – non-trauma-focused psychological interventions, other-TF-PIs – other trauma-focused psychological interventions (i.e., other than TF-CBT and EMDR), PCC – passive control conditions (e.g., waitlist), PIs – psychological interventions, SMD – standardized mean differences (i.e., Hedges’ g), TF-CBT – trauma-focused cognitive behaviour therapy

**Appendix Y: Forest plot for NMA at FU2 (i.e., > 5 months follow-up) with passive control conditions as reference group**

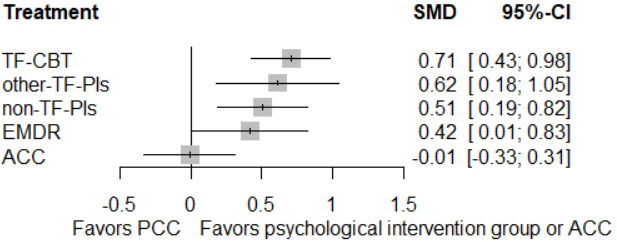

ACC – active control conditions (e.g., treatment-as-usual), EMDR – eye movement desensitization and reprocessing, FU1 – follow-up 2 (i.e., > 5 months follow-up), non-TF-PIs – non-trauma-focused psychological interventions, other-TF-PIs – other trauma-focused psychological interventions (i.e., other than TF-CBT and EMDR), PCC – passive control conditions (e.g., waitlist), TF-CBT – trauma-focused cognitive behaviour therapy

**Appendix Z: Forest plot for NMA at FU2 (i.e., > 5 months follow-up) with non-TF-PIs as reference group**

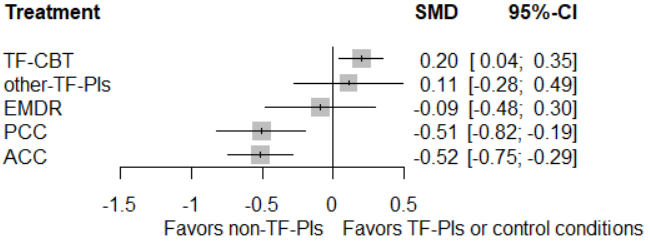

ACC – active control conditions (e.g., treatment-as-usual), EMDR – eye movement desensitization and reprocessing, FU1 – follow-up 2 (i.e., > 5 months follow-up), non-TF-PIs – non-trauma-focused psychological interventions, other-TF-PIs – other trauma-focused psychological interventions (i.e., other than TF-CBT and EMDR), PCC – passive control conditions (e.g., waitlist), TF-CBT – trauma-focused cognitive behaviour therapy

**Appendix AA: Funnel plot for NMA on efficacy at FU2 (i.e., > 5 months follow-up)**

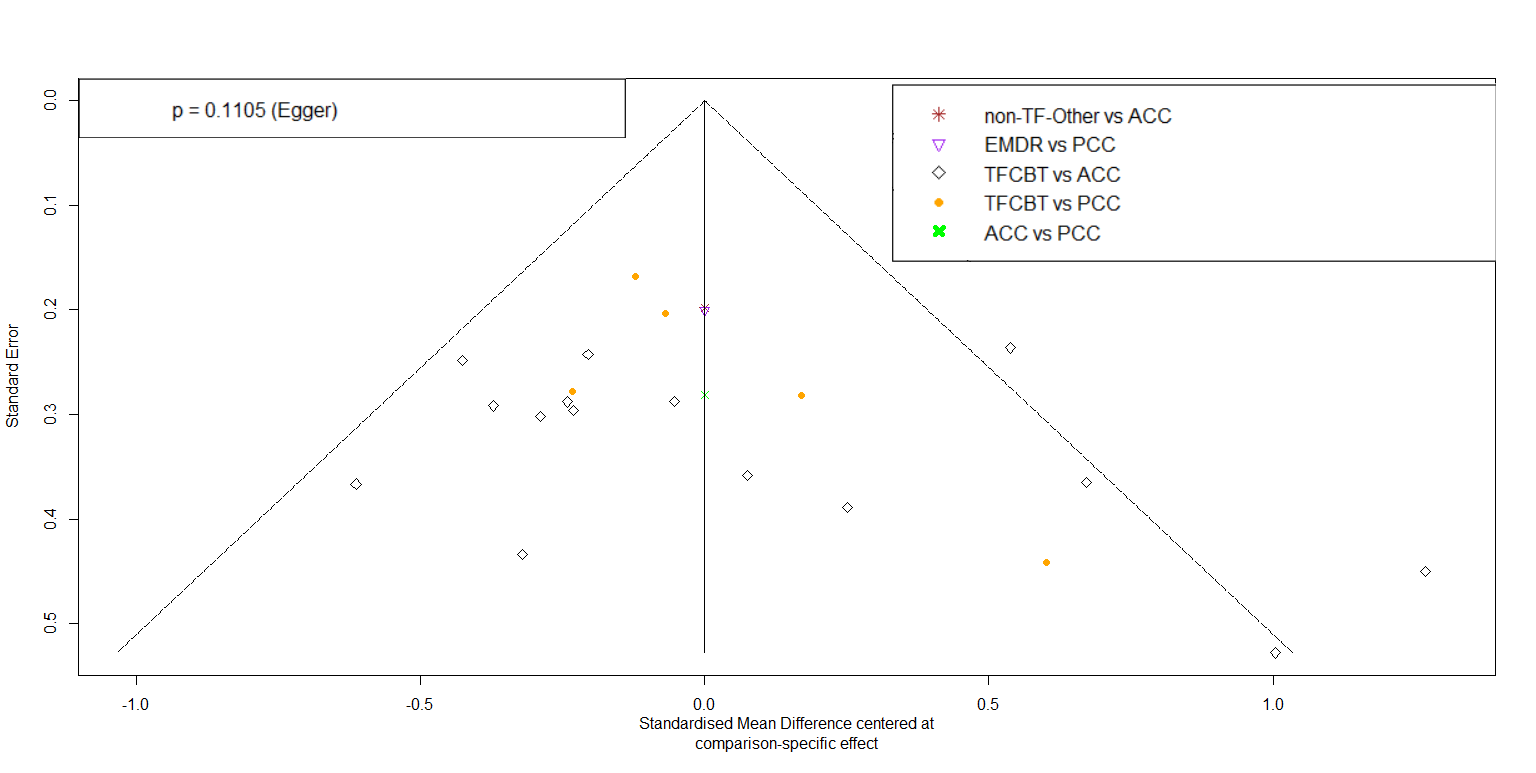

ACC – active control conditions (e.g., treatment-as-usual), EMDR – eye movement desensitization and reprocessing, FU1 – follow-up 2 (i.e., > 5 months follow-up), non-TF-PIs – non-trauma-focused psychological interventions, other-TF-PIs – other trauma-focused psychological interventions (i.e., other than TF-CBT and EMDR), PCC – passive control conditions (e.g., waitlist), TF-CBT – trauma-focused cognitive behaviour therapy

**Appendix AB: Outlier-adjusted funnel plot for NMA on efficacy at FU2 (i.e., > 5 months follow-up)**

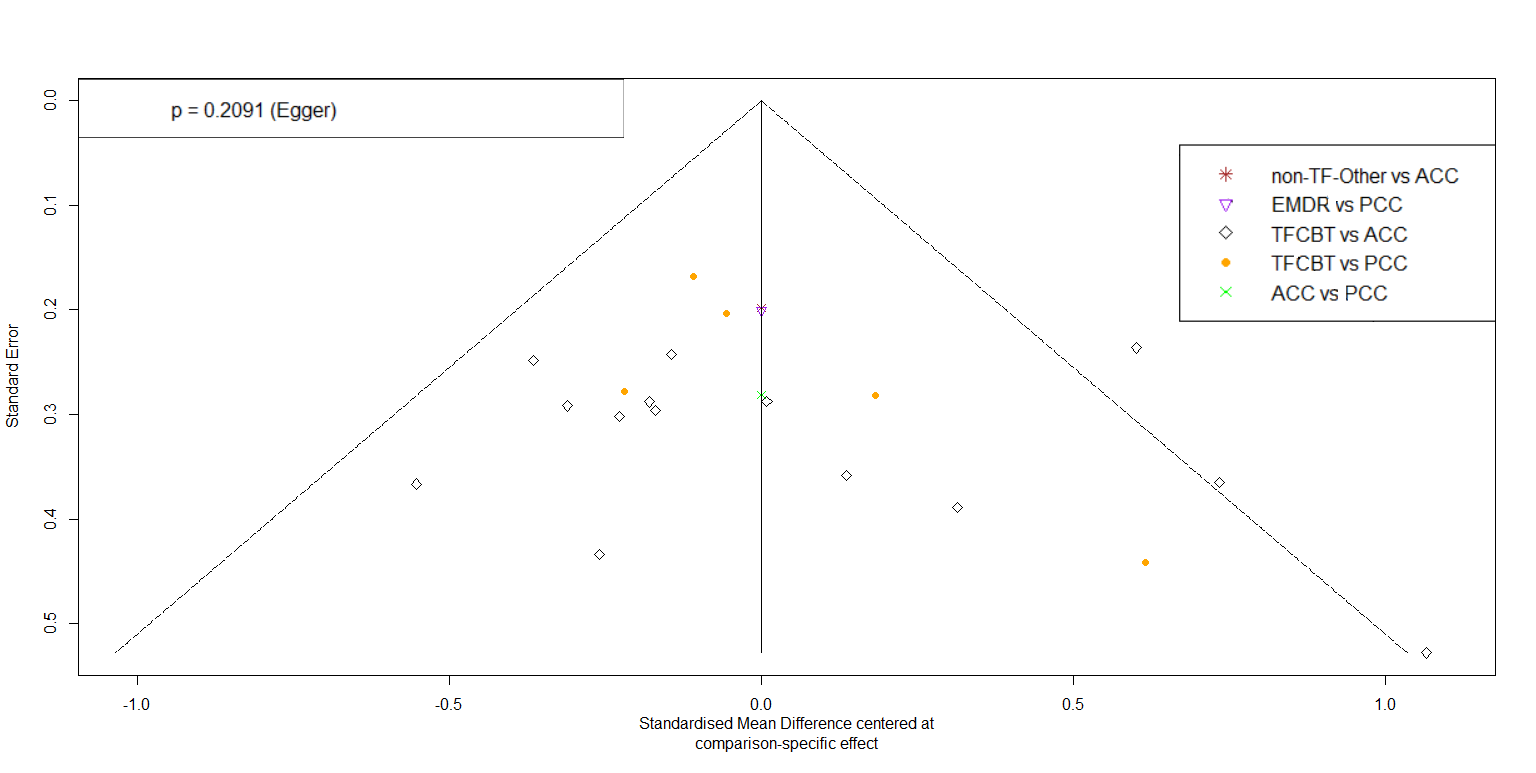

ACC – active control conditions (e.g., treatment-as-usual), EMDR – eye movement desensitization and reprocessing, FU1 – follow-up 2 (i.e., > 5 months follow-up), non-TF-PIs – non-trauma-focused psychological interventions, other-TF-PIs – other trauma-focused psychological interventions (i.e., other than TF-CBT and EMDR), PCC – passive control conditions (e.g., waitlist), TF-CBT – trauma-focused cognitive behaviour therapy

**Appendix AC: Netheat plot for NMA on efficacy at FU2 (i.e., > 5 months follow-up)**

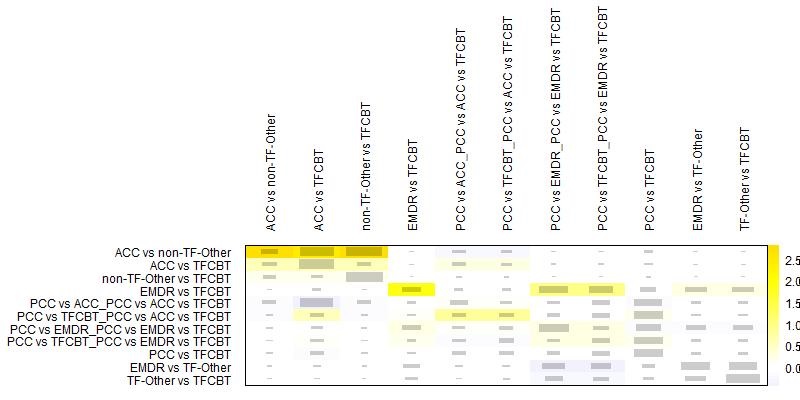

ACC – active control conditions (e.g., treatment-as-usual), EMDR – eye movement desensitization and reprocessing, FU1 – follow-up 2 (i.e., > 5 months follow-up), non-TF-PIs – non-trauma-focused psychological interventions, other-TF-PIs – other trauma-focused psychological interventions (i.e., other than TF-CBT and EMDR), PCC – passive control conditions (e.g., waitlist), TF-CBT – trauma-focused cognitive behaviour therapy
